# Translational modeling-based evidence for enhanced efficacy of standard-of-care drugs in combination with anti-microRNA-155 in non-small-cell lung cancer

**DOI:** 10.1101/2024.03.14.24304306

**Authors:** Prashant Dogra, Vrushaly Shinglot, Javier Ruiz-Ramírez, Joseph Cave, Joseph D. Butner, Carmine Schiavone, Dan G. Duda, Ahmed O. Kaseb, Caroline Chung, Eugene J. Koay, Vittorio Cristini, Bulent Ozpolat, George A. Calin, Zhihui Wang

**Affiliations:** Mathematics in Medicine Program, Department of Medicine, Houston Methodist Research Institute, Houston, TX, USA; Department of Physiology and Biophysics, Weill Cornell Medical College, New York, NY, USA; Princess Margaret Cancer Centre, Toronto, ON, Canada; Physiology, Biophysics, and Systems Biology Program, Graduate School of Medical Sciences, Weill Cornell Medicine, New York, NY, USA; Department of Radiation Oncology, The University of Texas MD Anderson Cancer Center, Houston, TX, USA; Department of Chemical, Materials and Industrial Production Engineering, University of Naples Federico II, Naples, Italy; Edwin. L. Steele Laboratories for Tumor Biology, Department of Radiation Oncology, Massachusetts General Hospital and Harvard Medical School, Boston, MA, USA; Department of Gastrointestinal Medical Oncology, The University of Texas MD Anderson Cancer Center, Houston, TX, USA; Neal Cancer Center, Houston Methodist Research Institute, Houston, TX, USA; Department of Imaging Physics, University of Texas M.D. Anderson Cancer Center, Houston, TX, USA; Department of Nanomedicine, Houston Methodist Research Institute, Houston, TX, USA; Department of Translational Molecular Pathology, The University of Texas MD Anderson Cancer Center, Houston, TX, USA

**Author notes:** **Correspondence should be addressed to: Prashant Dogra, Ph.D.**, Assistant Research Professor, Mathematics in Medicine, Houston Methodist Research Institute, Houston, TX, Or **Zhihui Wang, Ph.D.**, Professor, Mathematics in Medicine, Houston Methodist Research Institute, Houston, TX.

**Keywords:** non-small-cell lung cancer, microRNA, nanomedicine, mathematical modeling, pharmacokinetics, pharmacodynamics, RECIST 1.1, drug synergism, survival analysis

## Abstract

Elevated microRNA-155 (miR-155) expression in non-small-cell lung cancer (NSCLC) promotes cisplatin resistance and negatively impacts treatment outcomes. However, miR-155 can also boost anti-tumor immunity by suppressing PD-L1 expression. We developed a multiscale mechanistic model, calibrated with *in vivo* data and then extrapolated to humans, to investigate the therapeutic effects of nanoparticle-delivered anti-miR-155 in NSCLC, alone or in combination with standard-of-care drugs. Model simulations and analyses of the clinical scenario revealed that monotherapy with anti-miR-155 at a dose of 2.5 mg/kg administered once every three weeks has substantial anti-cancer activity. It led to a median progression-free survival (PFS) of 6.7 months, which compared favorably to cisplatin and immune checkpoint inhibitors. Further, we explored the combinations of anti-miR-155 with standard-of-care drugs, and found strongly synergistic two- and three-drug combinations. A three-drug combination of anti-miR-155, cisplatin, and pembrolizumab resulted in a median PFS of 13.1 months, while a two-drug combination of anti-miR-155 and cisplatin resulted in a median PFS of 11.3 months, which emerged as a more practical option due to its simple design and cost-effectiveness. Our analyses also provided valuable insights into unfavorable dose ratios for drug combinations, highlighting the need for optimizing dose regimen to prevent antagonistic effects. Thus, this work bridges the gap between preclinical development and clinical translation of anti-miR-155 and unravels the potential of anti-miR-155 combination therapies in NSCLC.

## Introduction

Non-small-cell lung cancer (NSCLC) is responsible for more than 80% of lung cancer cases (1), and remains a formidable challenge, predominantly due to the development of resistance to platinum-based chemotherapies, resulting in relapse and patient mortality (2). Notably, the overexpression of microRNA-155 (miR-155) in tumor cells has emerged as a key contributor to chemoresistance and is associated with increased tumor aggressiveness and unfavorable prognoses in NSCLC (3,4). Our group previously identified a novel mechanism by which miR-155 induces resistance to cisplatin through a TP53-mediated feedback loop (3). Additionally, high expression of miR-155 in tumor-associated macrophages (TAMs) and their secreted exosomes further contributes to miR-155 levels in cancer cells (5,6).

*In vivo* therapeutic targeting of miR-155 through its antagonist, anti-miR-155, using nanoparticle (NP)-mediated delivery, has shown considerable potential in controlling tumor growth and enhancing the efficacy of cisplatin (3). However, there are challenges associated with the translation of anti-miR-155 therapy. Given the suppressive effect of miR-155 on PD-L1 expression (7,8), there is an anticipated negative impact of anti-miR-155 on anti-cancer immunity through enhanced expression of the immune checkpoint programmed death ligand 1 (PD-L1). This could challenge the development of anti-miR-155 therapy, but also presents an opportunity since elevated PD-L1 expression often positively correlates with the prognosis of patients receiving immune checkpoint inhibitors (ICIs) in NSCLC (9,10). We thus hypothesized that combining anti-miR-155 with ICIs may yield synergistic effects, mutually enhancing their efficacy in NSCLC. The optimal dose ratios for this combination to prevent antagonistic effects will require careful evaluation. Furthermore, incorporating cisplatin into a three-drug combination holds the potential to further enhance the efficacy of the therapy. Addressing these questions necessitates an in-depth exploration of dose-response relationships and performing drug combination studies.

To this end, we have taken a modeling and simulation-based approach. Mathematical modeling has previously been shown to be a useful tool to investigate the mechanisms relevant to tumor response to miRNA-based treatments. For instance, Aguda et al. developed a system of ordinary differential equations (ODEs) with a delay term to study feedback loops between the oncogenes Myc, EF2, and miR-17-92 (11). This model was then expanded by integrating 9 different mechanisms, aiming to explain how miRNAs regulate translation (12), and to study how the inactivation of a transcription factor is involved in cancer and cardiac dysfunction (13). In another notable study (14), an energy availability pathway involving miR-451 was analyzed in order to elucidate the difference between invasion and proliferation regimes in cancer cells. The model provided an explanation for the growth-invasion cycling patterns of glioma cells in response to high/low glucose uptake in the tumor microenvironment, and suggested new targets for drugs, associated with miR-451 upregulation. In addition to these examples, a signaling pathway relating miR-21, miR-155, and miR-205 to the proliferation and apoptosis of NSCLC cells has been studied with a series of modeling studies (15,16). The authors identified a positive correlation between expression of these miRNAs in tumor tissue and blood, thereby establishing these miRNAs as potential serum biomarkers for early detection of NSCLC.

While previous modeling efforts focused on the molecular effects of miRNAs on tumor growth dynamics, they lacked the inclusion of a drug delivery system, its associated pharmacokinetics, and the transport phenomena necessary to assess the anti-cancer therapeutic efficacy of miRNAs or their antagonists. We previously developed a multiscale model of tumor growth dynamics integrated to a pharmacokinetic model to evaluate the translational potential of exogenously delivered miR-22 in triple-negative breast cancer (17). In the present study, we have extended this modeling work to further incorporate tumor-immune interactions and related molecular effects of miR-155 to evaluate the translational potential of anti-miR-155 in early-stage NSCLC, both as a monotherapy and in combination with standard-of-care drugs. The model has been calibrated with *in vivo* data and extrapolated to humans for translationally relevant simulations and analyses. We also performed sensitivity analyses to identify the determinants of tumor response and simulated clinically relevant treatment scenarios in a virtual patient cohort to establish dose-response relationships and evaluate drug combinations for possible synergistic effects.

Our study highlights the capacity of anti-miR-155 to enhance the efficacy of standard-of-care drugs in NSCLC. It also provides insights into optimizing dosing conditions to maximize therapeutic benefits while minimizing antagonistic effects. Importantly, given the favorable safety of cobomarsen, an miR-155 inhibitor, reported in a phase 1 clinical trial (18), our computational investigation yields valuable quantitative insights that support the progression of anti-miR-155 to advanced clinical trials for NSCLC patients.

## Results and Discussion

### Model development, parameterization, and calibration

We developed a multiscale mechanistic model of tumor response to systemic therapies to investigate the translational potential of nanoparticle (NP)-delivered anti-miR-155 therapy for use in combination with standard-of-care chemotherapy and/or ICI immunotherapies in NSCLC. Extending our previous work (17,19,20), the current model comprises two major compartments, as illustrated in **Fig. 1**: the plasma compartment, representing systemic circulation and the site of drug administration, and the tumor compartment, containing cancer cells and infiltrating immune cells, such as tumor-associated macrophages (TAMs) and cytotoxic CD8+ T cells that are recruited from the plasma compartment. At the molecular scale, the model incorporates immune checkpoint receptor-ligand interactions and the influence of miR-155, a critical player in NSCLC, on tumor proliferation, PD-L1 expression, and chemoresistance (3,7,8). The detailed processes of drug transport, cellular processes, biophysical interactions, and molecular effects are described by a set of mechanistic equations (**Equations S1–S29**; see **Supplementary Methods**).

**Figure 1.**
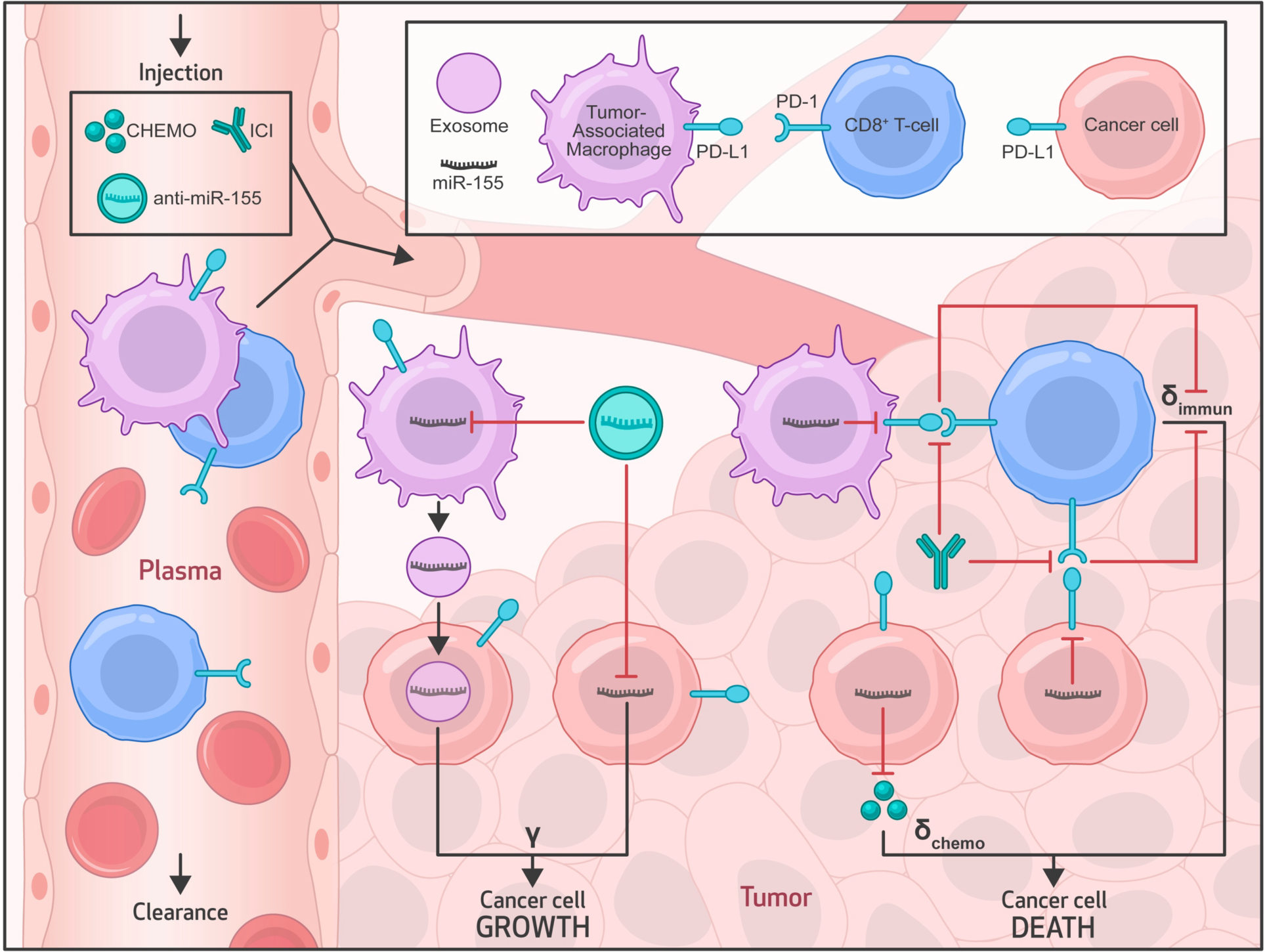
Multiscale mechanistic model. Model schematic shows the key transport processes, system interactions, and model variables in the plamsa and tumor compartments. The plasma compartment characterizes the systemic pharmacokinetics of drugs and nanoparticles, and serves as the source of therapeutics and immune cells (tumor-associated macrophages (TAMs), CD8+ T cells) for the tumor. In the tumor compartment, molecular-scale interactions of miR-155 induce tumor proliferation, chemoresistance, and suppression of PD-L1 expression in tumor cells and TAMs. miR-155 is overexpressed in tumor cells and TAMs, where the latter additionally supplies miR-155 to tumor cells via exosomal secretions. Mechanisms of CD8+ T cell-mediated tumor immunosurveillance and PD1/PD-L1 signaling-mediated immune escape are also modeled. *γ*, *δ*_chemo_, and *δ*_immun_ represent tumor proliferation rate, tumor death rate due to chemotherapy, and tumor death rate due to immunosurveilance by CD8^+^ T cells, respectively. Abbreviations: ICI-immune checkpoint inhibitor, CHEMO-chemotherapy.

To ensure that our model can reproduce reference biological behavior and remain consistent with physiological constraints observed *in vivo*, the baseline values of model parameters (detailed in **Tables S1** and **S2** of **Supplementary Methods**) were either derived from existing literature for mice or estimated through nonlinear least squares fitting of the model to experimental data retrieved from literature. To enhance the statistical identifiability of model parameters and reduce uncertainty in parameter estimation, we augmented the data volume by combining multiple datasets derived from diverse preclinical studies. This approach also allowed us to capture the inherent biological variability that exists among experimental settings and tumor characteristics, improving the applicability and adaptability of the model to diverse scenarios. As a result, the gathered datasets encompassed treatments of mice engrafted with patient-derived xenografts or cell line xenografts of NSCLC through various modalities, including anti-miR-155-loaded liposomes (3), cisplatin (3), a combination of anti-miR-155 and cisplatin (3), anti-PD-L1 antibody atezolizumab (21), and anti-PD-1 antibody pembrolizumab (22). This approach also ensured that the model can reliably simulate treatments involving both anti-miR-155 therapy and standard-of-care drugs, thereby allowing us to conduct combination therapy experiments *in silico*. The overall calibration process involved initialization of model equations with appropriate initial conditions (**Table S3**) and implementation of treatment regimens obtained from the corresponding experimental studies to predict tumor response to various therapies, followed by fitting of the model solutions to the pooled experimental tumor growth kinetics data to estimate unknown parameters.

To begin with, as indicated by the kinetics of NP mass in systemic circulation, i.e., the plasma compartment (**Fig. 2A**), we simulated the experimental treatment protocol involving twice-a-week injections of NPs loaded with 4,000 ng of anti-miR-155 (i.e., 0.2 mg/kg), starting at one week following inoculation of NSCLC cells in mice (3). The model captured the systemic PK of NPs, which is primarily governed by a combination of hepatobiliary excretion (characterized by *k*_Cl_) and metabolic degradation (characterized by *δ*_NP_). It also reliably predicted NP delivery to the tumor interstitium, which peaked at ∼3% of the injected dose (%ID) of NPs for every injection (**Fig. 2A**, **inset**), dictated by tumor microvascular surface area (*S*) and microvascular permeability to NPs (*P*_NP_). Following the diffusion of NPs through a characteristic length (Len) of the intercapillary distance in tumor interstitium, NPs are taken up by tumor cells and TAMs leading to the intracellular release of cargo, i.e., anti-miR-155. The corresponding intracellular concentration of anti-miR-155 exhibited a gradual increase over time, mostly ranging between 0.5–1.5 mg/mL during treatment, before starting to decline with the end of treatment (**Fig. 2B**).

**Figure 2.**
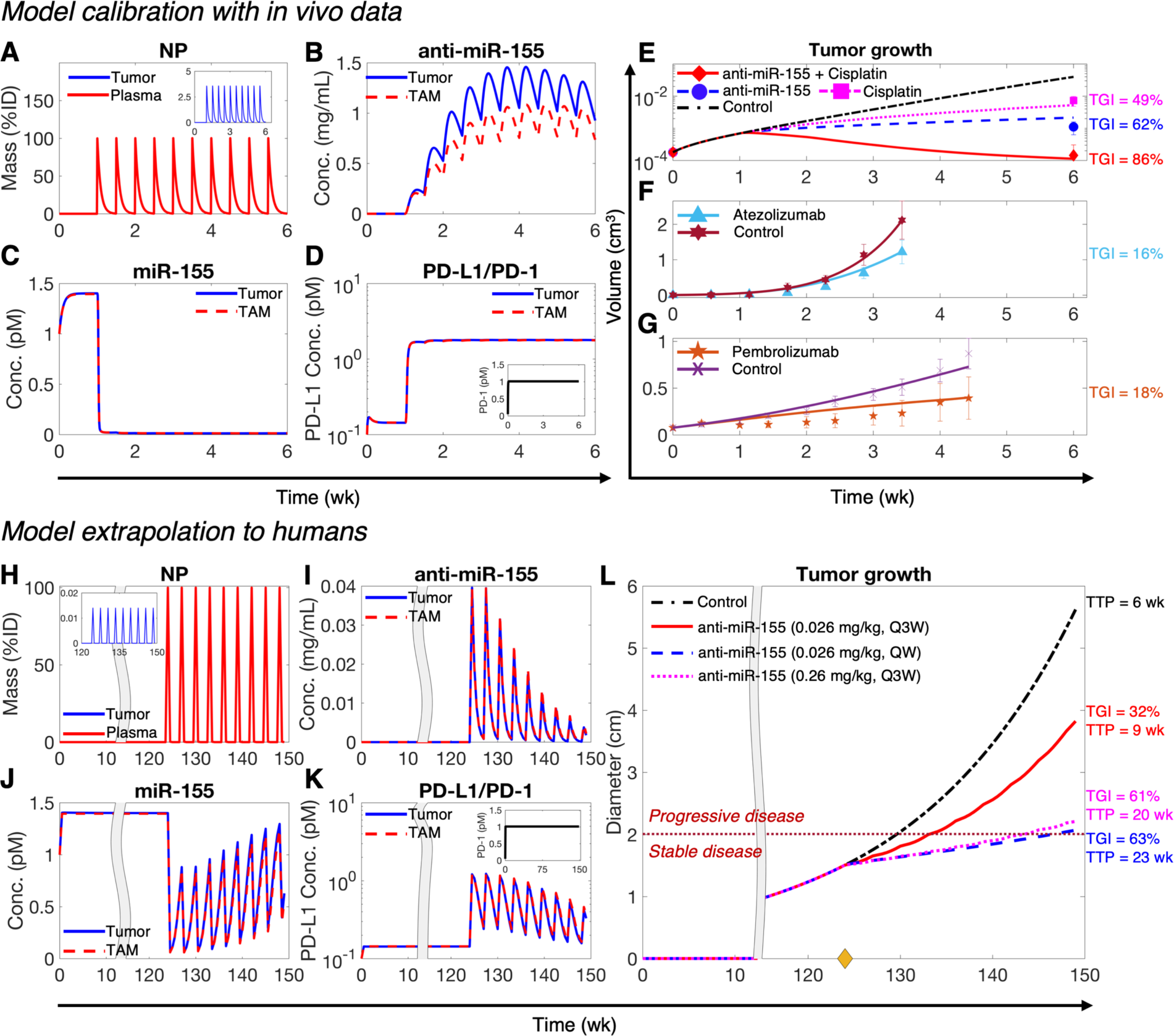
Model calibration and extrapolation to humans. **A-D)** Numerical solution of the model exhibiting kinetics of key variables under treatment with NP-delivered anti-miR-155 in NSCLC-bearing mice. **A)** Mass kinetics of NPs in plasma and tumor interstitium (inset) following twice-a-week injection of NPs loaded with a dose of 4,000 ng of anti-miR-155. %ID represents percent of injected dose. **B)** Concentration kinetics of NP-delivered anti-miR-155 in tumor cells and TAMs. **C)** Concentration kinetics of miR-155 in tumor cells and TAMs. **D)** Concentration kinetics of unbound PD-L1 on tumor cells and TAMs, and unbound PD-1 on CD8+ T cells (inset). **E-G)** Non-linear least squares fits of the model to published in vivo datasets of tumor volumetric growth kinetics in NSCLC under control conditions and treatment with **E)** anti-miR-155, cisplatin, combination of anti-miR-155 and cisplatin, **F)** atezolizumab, **G)** and pembrolizumab. **H-K)** Numerical solution of the allometrically scaled model showing key system variables following treatment with anti-miR-155 loaded NPs *in an average adult patient*. **H)** Mass kinetics of NPs in plasma and tumor interstitium (inset) following once every three weeks (Q3W) injection of NPs loaded with *the allometrically scaled* dose *of* anti-miR-155 *in humans (i.e.,* 0.026 mg/kg). **I)** Concentration kinetics of NP-delivered anti-miR-155 in tumor cells and TAMs. **J)** Concentration kinetics of miR-155 in tumor cells and TAMs. **K)** Concentration kinetics of unbound PD-L1 on tumor cells and TAMs, and unbound PD-1 on CD8+ T cells (inset). **L)** Corresponding tumor growth kinetics under control and treatment conditions involving anti-miR-155 at three different dosages (0.026 mg/kg, Q3W; 0.026 mg/kg, QW (once weekly); 0.26 mg/kg, Q3W). The yellow diamond on x-axis marks the initiation of treatment; the dotted dark red line defines the threshold to transition from stable disease to progressive disease, as per RECIST 1.1. Abbreviations: TGI-tumor growth inhibition, TTP-time to progression.

The concentration of miR-155 in cancer cells and TAMs declined rapidly upon intracellular delivery of anti-miR-155 (**Fig. 2C**). This is expected due to binding of the complementary anti-miR-155 oligonucleotides to miR-155, which enhances its degradation. It can be observed that miR-155 was almost completely depleted in both cell types, with levels (∼0.01 pM) maintained throughout the duration of treatment. Of note, prior to anti-miR-155 treatment, the concentration of miR-155 saturated to ∼1.5 pM in cancer cells and TAMs, which can be attributed to the dynamic equilibrium between miR-155 synthesis and degradation. Given that miR-155 is a negative regulator of PD-L1 (7,8), the predicted expression level of PD-L1 on the surface of tumor cells and TAMs increased by an order of magnitude after intracellular miR-155 suppression (**Fig. 2D**). The expression of the corresponding PD-1 receptor on tumor-infiltrating cytotoxic T cells, which is not mediated by miR-155, remained stable at its equilibrium value of 1 pM (**Fig. 2D**, inset).

By incorporating the paradoxical roles of miR-155 in inducing cancer cell proliferation and suppressing PD-L1 expression to enhance T cell-mediated tumor cell death, the model captures the net effect of anti-miR-155 therapy on tumor response. The resulting predictions of volumetric tumor growth were fitted to experimental data. Treatment with anti-miR-155 monotherapy inhibited tumor growth (**Fig. 2E**), leading to a ∼62% tumor growth inhibition (TGI), relative to the control case by the end of treatment. TGI is defined as the change in tumor size under treatment with respect to the control scenario (see **Methods** for details). Consistent with a key role of miR-155 in mediating cisplatin resistance in NSCLC (3), simulation of the combined cisplatin and anti-miR-155 therapy exhibited a stronger TGI (∼86%), which was greater than the TGI for anti-miR-155 (∼62%) and cisplatin (∼49%) monotherapies (**Fig. 2E**). These data are consistent with the chemosensitization effect of anti-miR-155 and potential synergism with cisplatin, which was studied subsequently in greater detail. The simulated regimen, based on experimental data (3), involved once weekly IV injection of 8 mg/kg cisplatin in the mono- and combination therapy scenarios; the corresponding systemic and intratumoral PK predictions and the kinetics of molecular variables are shown in **Figs. S1** and **S2**.

In addition, the model was employed to simulate the treatment of NSCLC-bearing mice with 10 mg/kg anti-PD-L1 atezolizumab (**Fig. S3**) and 5 mg/kg anti-PD-1 pembrolizumab (**Fig. S4**) ICI, which was then fitted to the experimental data. As shown in **Fig. 2F,G**, atezolizumab and pembrolizumab led to a ∼16% and ∼18% TGI by the end of treatment, respectively. Overall, the combined fits of predicted volumetric tumor growth for the five treatment scenarios and two controls were in close agreement with experimental data, as indicated by a strong Pearson correlation coefficient *R>*0.99 and *p*<0.0001 (**Fig. S6**), thereby enabling confidence in the predictive accuracy of the model.

### Model extrapolation to humans

We next sought to extrapolate the calibrated model to humans for assessing the translational potential of anti-miR-155 and for testing our hypothesis that adding anti-miR-155 therapy can increase the efficacy of standard-of-care drugs in NSCLC. To achieve this, we substituted the values of parameters for mice with population averages for humans from literature, or allometrically scaled the values of unknown parameters from mice to humans using established scaling factors (**Tables S1, S2**; see **Interspecies Scaling** in **Methods**). **Figure 2H-L** shows a representative simulation for treating an average patient (i.e., defined by baseline values of biological and physiological parameters, **Tables S1, S2**) with an allometrically scaled human equivalent dose (scaled from mice) of anti-miR-155. Here, we simulated a ∼six-month treatment regimen involving 0.026 mg/kg dose of anti-miR-155 given once every three weeks (Q3W) for nine treatment cycles, starting at 124 weeks post-tumor inception with a single cancer cell. Note that the tumor had a diameter of ∼1.5 cm at the time of initiation of treatment (**Fig. 2L**), representing a localized NSCLC tumor that can be classified as stage IA2 as per the TNM staging system (a standardized clinical system used for cancer staging) (23).

As shown in **Fig. 2H**, the model realized the intended treatment regimen and predicted the systemic and tumor interstitial PK of NPs over a course of nine treatment cycles. The subsequent delivery of anti-miR-155 into tumor cells and TAMs exhibited a highly fluctuating concentration profile of intracellular anti-miR-155, almost completely depleting between injections due to metabolic degradation of anti-miR-155 (**Fig. 2I**). Unlike the preclinical scenario where mice were injected with anti-miR-155 twice per week, allowing greater accumulation of drug intracellularly (**Fig. 2B**), the reduced treatment frequency in the clinical scenario explains the strong fluctuations in drug concentration. Furthermore, due to a continuously increasing tumor volume, the peak concentration of anti-miR-155 in the tumor decreased with each subsequent dose, as indicated by the regularly declining amplitude of the concentration profile of anti-miR-155 (**Fig. 2I**). To verify these interpretations, we simulated a once weekly (QW) injection of the same dose of anti-miR-155, and as shown in **Fig. S7B**, the intracellular concentration of anti-miR-155 exhibited a relatively steady profile, with concentration values mostly maintained between 0.1–0.25 mg/mL throughout the six-month duration of treatment.

The preclinical scenario showed that a relatively more stable and higher concentration of anti-miR-155 (**Fig. 2B**) was achieved intracellularly, leading to a more consistent suppression of miR-155 and elevation of PD-L1 over the course of treatment (**Fig. 2C,D**). In contrast, with the Q3W regimen, strong fluctuations were observed in the expression kinetics of miR-155 (**Fig. 2J**) and PD-L1 (**Fig. 2K**) in the average patient, corresponding to the fluctuations of anti-miR-155. However, the average behavior was consistent with the expected effect of anti-miR-155 on miR-155 and PD-L1. Upon increasing the treatment frequency to QW, the fluctuations disappeared and the suppression and elevation of miR-155 and PD-L1, respectively, was more stable throughout the course of treatment (**Fig. S7C,D**). The resultant therapeutic effect of anti-miR-155 on tumor growth corresponded to a ∼32% TGI at the end of treatment in the Q3W scenario, which was less than the QW scenario that exhibited (∼63% TGI) (**Fig. 2L**). This difference in therapeutic efficacy can be attributed to a higher intracellular concentration of anti-miR-155 and a more uniform suppression of miR-155 in the latter case.

These treatments were also assessed using RECIST 1.1 to measure time-to-progression (TTP), which is a clinical endpoint defined as the length of time after the initiation of treatment when a patient’s disease starts to progress and does not remain stable (24). TTP was determined by monitoring the change in tumor diameter from baseline (see **Treatment response evaluation** in **Methods** for details). If the change in diameter from the initial tumor exceeded 20%, with a minimum change of 5 mm, it was considered as progressive disease, as indicated by the threshold marked by the dotted dark red line in **Fig. 2L**. Our observations revealed that the Q3W dosing scenario exhibited a shorter TTP of ∼9 weeks compared to the QW scenario, which had a TTP of ∼23 weeks. Additionally, the control case exhibited a TTP of < 6 weeks. Notably, we also observed that a higher TGI of ∼61% and an extended TTP of ∼20 weeks could also be achieved with a Q3W injection frequency, provided that a dose of anti-miR-155 at an order of magnitude higher (0.26 mg/kg) was administered (see **Figs. 2L, S8**). These findings underscore the importance of establishing the dose-response relationship of anti-miR-155 to guide appropriate dose selection for maximal therapeutic efficacy.

We note that these observations reflect outcomes in an average patient, hence they cannot be generalized to a diverse patient population. It is thus important to quantify the uncertainty in these findings to account for the effect of population-scale variability in biological factors. To address this, we examined the effect of treatment regimens on tumor response, while accounting for variability in model parameter values, as discussed in the next section.

### Parameter sensitivity analyses

To investigate the effect of various treatment regimens on tumor response to anti-miR-155 therapy, we first sought to identify biological parameters with the most significant impact on tumor response to therapy, followed by the simulation of a comprehensive dose-response study to quantify uncertainty in model predictions due to biological variability of the identified model parameters. Therefore, we first conducted a global sensitivity analysis (GSA) and a local sensitivity analysis (LSA). GSA involves simultaneous perturbation of multiple model parameters, whereas LSA involves perturbation of individual parameters. The parameters were perturbed over a range of ±50% of their respective baseline values to simulate scenarios that deviated from the average behavior of the model and calculate TGI. For GSA, this was followed by calculation of sensitivity indices (SI), which are a measure of parameter importance in determining model outcomes (see **Methods**). The 28 model parameters investigated during GSA involved parameters that characterize the biological processes, interactions, and therapy effects involved in governing tumor dynamics.

As shown in **Fig. 3A**, based on the results of one-way ANOVA and Tukey’s test on SI, the investigated parameters were rank-ordered and separated into eight categories, exhibiting their relative impact on treatment response (i.e., TGI) to anti-miR-155. The top two categories comprise parameters *γ* and *A_M,L_* that directly influence tumor proliferation. Here, the intrinsic tumor growth rate *γ*, as observed via LSA (inset), is inversely correlated to TGI, exhibiting an inverse S-shaped curve. This indicates that within the investigated parameter range (±50% of the baseline value), rapidly growing tumors are associated with poorer response. This is consistent with observations in literature, where pre-treatment tumor growth rate of NSCLC and other solid tumors has been found to negatively affect treatment outcomes to ICIs (25,26) and chemoradiotherapy (27–29). This can be attributed to timescale differences in tumor growth- and death-related processes, leading to poorer response due to predominance of growth in aggressive tumors. Furthermore, the delivery of anti-miR-155-loaded NPs from systemic circulation to the tumor interstitium is governed by the product of vascular permeability *P*_NP_ and vascular surface area per unit volume *S* (**Eq. S12**). Because larger tumors have a relatively smaller vascular surface area per unit volume for extravasation of NPs (30), poorer outcomes with anti-miR-155 therapy in fast-growing tumors are expected due to reduced drug delivery and tumor exposure. However, note that the relation between TGI and *γ* is non-monotonic, such that beyond ∼15% reduction in *γ* from the baseline, tumor response to anti-miR-155 gradually deteriorates with decreasing *γ*, which points out to the inherent non-linearity of the model system. Next, the coefficient *A_M,L_* which modulates the tumor proliferation-inducing effect of miR-155, follows *γ* in ranking. Since *A_M,L_* (value >1) positively regulates tumor proliferation (**Eq. S10**), increasing the value of *A_M,L_* leads to rapidly growing tumors, thereby *A_M,L_* shows a behavior that is analogous to the *γ* parameter, as observed through LSA (**Fig. 3A**, inset).

**Figure 3.**
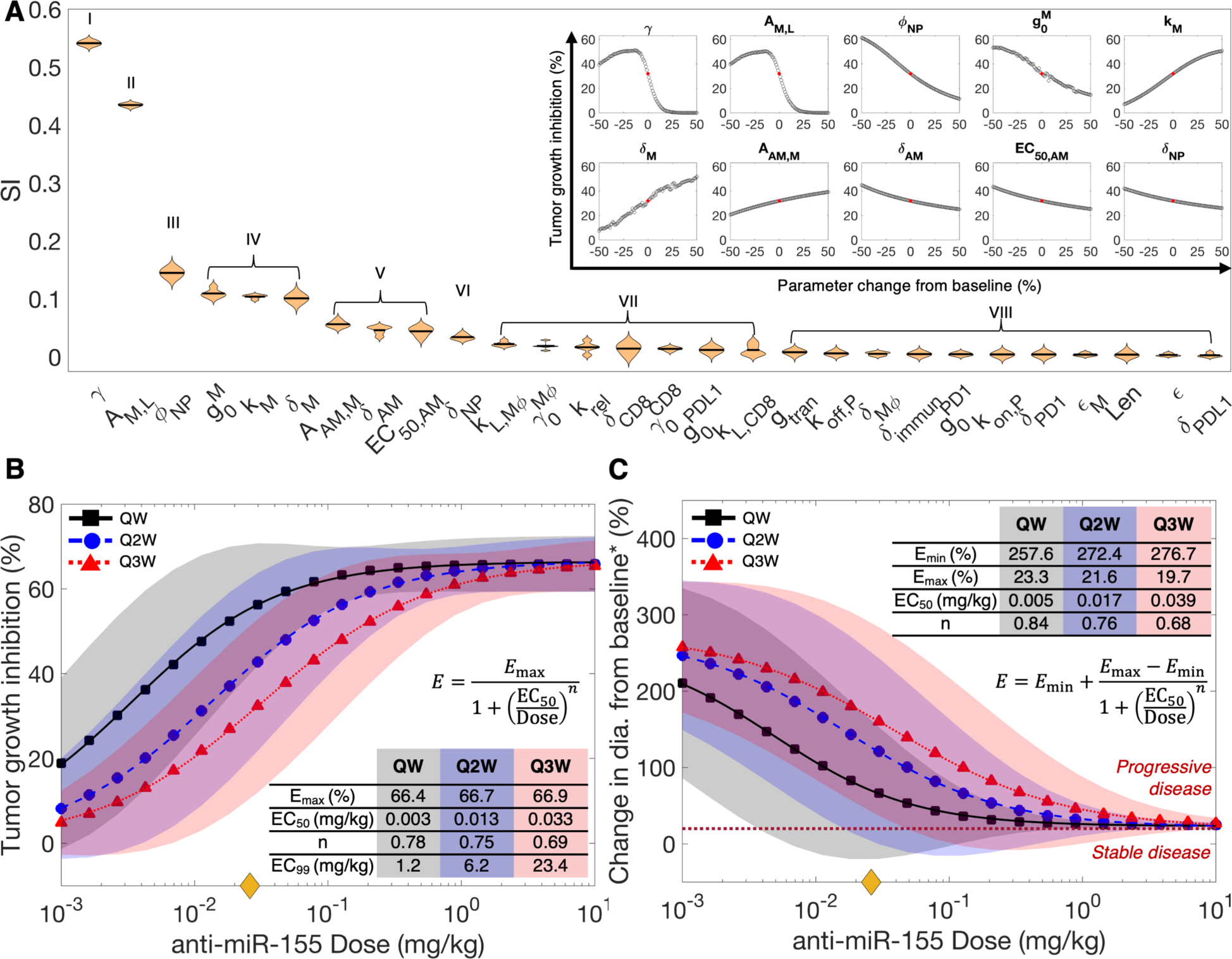
Sensitivity analyses. **A)** Violin plot displaying the ranking of model parameters for their impact on anti-miR-155-induced tumor growth inhibition (TGI), as obtained from global sensitivity analysis (GSA). Multivariate linear regression analysis-based regression coefficients (labeled as sensitivity indices (SI)) were used to rank order the parameters using one-way ANOVA and Tukey’s test. Inset shows results of local sensitivity analysis (LSA) of parameters that significantly affect model output, as determined from GSA. The red dot denotes TGI obtained with baseline parameter values, i.e., without perturbation. **Dose-response relationship.** To evaluate the effect of treatment regimen (i.e., anti-miR-155 dose and treatment frequency) on tumor response, **B)** TGI and **C)** change in tumor diameter from baseline* were predicted for different doses and injection frequencies (QW-once weekly, Q2W-once every 2 weeks, and Q3W-once every 3 weeks). **B)** The Hill equation was fit to TGI versus dose curves to quantitatively characterize the dose-response curves, with parameter estimates shown in the inset. The yellow diamond on the *x*-axis denotes the allometrically scaled dose of 0.026 mg/kg. **C)** RECIST 1.1 was used to characterize tumor response as per clinical standards and the modified Hill equation was fit to the results. The dotted dark-red line defines the threshold to transition from stable disease to progressive disease, as per RECIST 1.1. **\***Baseline refers to the tumor diameter at the time of treatment initiation or the smallest tumor diameter recorded since the beginning of treatment, depending on the specific response category being considered, as defined by RECIST 1.1 guidelines. Shaded regions denote 90% prediction intervals.

Following this in ranking is the diameter of NPs *Φ*_NP_, which together with the size of microvascular wall pores governs vascular permeability of NPs *P*_NP_, such that smaller NPs permeate more effectively and lead to more efficient delivery of anti-miR-155 to the target cells, thereby causing improved treatment response, as indicated by the negative correlation observed between *Φ*_NP_ and TGI (**Fig. 3A**, inset).

Next, miR-155-related parameters (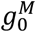, *k_M_*, and *δ_M_*), part of group IV, are also pivotal in determining the efficacy of anti-miR-155 therapy. Enhanced production of miR-155 governed by its production rate 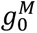, or reduced degradation of miR-155 governed by its degradation rate *δ_M_* leads to poorer tumor outcomes, as indicated by the negative and positive correlations of 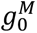 and *δ_M_*, respectively, with TGI (**Fig. 3A**, inset). This is attributable to the increased expression of miR-155 in cancer cells and TAMs, which positively affects tumor proliferation, depending upon its Michaelis constant or potency (*k_M_*) and the previously discussed stimulation coefficient *A_M,L_* (**Eq. S10**), leading to poorer outcomes. However, the greater the *k_M_*, the less potent is miR-155 in inducing tumor growth, leading to improved outcomes to anti-miR-155 therapy, hence the positive correlation between *k_M_* and TGI (**Fig. 3A**, inset).

Finally, categories V and VI comprising *A*_AM,*M*_, *δ*_AM_, EC_50,AM_, and *δ*_NP_ are the last set of parameters that significantly impact tumor response to anti-miR-155 therapy. These parameters influence the tumoral concentration, potency, and efficacy of anti-miR-155 in cancer cells and TAMs. Since anti-miR-155 acts by enhancing the degradation rate *δ_M_* of miR-155 (**Eqs. S1 and S4**), its effectiveness is modulated by the coefficient *A*_AM,*M*_ (value >1), such that it correlates positively with TGI (**Fig. 3A**, inset). Furthermore, anti-miR-155 degradation rate *δ*_AM_ acts by influencing the concentration of anti-miR-155 in cells, such that increased value of *δ*_AM_ will lead to reduced concentration of anti-miR-155 in the cells, thereby leading to sub-optimal tumor response, as indicated by its negative correlation with TGI (**Fig. 3A**, inset). Similarly, EC_50,*AM*_ negatively correlates with tumor response, which indicates the influence of the potency of anti-miR-155 in modulating the degradation of miR-155 to suppress tumor proliferation. Finally, *δ*_NP_ that represents the degradation rate of NPs can influence their systemic availability, thereby affecting the tumor exposure of anti-miR-155, leading to a negative correlation with tumor response (**Fig. 3A**, inset).

Thus, through sensitivity analyses we were able to identify the ten system parameters that significantly impact tumor response to anti-miR-155 therapy, which were then used to establish a dose-response relationship for anti-miR-155 and quantify the associated uncertainty in model predictions due to population-scale variability in parameter values. Importantly, these analyses also highlighted the control parameters of the system, e.g., the ones related to NP design (***Φ***_NP_, *δ*_NP_), which can be fine-tuned experimentally to achieve the desired treatment outcomes.

### Dose-response relationship

To investigate the effect of treatment regimen, i.e., dose and treatment frequency, on the efficacy of anti-miR-155 therapy, we used the clinical model to simulate treatments with varying doses of anti-miR-155: once a week (QW), once every two weeks (Q2W), and once every three weeks (Q3W) treatment schedules for a duration of ∼six months (see **Methods** for details). We used two metrics to evaluate treatment effect: 1) TGI and 2) change in tumor diameter from baseline, as defined by RECIST 1.1. While the allometrically scaled human equivalent dose for anti-miR-155 was 0.026 mg/kg (marked by yellow diamond on the *x*-axis, **Fig. 3B,C**), we studied the effect of dose on TGI across four orders of magnitude, ranging from 10^−3^–10 mg/kg, and fitted the Hill equation (**Eq. 1**) to quantitatively characterize dose-response relationships to enable appropriate dose selection in subsequent translational analysis. To quantify the uncertainty in predictions, 1,000 distinct combinations of the most sensitive model parameters (identified from GSA) were generated through Latin Hypercube sampling within a ±10% range of their baseline values, and then tumor responses were obtained for each dose across these 1,000 unique scenarios to determine 90% prediction intervals for the dose-response relationships (see **Methods**).

As shown in **Fig. 3B**, anti-miR-155 exhibited an average maximal efficacy (*E*_max_) of 66.7±0.2% TGI across the three treatment schedules. The dose-response curves indicated that the drug’s effect saturated for the QW scenario at a slightly lower dose compared to the Q2W and Q3W regimens. Thus, across all treatment schedules, the saturation dose (approximated as EC_99_ or dose leading to 99% of *E*_max_) fell within the range of 1–24 mg/kg (**Fig. 3B, inset**). This range indicates the dose at which almost all molecular targets of anti-miR-155 were fully occupied. Moreover, the Hill coefficient (*n*), a measure of the curve’s steepness or slope, was relatively consistent across the different treatment schedules, averaging 0.74±0.04. The Hill coefficient quantifies the degree of cooperativity in the binding of the drug to its molecular targets, and a value around 1 suggests limited cooperativity. Of note, the drug’s potency, represented by the EC_50_, exhibited a rightward shift with decreasing treatment frequency (i.e., 0.003 mg/kg for QW, 0.013 mg/kg for Q2W, and 0.033 mg/kg for Q3W). This shift can be attributed to changes in the pharmacokinetics of the drug due to delayed injections, indicating that reduced exposure of the tumor to anti-miR-155 under less frequent injections necessitates higher doses to achieve the same therapeutic effect (compare **Figs. 2H-L, S7, S8**). This finding underscores the importance of treatment schedule considerations in optimizing anti-miR-155 efficacy.

We also evaluated the dose-response relationship in terms of the RECIST 1.1-derived measure of treatment response, i.e., % change in tumor diameter from baseline or the smallest diameter, whichever is smaller following treatment. This relation was characterized by a modified form of Hill equation (**Eq. 2**). As shown in **Fig. 3C**, with the increase in dose, the % change decreased significantly from 268.9±10% (i.e., *E*_min_) to 21.5±1.8% (i.e., *E*_max_) across the various treatment schedules. Thus, the average response across all studied dosages consistently fell within the ‘progressive disease’ category (i.e., above the dotted dark-red line in **Fig. 3C**). This indicates that regardless of the dose and treatment frequency, the tumor diameter increased from baseline or its smallest size by at least 20%, since the initiation of treatment. The other parameters that were characterized, including Hill coefficient *n* and EC_50_ showed similar trends as the previous results (**Fig. 3C, inset**). Thus, having established dose-response relationships, we turned our attention to the application of these insights in evaluating the clinical efficacy of anti-miR-155 in a virtual patient cohort. This next phase allowed us to extrapolate our findings to a more realistic clinical context and assess the implications for patient outcomes.

### Clinical efficacy of anti-miR-155 in a virtual patient cohort

To assess the clinical efficacy of anti-miR-155 therapy, we employed a virtual patient cohort representing stage IA, mirroring the characteristics of early-stage, localized NSCLC (1−3 cm in diameter) per the TNM staging system (see **Methods** for details of virtual patient generation; **Fig. S9**) (23). This cohort, comprising 1,000 virtual patients, underwent treatment with various doses of anti-miR-155 administered once every three weeks (Q3W) for ∼six months, equivalent to nine treatment cycles. The selected doses were strategically chosen from our established dose-response Hill equation for the Q3W scenario (**Fig. 3B**), encompassing a spectrum of doses, including the allometrically scaled dose, EC_50_, EC_60_, EC_70_, EC_80_, EC_90_, EC_95_, and EC_99_. This approach allowed us to explore a wide range of doses while aligning with the Q3W clinical regimen commonly used for standard-of-care drugs for NSCLC, such as pembrolizumab, atezolizumab, and cisplatin. More frequent dosing schedules, e.g., Q2W or QW, may offer continuous drug exposure, but can present challenges in terms of patient compliance, resource allocation, and potential for increased treatment-related adverse events. To evaluate the clinical efficacy of anti-miR-155, the model-predicted tumor trajectory for every virtual patient was evaluated to determine using RECIST 1.1 the progression-free survival (PFS) as the clinical endpoint of interest.

As depicted in **Fig. 4A**, the average tumor growth trajectory for the virtual patient cohort exhibited slower progression from the initiation of treatment (at time zero) compared to the control scenario, with the degree of suppression positively correlating with the administered dose. To quantify the suppression in tumor growth, we assessed the time to progression (TTP), defined as the duration from the initiation of treatment when the tumor diameter exceeded 20%, with a minimum increase of 5 mm, following RECIST 1.1 (see **Methods**). The dotted dark-red line represents the average threshold used for the virtual cohort to distinguish between ‘progressive disease’ and ‘stable disease’ (**Fig. 4A**). Increasing the dose resulted in a longer time to cross this threshold of TTP, indicating slower tumor progression. The distribution of TTP across the virtual cohort for the control and treatment groups revealed a significant increase in the mean values with increasing dose (**Fig. 4B**; *p*-value < 0.0001, one-way ANOVA). Specifically, the mean TTP for the control scenario was 1.49±0.43 months, which increased to 2.48±1.17 months for the allometrically scaled dose (0.026 mg/kg) and 6.87±0.96 months for the EC_99_ dose (23.37 mg/kg). As per Tukey’s test, all pairs were significantly different, except the allometrically scaled dose (Allo.) and the EC_50_ group.

**Figure 4.**
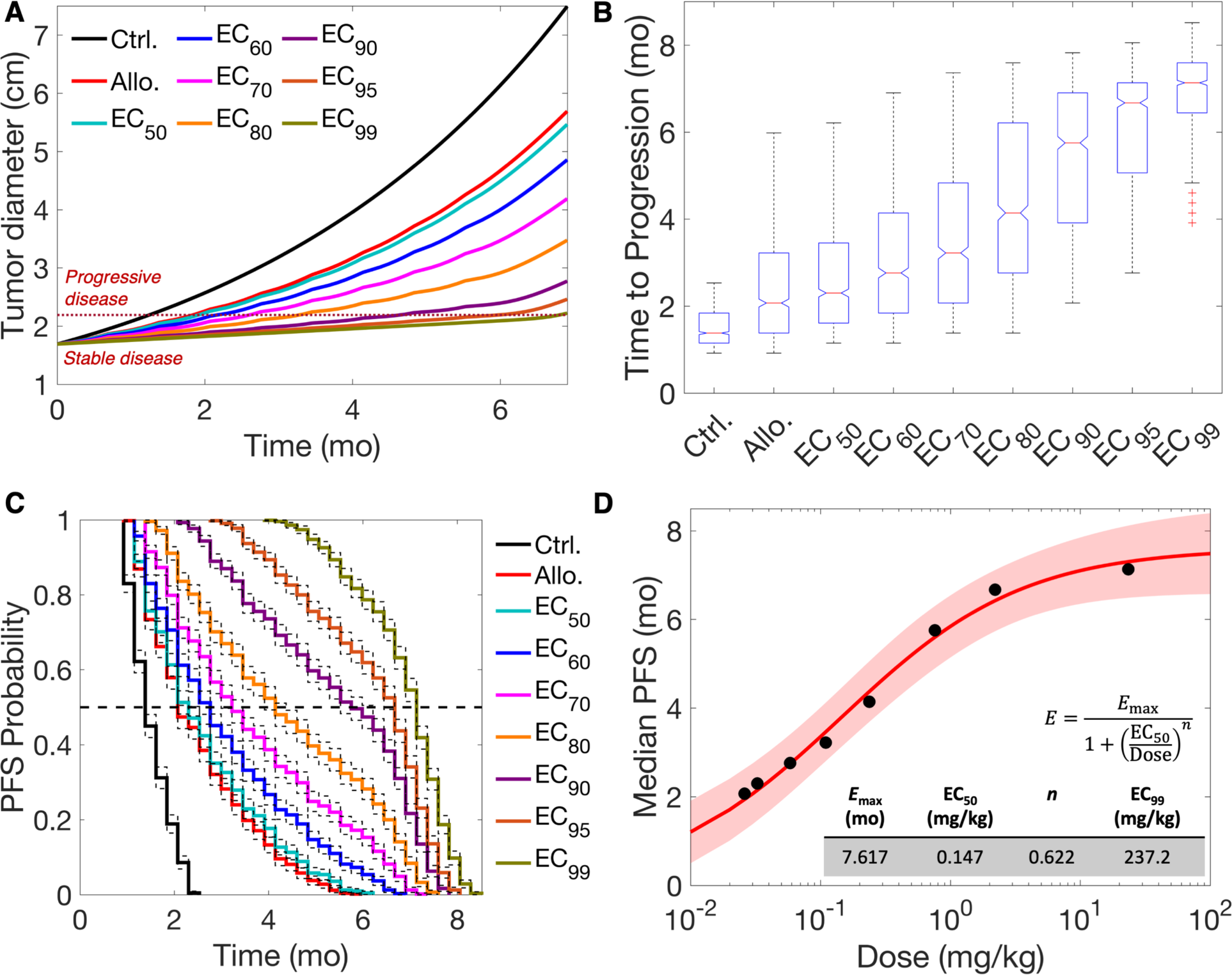
Clinical efficacy of anti-miR-155 in a virtual patient cohort. 1,000 virtual patients were generated and based on the dose-response curve for Q3W treatment scenario, tumor response to the allometrically scaled dose (Allo.) and other EC*i* doses was predicted for the virtual cohort. **A)** Average tumor growth kinetics across the virtual patients for different doses is shown: these formed the basis to characterize response using RECIST 1.1. Time to progression (TTP) marks the event when tumor diameter crosses the threshold of 20% increase in value from baseline (with a minimum of 0.5 cm increase), leading to the response being characterized as ‘progressive disease’. **B)** Distribution of TTP across virtual patients is shown, which shows a significant increase with increasing dose. **C)** Kaplan-Meier analysis using TTP was performed for the various doses to obtain progression-free survival (PFS) probability over time, which showed an increase in median PFS with increasing dose. **D)** Hill equation was used to characterize the relation between median PFS and dose. Note: Median PFS represents the time when 50% of patients experienced progressive disease, i.e., PFS probability becomes 0.5.

Next, we utilized the TTP values for each group to conduct Kaplan-Meier analysis for determining the PFS probability, a crucial clinical endpoint in oncology indicating the probability of remaining free from disease progression at a given time. As shown in **Fig. 4C**, the PFS curve for the control group displayed a rapid decline in PFS probability over time, indicative of rapid transition to progressive disease. In contrast, corresponding to the dose, the PFS curves for the treatment groups showed slower decline in PFS probability, indicative of prolonged periods of disease stability and thus slower progression. As reported in **Table 1**, the median PFS, representing the time when 50% of patients experienced progressive disease, i.e., PFS probability became 0.5, consistently increased with higher doses. Further, we measured the hazard ratios for assessing the risk of disease progression in treatment groups compared to the control group, and as shown in **Table 1**, the hazard ratio was consistently less than 1. The consistently low hazard ratios across treatment groups suggest a substantial reduction in the risk of disease progression associated with anti-miR-155 treatment. Further, the hazard ratio exhibited a dose-dependent reduction, demonstrating a clear dose-response relationship where higher doses of anti-miR-155 were associated with a greater reduction in the risk of disease progression. Finally, we characterized the relationship between median PFS and dose by fitting the Hill equation, and as shown in **Fig. 4D**, the median PFS for anti-miR-155 saturated at ∼7.6 months (95% CI, 6.8– 8.5 months), corresponding to a dose (approximated as EC_99_) of 237.2 mg/kg. This indicates that a very high dose is required for anti-miR-155 monotherapy to exhibit its maximal therapeutic efficacy in the Q3W scenario. This may have important implications for the translation of this strategy, given the adverse events as observed in phase 1 clinical trials of miR-34a mimic, where the recommended phase 2 dose did not exceed ∼2.5 mg/kg for any solid tumor (31). However, in the case of phase 1 trials of cobomarsen, an inhibitor of miR-155, for cutaneous T cell lymphoma, no adverse events were reported until a dose of ∼12.9 mg/kg (18). Given the disparity in the literature regarding the maximum tolerated dose of miRNAs or their inhibitors, we selected a more conservative dose of 2.5 mg/kg as the maximum tolerated dose of anti-miR-155 for subsequent analysis. As per the Hill equation, this dose corresponds to EC_85_ and will lead to a median PFS of 6.5 months, compared to the 12.9 mg/kg dose that yields a median PFS of 7.1 months with a five-fold increase in dose.

**Table 1.** Median PFS and hazard ratio for disease progression.

| Dose | Ctrl. | Allo. | EC<sub>50</sub> | EC<sub>60</sub> | EC<sub>70</sub> | EC<sub>80</sub> | EC<sub>90</sub> | EC<sub>95</sub> | EC<sub>99</sub> |
| --- | --- | --- | --- | --- | --- | --- | --- | --- | --- |
| Median PFS (months) | 1.4 | 2.1 | 2.3 | 2.8 | 3.2 | 4.1 | 5.8 | 6.7 | 7.1 |
| Hazard ratio | – | 0.31 | 0.28 | 0.2 | 0.13 | 0.07 | 0.01 | 7e-9 | 7e-9 |

Abbreviations: Ctrl.- control, Allo.- allometrically scaled dose.

Of note, none of the doses led to a partial or complete response as per RECIST 1.1 (**Fig. 4A**). This observation, combined with the need for high doses to achieve maximal therapeutic efficacy, underscores the challenges of achieving significant clinical responses with anti-miR-155 monotherapy. Based on these findings, we sought to determine whether combinations of anti-miR-155 with standard-of-care drugs could act synergistically to address the limitations of anti-miR-155 monotherapy and potentially improve outcomes.

### Drug combination studies and synergy evaluation

To facilitate drug combination studies involving anti-miR-155 and standard-of-care drugs for NSCLC, we first assessed the ability of the allometrically scaled model (for humans) to predict clinical endpoints for standard-of-care drugs. Due to potential discrepancies in interspecies scaling of drug-specific model parameters, the initial predictions of PFS probability required adjustments to align with clinical observations for the selected standard-of-care drugs (see **Supplementary Results S1**).

Following the fine-tuning of model parameters to improve predictions for standard-of-care drugs, we performed drug combination studies, exploring two- and three-drug combinations of anti-miR-155 with standard-of-care drugs across a range of doses (see **Methods** for details). These combinations included anti-miR-155 in non-constant dose ratios with 1) pembrolizumab, 2) atezolizumab, 3) cisplatin, 4) cisplatin and pembrolizumab, and 5) cisplatin and atezolizumab. Treatment with the combination therapies was simulated in our virtual patient cohort (*N* = 1,000) over a six-month period using a Q3W regimen, equivalent to nine treatment cycles in patients. Note that in the case of cisplatin, we administered only six cycles to reflect standard clinical practice (32). For every drug combination, median PFS was calculated through Kaplan-Meier analysis based on TTP in virtual patients. Additionally, the corresponding TGIs at the end of treatment were calculated to evaluate synergy using the Chou-Talalay method (33). We also calculated the median PFS corresponding to the monotherapy arms as a reference. For the four monotherapies, median PFS (i.e., efficacy) was less than 7.5 months. Specifically, treatment with cisplatin, anti-miR-155, atezolizumab, and pembrolizumab at their highest doses (clinical doses for standard-of-care drugs and 2.5 mg/kg for anti-miR-155; **Table 2**) resulted in median PFS values of approximately 2.5, 6.5, 6.7, and 7.4 months, respectively (**Fig. S12**). However, when drugs were used in two- and three-drug combinations, a consistent pattern of improved efficacy was observed (**Fig. 5A-E**).

**Figure 5.**
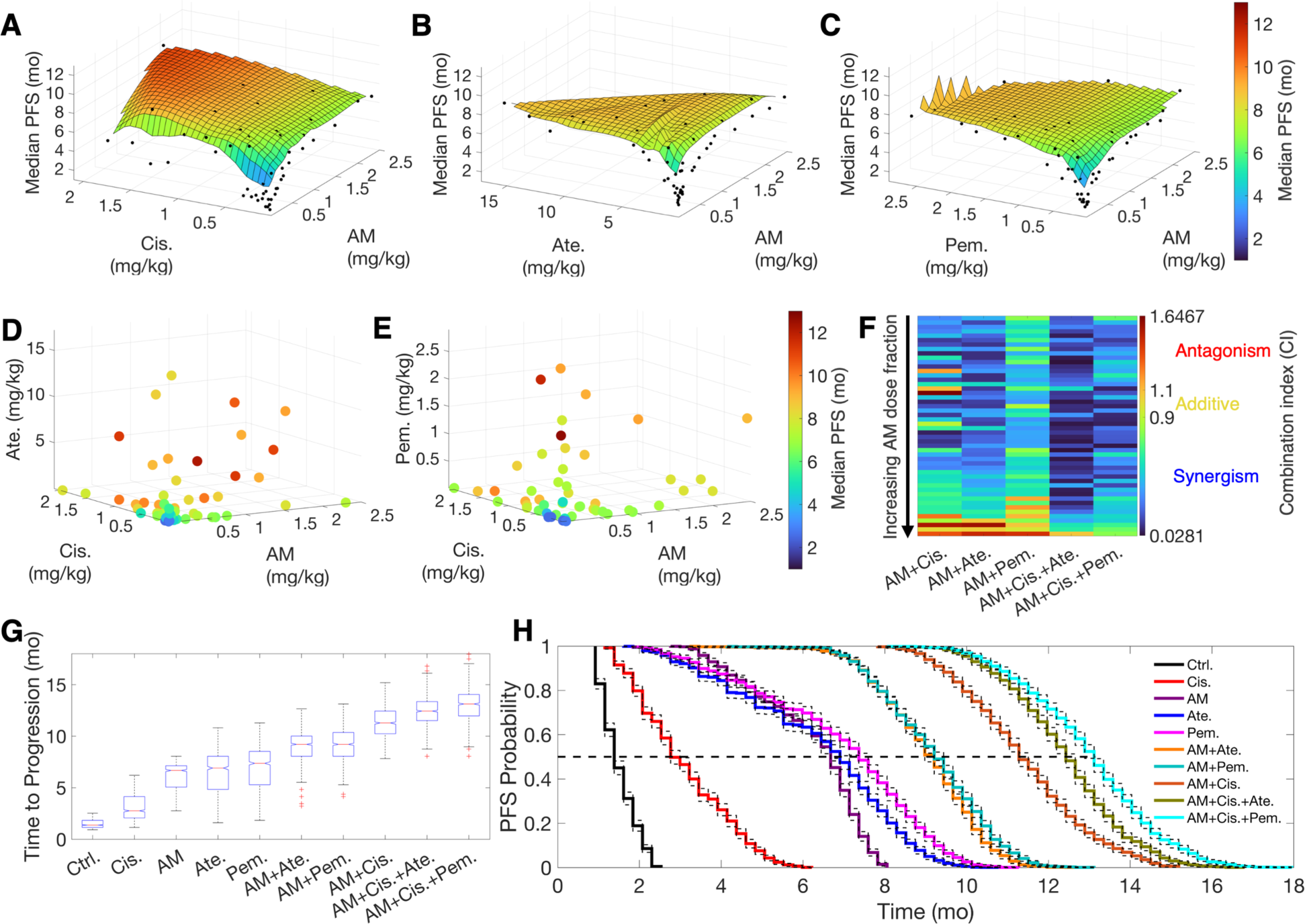
Drug combination studies and synergy evaluation. 50 samples obtained using Latin hypercube sampling for two- and three-drug combinations each were investigated for predictions of median PFS under the Q3W regimen. 3D and 4D plots show median PFS corresponding to combinations of anti-miR-155 (AM) with **A**) cisplatin (Cis.), **B)** atezolizumab (Ate.), **C)** pembrolizumab (Pem.), **D)** cisplatin and atezolizumab, and **E)** cisplatin and pembrolizumab. **F)** Combination indices (CI) for the various combinations are shown, with the combinations aligned in an ascending order of anti-miR-155 dose fractions. Note that CI < 0.9 denotes synergism, 0.9 < CI < 1.1 denotes additive combinations, and CI > 1.1 indicates antagonism. **G)** Model-predicted time to progression and **H)** PFS for various monotherapies and strongly synergistic combination therapies following Q3W regimen.

**Table 2.** Median PFS corresponding to monotherapies and strongly synergistic combination therapies. All drugs were injected Q3W for nine treatment cycles, except cisplatin, which was given for six cycles only. Note, standard-of-care monotherapies were administered at their clinically prescribed doses, with calculations based on assumptions of body weight of 70 kg and body surface area of 1.9 m^2^.

| Drug/s | Dose/s (mg/kg) | Median PFS (months) |
| --- | --- | --- |
| Control | – | 1.4 |
| Cisplatin (Cis.) | 2.04 (≡ 75 mg/m<sup>2</sup>) | 2.8 |
| anti-miR-155 (AM) | 2.5 | 6.7 |
| Atezolizumab (Ate.) | 17.14 (≡ 1200 mg) | 6.9 |
| Pembrolizumab (Pem.) | 2.86 (≡ 200 mg) | 7.4 |
| AM+Ate. | 0.21 + 15.94 | 9.2 |
| AM+Pem. | 1.31 + 2.04 | 9.2 |
| AM+Cis. | 1.62 + 1.89 | 11.3 |
| AM+Cis.+Ate. | 1.34 + 1.36 + 3.12 | 12.4 |
| AM+Cis.+Pem. | 1.24 + 1.75 + 0.88 | 13.1 |

Abbreviations: AM- anti-miR-155, Cis.- Cisplatin, Ate.- Atezolizumab, Pem.- Pembrolizumab.

The combination of anti-miR-155 with cisplatin demonstrated a remarkable dose-dependent enhancement in efficacy, as illustrated in **Fig. 5A**. In this combination, median PFS reached up to 11 months at doses that were below the clinical dose of cisplatin and the selected upper limit for anti-miR-155 (1.89 mg/kg and 1.62 mg/kg, respectively; **Table 2**). The concurrent administration of anti-miR-155 effectively reduced cisplatin resistance attributed to the blockade of miR-155/TP53-negative feedback (3), substantially improving cisplatin efficacy at reduced doses. This observation indicates synergistic activity, supported by the combination index (CI) values obtained using the Chou-Talalay method. As shown in **Fig. 5F** and **Supplementary Table S4**, 86% of the dose combinations between anti-miR-155 and cisplatin exhibited synergism (CI < 0.9). Further, as described in **Table 3**, combinations that simultaneously adhered to a set of conditions achieved strong synergism (i.e., CI < 0.3). These conditions included a cisplatin dose greater than 0.25 mg/kg, a dose fraction of anti-miR-155 exceeding 1.4%, and a total combination dose surpassing 0.55 mg/kg. We, however, also observed antagonistic combinations, primarily when using a cisplatin dose of less than 0.07 mg/kg combined with either a low dose of anti-miR-155 (< 0.013 mg/kg) or a high dose of anti-miR-155 (> 1.4 mg/kg). This antagonism is attributed to the dual molecular effects of miR-155, which induce cisplatin resistance in cancer cells while suppressing PD-L1 expression in cancer cells and TAMs to favor tumor immunosurveillance. Higher doses of anti-miR-155 can diminish the benefits of cisplatin sensitization by reinforcing tumor immunosurveillance blockade, ultimately leading to poorer outcomes. Similarly, when both cisplatin and anti-miR-155 are administered at low doses, the same antagonistic effect may come into play due to limited cisplatin sensitization.

**Table 3.** Dose conditions associated with strong synergy (CI < 0.3) and antagonism (CI > 1.1) in various drug combinations.

| Combinations | Conditions for strong synergy | Conditions for antagonism |
| --- | --- | --- |
| AM+Cis. | Dose of Cis. > 0.25 mg/kg AND Dose fraction* of AM > 1.4% AND Total dose > 0.55 mg/kg | Dose of Cis. < 0.07 mg/kg AND (Dose of AM < 0.013 mg/kg OR Dose of AM > 1.4 mg/kg) |
| AM+Ate. | Dose of Ate. > 0.22 mg/kg AND Dose of AM > 0.026 mg/kg AND Dose fraction of AM < 42% | Dose of AM > 1.9 mg/kg AND Dose fraction of Ate. < 1.1% |
| AM+Pem. | All combinations led to CI > 0.3 | Dose of AM > 1.32 mg/kg AND Dose fraction of Pem. < 7.1% |
| AM+Cis.+Ate. | Dose fraction of AM < 60% AND Combined dose of AM+Cis. > 0.05 mg/kg AND Total dose > 0.2 mg/kg | All combinations led to CI < 1.1 |
| AM+Cis.+Pem. | Dose of Cis. > 0.065 mg/kg AND Dose of AM < 1.25 mg/kg AND Dose fraction of Pem. < 74% AND Total Dose > 0.41 mg/kg | All combinations led to CI < 1.1 |

\*Dose fraction is the fraction of a given drug's dose in the total dose of all drugs in the combination. Abbreviations: AM- anti-miR-155, Cis.- Cisplatin, Ate.- Atezolizumab, Pem.- Pembrolizumab.

We next evaluated two-drug combinations of anti-miR-155 with the ICIs atezolizumab and pembrolizumab. These combinations are of particular interest due to the increased expression of PD-L1 on tumor cells and TAMs upon treatment with anti-miR-155 (**Fig. 2K**), which can potentially augment the efficacy of co-administered ICI, particularly anti-PD-L1 antibodies, due to the increased availability of drug binding sites (34,35). The two combinations exhibited dose-dependent improvement in efficacy, with the highest observed median PFS value of ∼9 months in both cases (**Fig. 5B,C**), compared to the values of 6.7 and 7.4 months for atezolizumab and pembrolizumab monotherapies, respectively (**Fig. S12**). Notably, as reported in **Table 2**, the improved efficacy with combination therapies was attained at doses lower than the clinical dose of atezolizumab (∼16 mg/kg) and pembrolizumab (∼2 mg/kg), as well as the chosen upper limit of anti-miR-155 (0.21 and 1.31 mg/kg for the atezolizumab and pembrolizumab combination, respectively). This indicates synergy, as 92% and 86% of the studied combinations of anti-miR-155 with atezolizumab and pembrolizumab exhibited synergism (CI < 0.9), respectively (**Fig. 5F** and **Supplementary Tables S5, S6**). Furthermore, a strong synergy (CI < 0.3) was observed with anti-miR-155 combined with atezolizumab but not with pembrolizumab. For the former, a dose of atezolizumab greater than 0.22 mg/kg, combined with a dose of anti-miR-155 greater than 0.026 mg/kg but comprising a dose fraction of less than 42%, led to strong synergy (**Table 3**). Antagonistic combinations (CI > 1.1) were also observed under specific dose conditions (**Table 3**). Combinations of anti-miR-155 with atezolizumab, involving anti-miR-155 doses exceeding 1.9 mg/kg and atezolizumab dose fractions below 1.1%, as well as combinations of anti-miR-155 with pembrolizumab, comprising anti-miR-155 doses greater than 1.32 mg/kg but with pembrolizumab dose fractions below 7.1% exhibited antagonism. These findings are consistent with anti-miR-155’s stimulatory effect on PD-L1 expression in tumor cells and TAMs. Higher doses of anti-miR-155 elevate PD-L1 expression to high levels, overwhelming the low doses of atezolizumab or pembrolizumab to effectively block PD-L1 and PD-1 interactions.

Since the two-drug combinations of anti-miR-155 with standard-of-care therapies showed synergy and increased median PFS over monotherapies, we extended our investigation to three-drug combinations to explore further enhancement in efficacy. Combinations of anti-miR-155 and cisplatin with atezolizumab (**Fig. 5D**) or pembrolizumab (**Fig. 5E**) exhibited a further increase in efficacy, leading to a median PFS of up to 12.2 and 12.7 months, respectively, thereby surpassing the efficacy of the anti-miR-155 and cisplatin two-drug combination of 11 months. Notably, the three-drug combinations achieved higher efficacy at further reduced doses (**Table 2**). In the best-performing combination with atezolizumab, doses of anti-miR-155 and cisplatin were 82% and 71%, respectively, of the doses used in the most efficacious two-drug combination. Similarly, in the best-performing combination with pembrolizumab, doses of anti-miR-155 and cisplatin were 77% and 93% of the doses used in the most efficacious two-drug combination. Further corroborated by the Chou-Talalay analysis, 98% and 100% of the three-drug combinations with atezolizumab and pembrolizumab, respectively, were synergistic (CI < 0.9; **Fig. 5F** and **Supplementary Tables S7, S8**); 80% and 46% of the combinations involving atezolizumab and pembrolizumab, respectively, showed strong synergy (CI < 0.3), while none of the combinations were antagonistic in these scenarios. The conditions necessary for strong synergy are reported in **Table 3**.

Finally, as representative examples, we simulated a six-month Q3W treatment using monotherapies at clinical doses and the upper limit of anti-miR-155, strongly synergistic (CI < 0.3) two-drug combinations (at doses leading to the highest median PFS), and strongly synergistic three-drug combinations (also at doses leading to the highest median PFS) in our virtual patient cohort (*N* = 1,000). This simulation allowed us to obtain the distribution of TTP and generate PFS curves through Kaplan-Meier analysis for comparison between groups. In showcasing these best results from various mono- and combination therapies, we provide a comprehensive view of the treatments that demonstrated the greatest potential.

As depicted in **Fig. 5G**, the boxplot illustrates the distribution of TTP for different treatment groups versus control. A significant increase in the mean value of TTP is evident as we transition from the control group to the three-drug combination group (p-value < 0.0001; One-way ANOVA). The control group exhibited the shortest median TTP (1.4 months), followed by the monotherapies and two-drug combinations, with the three-drug combinations showcasing the longest median TTP (12.4 and 13.1 months with atezolizumab and pembrolizumab, respectively). Of note, the two-drug combination of anti-miR-155 and cisplatin was comparable to the three-drug combinations, yielding a median TTP of 11.3 months. This finding highlights the enhanced therapeutic efficacy of the two-drug combination, potentially offering an effective solution when three-drug combinations are not feasible. Such practicality may stem from the simplicity of a two-drug regimen, reduced potential for toxicity, and cost-effectiveness. Lastly, the observed trend in TTP aligns with the values of median PFS reported in **Table 2**, as obtained from the corresponding PFS probability curves shown in **Fig. 5H**.

## Conclusions

In this study, we conducted a comprehensive investigation of the translational potential of anti-miR-155 therapy for NSCLC. We began by developing a multiscale mechanistic model that accounts for the complex biophysical interactions, physiological processes, and transport phenomena associated with the delivery of systemically administered NP-loaded anti-miR-155 and its effects on tumor growth dynamics. This model was rigorously calibrated with *in vivo* datasets from NSCLC-bearing mice, followed by interspecies scaling to humans through allometric scaling techniques, providing a solid foundation for subsequent translational analyses. Our study focused on addressing two crucial aspects: (1) the translational efficacy of anti-miR-155 monotherapy for NSCLC and (2) the potential for enhancing treatment outcomes through combination therapies involving anti-miR-155 and standard-of-care drugs in NSCLC.

Our modeling and simulations demonstrated the efficacy of anti-miR-155 monotherapy in a virtual patient cohort, showing dose-dependent suppression of tumor growth that led to enhanced efficacy, as defined by the clinically relevant endpoints of median PFS and TTP. At a dose of 2.5 mg/kg given once in three weeks over six months, anti-miR-155 monotherapy exhibited a median PFS of ∼6.7 months, compared to the control and cisplatin groups with median PFS of 1.4 and 2.8 months, respectively.

To further enhance the efficacy of anti-miR-155, we explored the possibility of combining it with standard-of-care drugs. We identified strongly synergistic two- and three-drug combinations involving anti-miR-155, cisplatin, atezolizumab, and pembrolizumab that showed remarkable efficacy at reduced doses. A particularly compelling combination of anti-miR-155, cisplatin, and pembrolizumab delivered a median PFS of 13.1 months, while substituting atezolizumab produced a comparable median PFS of 12.4 months. Notably, a simplified combination involving only anti-miR-155 and cisplatin demonstrated a median PFS of 11.3 months, potentially offering practical advantages due to simplicity and cost-effectiveness when compared to the more complex three-drug combinations. This combination takes advantage of the anti-miR-155-mediated blockade of the miR-155/TP53-negative feedback mechanism of cisplatin resistance in NSCLC. Our analyses also provided critical insights into suboptimal combination doses, underscoring the need for careful dose regimen design to prevent potential antagonistic effects that could compromise treatment outcomes.

This modeling study has several limitations, including simplifications in our mathematical model, such as the absence of spatial heterogeneity in the tumor compartment and the assumption of uniform distribution of NPs. Also, while the approach of allometric scaling to obtain a clinically relevant model offers a bridge between preclinical and clinical settings, it can introduce uncertainties in model predictions. Furthermore, our analysis lacked the genetic and PD-L1 expression heterogeneity of NSCLC tumors that is known to influence outcomes, and thus warrants future investigations for more accurate predictions to support treatment personalization. Addressing these limitations is crucial for advancing the clinical translation of anti-miR-155 therapy, and further research will aim to incorporate these aspects for a more comprehensive understanding and application in NSCLC treatment.

In summary, our study provides a strong quantitative rationale for considering anti-miR-155 therapy in the treatment of NSCLC. The findings of drug combination studies underscore the potential for synergy in combinations but also points to potential for antagonism. The multiscale mechanistic model, calibrated with *in vivo* data, can serve as a valuable tool for predicting clinical treatment endpoints and guiding personalized treatment strategies for systemic therapies. These results offer a foundation for future clinical trials of anti-miR-155, bringing us closer to more effective and safer therapeutic approaches for NSCLC patients.

## Methods

### Mathematical model development

Extending our previous modeling framework (17,19,20), the mechanistic model was formulated as a system of ordinary differential equations (ODEs) that incorporate the relevant biological processes and interactions to describe the temporal evolution of key physiological, pathological, and pharmacological variables. As shown in **Fig. 1**, while the plasma compartment characterizes the systemic pharmacokinetics of anti-miR-155-loaded NPs and standard-of-care drugs, the tumoral compartment encompasses tumor-immune interactions, intratumoral transport of drugs, and tumor growth dynamics under control or treatment conditions, thereby leading to a multiscale model of tumor response to systemic therapies that was adapted for NSCLC.

In NSCLC, miR-155 is known to be overexpressed not only in tumor cells (3) but also in TAMs (5), and this phenomenon creates a secondary source of miR-155 for tumor cells through TAM-secreted exosomes. Consequently, miR-155 overexpression in cancer cells downregulates tumor suppressor genes (36), which drives tumor progression. Importantly, miR-155 also plays a dual role in the therapeutic response, i.e., inducing cisplatin resistance through a miR-155/TP53-negative feedback mechanism (3) and suppressing PD-L1 expression in NSCLC (7,8). While PD-L1 suppression may promote tumor immunosurveillance by CD8+ T cells, it can also induce resistance to anti-PD-L1 and anti-PD-1 monotherapies (34,35). Our model captures these intricate effects of miR-155 overexpression in NSCLC, offering insights into innovative treatment strategies, particularly the potential of miR-155 antagonists (i.e., anti-miR-155) as a combination therapy to enhance the efficacy of standard-of-care drugs, including cisplatin, pembrolizumab, and atezolizumab, in NSCLC.

In translational modeling, it is crucial to comprehensively capture the multiscale transport phenomena involved in drug delivery to the tumor. This allows us to account for the inter-patient variability in relevant physiological processes and tumor characteristics to provide a more accurate representation of how anti-miR-155-loaded NPs and free drugs navigate the intricate landscape of the tumor microenvironment. After injection into the bloodstream (i.e., plasma compartment), the anti-miR-155-loaded NPs and free drugs, collectively referred to here as *agents*, are influenced by pharmacokinetic (PK) processes that determine their systemic concentration kinetics. From the bloodstream, these agents navigate their way into the tumor interstitium through a permeation-limited delivery process across tumor microvasculature. Once inside the tumor interstitium, where advection is limited due to high interstitial fluid pressure (37), the agents rely on diffusion to reach the cell membranes of their target cells. Depending on the type of agent, specific biophysical processes come into play to facilitate delivery to the target site and trigger the pharmacodynamic effects. For example, NPs are internalized into cancer cells through endocytosis, releasing anti-miR-155 into the cytosol. In contrast, chemotherapeutics diffuse directly into the cell cytosol, whereas antibodies bind to specific cell surface receptors and ligands, such as PD-1 and PD-L1 in the present work. Following successful delivery to the target site, the pharmacodynamic component of the model becomes active, where anti-miR-155 promotes the degradation of miR-155. Cisplatin, the reference chemotherapeutic, induces apoptotic cell death, while anti-PD-L1 and anti-PD-1 antibodies inhibit the tumor protective effects of PD-L1 and PD-1 immune checkpoints, respectively, leading to enhanced T cell-mediated tumor death.

In **Supplementary Methods S1**, we describe in detail the ODEs that capture the intricate dynamics of the modeled system (**Equations S1–S29**). Note that the equations are based on the law of mass action and obey the conservation of mass.

### Model parameterization and calibration

The model was first parameterized with physiological parameter values either obtained directly from literature for mice or estimated through nonlinear least squares fitting of the model to literature-derived *in vivo* tumor growth kinetics data. Specifically, the system of ODEs detailed in **Equations S1-S29** was solved numerically as an initial value problem (initial conditions given in **Supplementary Table S3**) in MATLAB R2022b using the built-in function *ode15s* to simulate the exact treatment protocols as described in the experimental studies. The resulting numerical solution was then fit to the *in vivo* data using the built-in function *lsqcurvefit* to minimize the error between observed data and model predictions. The selected datasets for model fitting encompass mice engrafted with patient-derived xenografts or cell line xenografts of NSCLC that were treated with anti-miR-155-loaded liposomes (3), cisplatin (3), combination of anti-miR-155 and cisplatin (3), atezolizumab (21), or pembrolizumab (22). Pearson correlation analysis was performed to evaluate the goodness of fit between the model and the *in vivo* data. Model parameters, including those estimated through fitting, are detailed in **Supplementary Tables S1** and **S2**.

### Interspecies scaling

To extrapolate our model from mice to humans, the parameter values for mice were either substituted with population averages for humans, or allometrically scaled as a function of body weights. Thus, the value of parameter *i* for humans 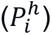 was determined by: 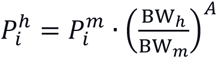, where 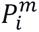 is the value of parameter *i* for mice, BW*_h_* and BW*_m_* are the assumed body weights for humans (BW*_h_* = 70 kg) and mice (BW*_m_* = 0.02 kg), respectively, and ***A*** is the allometric exponent. The choice of allometric exponent ***A*** varies depending on the subset of parameters being scaled. For instance, a value of ***A*** = −0.25 is applied to rate constants (38), a value of ***A*** = 0.75 is utilized for clearance (39), a value of ***A*** = 1 is applied to the volume of distribution (40), while a value of ***A*** = −0.33 is associated with dose scaling (41). **Supplementary Tables S1** and **S2** report values of the model parameters for both mice and humans.

### Treatment response evaluation

The model was used to study the effect of anti-miR-155 therapy, alone or in combination with standard-of-care drugs, on tumor growth dynamics. To assess treatment response, we employed two complementary metrics: 1) tumor growth inhibition (TGI) and 2) change in tumor diameter from baseline, as defined by RECIST 1.1 guidelines (24). Note that baseline here refers to the tumor size at the time of treatment initiation or the smallest tumor size recorded since the beginning of treatment. See **Supplementary Methods S2** for details.

### Parameter sensitivity analysis

To investigate the importance of the various model parameters in causing tumor shrinkage under treatment with anti-miR-155, we conducted a detailed parameter sensitivity analysis. We performed both local sensitivity analysis (LSA) and global sensitivity analysis (GSA) by perturbing specific parameters of interest. These parameters were systematically altered over a range of ±50% of their baseline values, and we measured their impact on tumor growth inhibition (TGI) under these modified conditions. See **Supplementary Methods S3** for details.

### Characterization of dose-response relationship

This analysis aimed to explore the impact of treatment regimen, i.e., dose and treatment frequency, on the efficacy of anti-miR-155 monotherapy. The various treatment regimens that were investigated included once a week (QW), once every two weeks (Q2W), and once every three weeks (Q3W) injections for time periods equivalent to 25, 13, and 9 treatment cycles, respectively; the total treatment duration was ∼six months. To comprehensively establish the dose-response relationship, 20 different doses of anti-miR-155 were examined between 10^−3^ to 10 mg/kg, sampled uniformly on log scale. Note that treatment for every scenario was initiated at 124 weeks post-tumor inception with a single cancer cell on day zero. The assessment of tumor response to every dose at the various treatment frequencies was based on two key metrics, defined previously: (1) TGI by the end of treatment and (2) change in tumor diameter from baseline as defined by RECIST 1.1.

To characterize the relationship between dose and TGI, we employed the Hill equation, a sigmoidal dose-response model, which is given as:

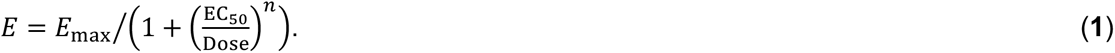

Here, *E* is the effect of anti-miR-155 measured as TGI; *E*_max_ is the maximal effect of the drug; EC_50_ is defined as half maximal effective dose (the dose that produces half of the maximal effect, i.e., *E*_max_/2) and is an indicator of drug potency; *n* is the Hill coefficient, which is a measure of the curve’s steepness or slope and quantifies the degree of cooperativity in the binding of the drug to its molecular target.

Note that the relationship between dose and RECIST 1.1-derived measure of treatment response was characterized by a modified form of Hill equation (42), given by:

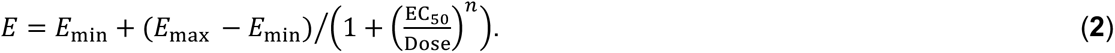

Here, *E*_min_ is the effect at zero dose.

To account for population-scale variability in treatment response or uncertainty in parameter estimates, 1,000 combinations of the most sensitive parameters identified from GSA were generated through Latin Hypercube sampling from ±10% range of their baseline values. By simulating tumor treatment at a given dose and frequency 1,000 times to test the various parameter combinations, we were able to obtain 90% prediction intervals to quantify uncertainty in dose-response relationships.

### Generation of virtual patient cohort

To evaluate the effects of drugs under clinically relevant scenarios, we generated a virtual patient cohort that can reasonably capture the biological and physiological variability of a patient population that had tumor characteristics representative of stage IA NSCLC, as per the TNM staging system (23). Specifically, our virtual cohort was intended to consist of tumors ranging in volumes from 0.5–10 cm^3^, such that the diameters of tumors varied between 1–2.68 cm, assuming spherical shape for tumors. To achieve this, we adapted the methodology of Allen et al. (43). See **Supplementary Methods S4** for details.

### Drug combination studies and synergy evaluation

The occurrence of synergy in drug combinations has the potential to enhance the efficacy of treatments and allow dose reduction for improved safety. To investigate the ability of anti-miR-155 to exhibit synergistic effects in combination with one or more standard-of-care drugs for NSCLC, we performed drug combination simulations, exploring two- and three-drug combinations of anti-miR-155 with standard-of-care drugs across a range of doses. These combinations included anti-miR-155 in non-constant dose ratios with 1) pembrolizumab, 2) atezolizumab, 3) cisplatin, 4) cisplatin and pembrolizumab, and 5) cisplatin and atezolizumab. To sample the doses effectively, we employed LHS, drawing fifty samples for each combination from a dose range spanning 0.01 mg/kg (the lower limit for all drugs) to the clinically prescribed dose for standard-of-care drugs. For anti-miR-155, the upper limit was set at 2.5 mg/kg based on our dose-response relationship analysis. Treatment with monotherapies and the various combination therapies were then simulated in our virtual patient cohort (*N* = 1,000) over a six-month period using a Q3W regimen, equivalent to nine treatment cycles. Note that, in the case of cisplatin, we administered only six cycles, aligning with clinical practice. Thus, for every sampled dose of monotherapies and combination therapies, median PFS was calculated from Kaplan-Meier analysis of the time to progression, as per RECIST 1.1, in virtual patients. Furthermore, the corresponding TGI at the end of treatment was calculated for combination therapies to evaluate synergy using the Chou-Talalay method (33). For this, a combination index (CI) was calculated using the open-source software COMPUSYN (https://www.combosyn.com/), such that CI < 1 is an indicator of synergism. The analysis report thus generated has been provided as **Appendix A** in supplemental material.

## Data Availability

Model code is available upon reasonable request to the authors.

## Acknowledgements

This research work was supported in part by the National Institutes of Health (NIH) Grants 1R01CA222007 (VC, BO, GAC, ZW), 1R01CA253865 (VC, BO, ZW), 1R01CA226537 (VC, ZW), 1R01AI165372 (ZW), R01DK132104 (ZW), 1R03EB033576 (PD), and P50CA217674 (KA). The funders had no role in study design, data collection and analysis, decision to publish, or preparation of the manuscript. PD also acknowledges Dr. Venkata K. Yellepeddi for helpful scientific discussions, Dr. Maria J. Peláez for assistance with data extraction, and Rachael E. Whitehead for her contributions to the model schematic.

## Author contributions

PD and ZW conceived the study. PD and JRR developed the model. PD and ZW designed the analyses. PD and VS performed the analyses with contributions from JC and CS. PD, VS, JDB, MC, DGD, AOK, CC, EJK, VC, BO, GAC, and ZW interpreted the biological and clinical relevance of the results. PD, VS, JRR, and ZW wrote the manuscript. JC, JDB, CS, MC, DGD, AOK, CC, EJK, VC, BO, and GAC edited the manuscript.

