## Supplementary Information for "Translational modeling-based evidence for enhanced efficacy of standard-of-care drugs in combination with anti-microRNA-155 in non-small-cell lung cancer"

#### Table of Contents

***Supplementary Methods*** ... **3**

S1. Model equations ... 3

S2. Treatment response evaluation ... 19

S3. Parameter sensitivity analysis ... 20

S4. Generation of virtual patient cohort ... 22

Table S1. List of biological parameters of the model. ... 23

Table S2. List of therapy related parameters of the model. ... 25

Table S3. Initial conditions of model variables. ... 27

***Supplementary Results*** ... **28**

S1. Clinical model calibration ... 28

Figure S1. Numerical solution of the model exhibiting kinetics of relevant variables under treatment with cisplatin. ... 30

Figure S2. Numerical solution of the model exhibiting kinetics of relevant variables under treatment with combination of cisplatin and nanoparticle-delivered anti-miR-155. ... 31

Figure S3. Numerical solution of the model exhibiting kinetics of relevant variables under treatment with atezolizumab. ... 32

Figure S4. Numerical solution of the model exhibiting kinetics of relevant variables under treatment with pembrolizumab. ... 33

Figure S5. Numerical solution of the model exhibiting kinetics of relevant variables under control conditions. ... 34

Figure S6. Pearson correlation of model fits to in vivo data shown in Figures 2E-G. ... 35

Figure S7. Numerical solution of the allometrically scaled model showing key system variables following treatment with 0.026 mg/kg anti-miR-155 once weekly. ... 36

Figure S8. Numerical solution of the allometrically scaled model showing key system variables following treatment with 0.26 mg/kg anti-miR-155 once in three weeks. ... 37

Figure S9. Virtual patient cohorts. ... 38

Figure S10. Global sensitivity analysis. ... 39

Figure S11. Clinical model calibration. ... 40

Figure S12. Predictions of median PFS for a virtual patient cohort under once in three weeks monotherapy regimen. ... 41

Table S4. Combination index (CI) values for combinations of anti-miR-155 and cisplatin. ... 42

Table S5. CI values for combinations of anti-miR-155 and atezolizumab. ... 43

Table S6. CI values for combinations of anti-miR-155 and pembrolizumab. ... 44

Table S7. CI values for combinations of anti-miR-155, cisplatin, and atezolizumab. ... 45

Table S8. CI values for combinations of anti-miR-155, cisplatin, and pembrolizumab. ... 46

***References*** ... **47**

#### Supplementary Methods

##### S1. Model equations

*The first seven equations characterize the kinetics of the expression of key molecular factors in the model.*

**Equation for miR-155 concentration in cancerous cells ($\boldsymbol{M_L(t)}$):**

$$\frac{dM_L(t)}{dt} = \overbrace{g_0^M}^{Production} + \overbrace{g_{\text{tran}} \cdot M_{\text{M}\phi}(t) \cdot \frac{\text{M}\phi(t)}{L(t)}}^{Exosomal\ tranfer} - \overbrace{\delta_M \cdot \left(1 + \frac{A_{\text{AM},M} \cdot \text{AM}_L(t)}{\text{EC}_{50,\text{AM}} + \text{AM}_L(t)}\right) \cdot M_L(t)}^{Degradation},$$

$$M_L(0) = M_0 \quad \textbf{(S1)}$$

We modeled the rate of change of miR-155 concentration inside cancer cells $(M_L(t))$ (units, pM) as the combination of three effects: production, exosomal transfer, and degradation. We assumed a constant rate of production of miR-155, characterized by the zero-order rate constant $g_0^M$. The second term represents the influx of miR-155 into cancer cells via miR-155-loaded exosomes secreted by TAMs. The rate of mass transfer in this process depends on the total mass of miR-155 inside TAMs $(M_{\text{M}\phi}(t) \cdot \text{M}\phi(\text{t}))$ and the transfer rate constant $g_{\text{tran}}$. $L(t)$ represents the total volume of cancer cells. The last term describes the degradation of miR-155. The degradation rate constant $\delta_M$ is regulated by the effects of the anti-miR-155 therapy, where the concentration of anti-miR-155 within cancer cells is denoted by $\text{AM}_L(t)$. These regulatory effects are modeled using Michaelis-Menten kinetics, where larger values of anti-miR-155 produce values closer to the dimensionless asymptote $A_{\text{AM},M}$, and thus an overall increment in the degradation rate of miR-155. $A_{\text{AM},M}$ is defined as an activation or stimulation factor that governs the degradation effects of anti-miR-155 on miR-155. The Michaelis constant is equivalent to the $EC_{50}$ of the anti-miR-155 therapy $(\text{EC}_{50,\text{AM}})$, with larger values of the $EC_{50}$ indicating a less effective therapy and vice-versa. Notably, in the absence of anti-miR-155, the whole degradation term becomes a regular first-order decay process. $\boldsymbol{M_0}$ represents the concentration of miR-155 at time zero, i.e., the initial condition. All nonzero initial conditions used in the model are listed in **Table S3**.

**Equation for unbound PD-L1 concentration on cancerous cells ($\boldsymbol{C^L_{P,\mathrm{free}}(t)}$):**

$$\frac{dC^L_{P,\mathrm{free}}(t)}{dt} = \overbrace{\frac{g_0^{\mathrm{PDL1}}}{1+\varepsilon_M \cdot M_L(t)}}^{Production} - \overbrace{\delta_{\mathrm{PDL1}} \cdot C^L_{P,\mathrm{free}}(t)}^{Degradation} - \overbrace{k_{\mathrm{on},P} \cdot C^L_{P,\mathrm{free}}(t) \cdot C^T_{P,\mathrm{free}}(t)}^{Binding\ to\ PD-1} -$$

$$\overbrace{k_{\mathrm{on,Ab1}} \cdot C^L_{P,\mathrm{free}}(t) \cdot C^L_{\mathrm{Ab1,free}}(t)}^{Binding\ to\ ICI} + \overbrace{k_{\mathrm{off},P} \cdot C^L_{P,\mathrm{bound}}(t)}^{Unbinding\ from\ PD-1} + \overbrace{k_{\mathrm{off,Ab1}} \cdot C^L_{\mathrm{Ab1,bound}}(t)}^{Unbinding\ from\ ICI},$$

$$C^L_{P,\mathrm{free}}(0) = C_0^{\mathrm{PDL1}} \qquad (\mathbf{S2})$$

There are six terms that contribute to the rate of change of unbound or free PD-L1 $\left(\boldsymbol{C^L_{P,\mathrm{free}}(t)}\right)$ (units, pM) on the surface of cancer cells. The production term is similar to the one presented in Eq. S1, except that the downregulation of production is due to the concentration of miR-155, governed by an efficiency factor $\boldsymbol{\varepsilon_M}$. The degradation term is a first-order decay process with decay constant $\boldsymbol{\delta_{\mathrm{PDL1}}}$. The third term shows how the free PD-L1 present on cancer cells can attach to its (free) receptor, PD-1, located on CD8$^+$ T cells $\left(\boldsymbol{C^T_{P,\mathrm{free}}(t)}\right)$. This is a second-order process with binding rate $\boldsymbol{k_{\mathrm{on},P}}$. Similarly, the fourth term represents how free PD-L1 can bind to an anti-PD-L1 immune checkpoint inhibitor (ICI) like atezolizumab, preventing it from binding to PD-1. The concentration of the free ICI in tumors is represented by $\boldsymbol{C^L_{\mathrm{Ab1,free}}(t)}$. This binding is also a second-order process with binding rate constant $\boldsymbol{k_{\mathrm{on,Ab1}}}$. The fifth and sixth terms are first-order processes with unbinding rate constants $\boldsymbol{k_{\mathrm{off},P}}$ and $\boldsymbol{k_{\mathrm{off,Ab1}}}$ which represent the reverse of the third and fourth processes, i.e., unbinding of the PD-L1/PD-1 complex ($\boldsymbol{C^L_{P,\mathrm{bound}}(t)}$) and atezolizumab/PD-L1 complex ($\boldsymbol{C^L_{\mathrm{Ab1,bound}}(t)}$), respectively, to reclaim free PD-L1. $\boldsymbol{C_0^{\mathrm{PDL1}}}$ is the initial condition of free PD-L1.

**Equation for the concentration of PD-L1/PD-1 complex between cancer cells and CD8⁺ T cells ($C^{L}_{P,\mathrm{bound}}(t)$):**

$$\frac{dC^{L}_{P,\mathrm{bound}}(t)}{dt} = \overbrace{k_{\mathrm{on},P} \cdot C^{L}_{P,\mathrm{free}}(t) \cdot C^{T}_{P,\mathrm{free}}(t)}^{Binding\ to\ PD-1} - \overbrace{k_{\mathrm{off},P} \cdot C^{L}_{P,\mathrm{bound}}(t)}^{Unbinding\ from\ PD-1}, \qquad C^{L}_{P,\mathrm{bound}}(0) = 0 \qquad \text{(S3)}$$

The rate of change of the PD-L1/PD-1 complex on cancer cells $\left(C^{L}_{P,\mathrm{bound}}(t)\right)$ (units, pM) is affected by two opposing mechanisms. One is the binding between PD-L1 on cancer cells and PD-1 on CD8⁺ T cells, which is the third term in Eq. S2. The other is the unbinding from PD-1, which is the fifth term in Eq. S2.

**Equation for miR-155 concentration in TAM ($M_{\mathrm{M\phi}}(t)$):**

$$\frac{dM_{\mathrm{M\phi}}(t)}{dt} = \overbrace{g^{M}_{0}}^{Production} - \overbrace{g_{\mathrm{tran}} \cdot M_{\mathrm{M\phi}}(t)}^{Exosomal\ tranfer} - \overbrace{\delta_{M} \cdot \left(1 + \frac{A_{\mathrm{AM},M} \cdot \mathrm{AM}_{\mathrm{M\phi}}(t)}{\mathrm{EC}_{50,\mathrm{AM}} + \mathrm{AM}_{\mathrm{M\phi}}(t)}\right) \cdot M_{\mathrm{M\phi}}(t)}^{Degradation},$$

$$M_{\mathrm{M\phi}}(0) = M_{0} \qquad \text{(S4)}$$

As mentioned in Eq. S1, TAMs also produce miR-155. The concentration of miR-155 within TAMs $\left(M_{\mathrm{M\phi}}(t)\right)$ (units, pM) is a function of three effects. The first is the overall production of miR-155, which is proportional to the production rate constant $g^{M}_{0}$. The second is the loss of miR-155 due to exosomal transfer, which corresponds to the second term of Eq. S1. The last term represents the degradation of miR-155 and is analogous to the degradation term in Eq. S1 and is now governed by the concentration of anti-miR-155 within TAMs ($\mathrm{AM}_{\mathrm{M\phi}}(t)$).

**Equation for unbound PD-L1 concentration on TAM ($C^{\mathrm{M\phi}}_{P,\mathrm{free}}(t)$):**

$$\frac{dC^{\mathrm{M\phi}}_{P,\mathrm{free}}(t)}{dt} = \overbrace{\frac{g_0^{\mathrm{PDL1}}}{1+\varepsilon_M \cdot M_{\mathrm{M\phi}}(t)}}^{Production} - \overbrace{\delta_{\mathrm{PDL1}} \cdot C^{\mathrm{M\phi}}_{P,\mathrm{free}}(t)}^{Degradation} - \overbrace{k_{\mathrm{on},P} \cdot C^{\mathrm{M\phi}}_{P,\mathrm{free}}(t) \cdot C^{T}_{P,\mathrm{free}}(t)}^{Binding\ to\ PD-1} -$$

$$\overbrace{k_{\mathrm{on,Ab1}} \cdot C^{\mathrm{M\phi}}_{P,\mathrm{free}}(t) \cdot C^{\mathrm{M\phi}}_{\mathrm{Ab1,free}}(t)}^{Binding\ to\ ICI} + \overbrace{k_{\mathrm{off},P} \cdot C^{\mathrm{M\phi}}_{P,\mathrm{bound}}(t)}^{Unbinding\ from\ PD-1} + \overbrace{k_{\mathrm{off,Ab1}} \cdot C^{\mathrm{M\phi}}_{\mathrm{Ab1,bound}}(t)}^{Unbinding\ from\ ICI},$$

$$C^{\mathrm{M\phi}}_{P,\mathrm{free}}(0) = C_0^{\mathrm{PDL1}} \qquad \textbf{(S5)}$$

The rate of change of the concentration of free PD-L1 on the surface of TAMs $\left(C^{\mathrm{M\phi}}_{P,\mathrm{free}}(t)\right)$ (units, pM) is governed by the same processes described in Eq. S2, the main difference being that the variables are now defined with respect to TAMs instead of cancer cells, thus the superscript $L$ has been replaced with $\mathrm{M\phi}$.

**Equation for concentration of PD-L1/PD-1 complex between TAMs and CD8⁺ T cells ($C^{\mathrm{M\phi}}_{P,\mathrm{bound}}(t)$):**

$$\frac{dC^{\mathrm{M\phi}}_{P,\mathrm{bound}}(t)}{dt} = \overbrace{k_{\mathrm{on},P} \cdot C^{\mathrm{M\phi}}_{P,\mathrm{free}}(t) \cdot C^{T}_{P,\mathrm{free}}(t)}^{Binding} - \overbrace{k_{\mathrm{off},P} \cdot C^{\mathrm{M\phi}}_{P,\mathrm{bound}}(t)}^{Unbinding}, \qquad C^{\mathrm{M\phi}}_{P,\mathrm{bound}}(0) = 0 \qquad \textbf{(S6)}$$

The equation describing the concentration of the PD-L1/PD-1 complex on TAMs $\left(C^{\mathrm{M\phi}}_{P,\mathrm{bound}}(t)\right)$ (units, pM) is analogous to Eq. S3, where the superscript $L$ has been replaced with $\mathrm{M\phi}$ to represent TAMs instead of cancer cells.

**Equation for unbound PD-1 concentration on CD8⁺ T cells ($C^{T}_{P,\mathrm{free}}(t)$):**

$$\frac{dC^{T}_{P,\mathrm{free}}(t)}{dt} = \overbrace{g_0^{\mathrm{PD1}}}^{Production} - \overbrace{\delta_{\mathrm{PD1}} \cdot C^{T}_{P,\mathrm{free}}(t)}^{Degradation} - \overbrace{k_{\mathrm{on},P} \cdot \left(C^{L}_{P,\mathrm{free}}(t) + C^{\mathrm{M\phi}}_{P,\mathrm{free}}(t)\right) \cdot C^{T}_{P,\mathrm{free}}(t)}^{Binding\ to\ PD-L1} -$$

$$\overbrace{k_{\mathrm{on,Ab2}} \cdot C^{T}_{P,\mathrm{free}}(t) \cdot C^{T}_{\mathrm{Ab2,free}}(t)}^{Binding\ to\ ICI} + \overbrace{k_{\mathrm{off},P} \cdot (C^{L}_{P,\mathrm{bound}}(t) + C^{\mathrm{M\phi}}_{P,\mathrm{bound}}(t))}^{Unbinding\ from\ PD-L1} + \overbrace{k_{\mathrm{off,Ab2}} \cdot C^{T}_{\mathrm{Ab2,bound}}(t)}^{Unbinding\ from\ ICI},$$

$$C^{T}_{P,\mathrm{free}}(0) = C_0^{\mathrm{PD1}} \qquad \textbf{(S7)}$$

Now that we have characterized the concentration of free PD-L1 on cancer cells and TAMs, we do the equivalent for PD-1 on CD8$^+$ T-cells. The rate of change of free PD-1 on CD8$^+$ T cells $\left(\boldsymbol{C}_{\boldsymbol{P},\mathbf{free}}^{\boldsymbol{T}}(\boldsymbol{t})\right)$ (units, pM) is similar to Eq. S2 or Eq. S5, with the following differences. The production rate of free PD-1 is a first-order process with no modulation. The third term incorporates the binding of PD-1 to PD-L1 on both cancer cells and TAMs, where both are second-order processes. Next, the anti-PD-1 ICI therapy such as pembrolizumab binds to PD-1 receptor, governed by the rate constant $\boldsymbol{k}_{\mathbf{on,Ab2}}$, preventing it from binding to PD-L1. Hence, we use the subscript $\mathbf{Ab2}$, instead of $\mathbf{Ab1}$, to distinguish between a drug targeting PD-1 and PD-L1, respectively. Lastly, the fifth and sixth terms represent the reverse of processes described in the third and fourth terms, respectively. $\boldsymbol{C}_{\mathbf{0}}^{\mathbf{PD1}}$ is the initial concentration of PD-1 on T cells.

*The next three equations characterize the population kinetics of key cells in the model, which collectively determine the volumetric tumor growth kinetics.*

**Equation for total TAM volume ($\mathbf{M\phi}(\boldsymbol{t})$):**

$$\frac{d\mathbf{M\phi}(t)}{dt} = \overbrace{\boldsymbol{\gamma}_{\mathbf{0}}^{\mathbf{M\phi}} \cdot \left(\frac{L(t)}{k_{L,\mathbf{M\phi}} + L(t)}\right)}^{Recruitment} - \overbrace{\boldsymbol{\delta}_{\mathbf{M\phi}} \cdot \mathbf{M\phi}(\boldsymbol{t})}^{Death}, \qquad \mathbf{M\phi(0) = M\phi_0 \neq 0} \qquad \textbf{(S8)}$$

The rate of change of TAM population $\left(\mathbf{M\phi}(\boldsymbol{t})\right)$ (units, cm$^3$) in the tumor depends on the recruitment of TAMs into the tumor and their death. The first term represents the recruitment of TAMs into the tumor, which is positively regulated by the population of cancer cells ($\boldsymbol{L}(\boldsymbol{t})$) in the tumor and has an upper bound at the maximum possible recruitment rate $\boldsymbol{\gamma}_{\mathbf{0}}^{\mathbf{M\phi}}$. $\boldsymbol{k}_{\boldsymbol{L},\mathbf{M\phi}}$ denotes the potency of cancer cells to promote the recruitment of TAMs. The second term represents the death of TAMs and is modeled as a first-order process governed by the death rate constant $\boldsymbol{\delta}_{\mathbf{M\phi}}$.

**Equation for total CD8$^+$ T-cell volume ($\mathbf{CD8}(\boldsymbol{t})$):**

$$\frac{d\mathbf{CD8}(t)}{dt} = \overbrace{\gamma_0^{\mathbf{CD8}} \cdot \left(\frac{L(t)}{k_{L,\mathbf{CD8}}+L(t)}\right)}^{Infiltration} - \overbrace{\delta_{\mathbf{CD8}} \cdot \mathbf{CD8}(t)}^{Death}, \qquad \mathbf{CD8}(0) = CD8_0 \neq 0 \qquad (\mathbf{S9})$$

The rate of change of CD8+ T-cell population $(\mathbf{CD8}(t))$ (units, cm$^3$) in the tumor follows the same structure as Eq. S8. In this case, the maximum infiltration rate of cells is $\gamma_0^{\mathbf{CD8}}$ and the potency of cancer cells to attract CD8+ T cells is $k_{L,\mathbf{CD8}}$. Cell death is determined by the death rate constant $\delta_{\mathbf{CD8}}$.

##### Equation for total cancer cell volume ($L(t)$):

$$\frac{dL(t)}{dt} = \overbrace{\gamma \cdot L(t) \cdot H(t)}^{Proliferation} - \overbrace{\frac{\delta_{\mathrm{immun}}}{1+\varepsilon\cdot(C_{P,\mathbf{bound}}^{L}(t)+C_{P,\mathbf{bound}}^{\mathrm{M\phi}}(t))} \cdot L(t) \cdot \mathbf{CD8}(t)}^{CD8^{+}\ T-cell-induced\ death} -$$

$$\overbrace{\delta_{\mathbf{chemo}} \cdot \left(\frac{C_{\mathbf{chemo},L}(t)}{\mathbf{EC}_{50,\mathbf{chemo}}\cdot H(t)+C_{\mathbf{chemo},L}(t)}\right) \cdot L(t)}^{Chemotherapy-induced\ death}, \qquad H(t) = \left(1 + \frac{A_{M,L}\cdot M_L(t)}{k_M+M_L(t)}\right),\ L(0) = L_0 \qquad (\mathbf{S10})$$

The population kinetics of cancer cells $(L(t))$ (units, cm$^3$) in the tumor depends upon 3 factors: cell proliferation, CD8+ T cell-mediated death, and chemotherapy-induced death. Assuming exponential growth, proliferation is a first-order process governed by the rate constant $\gamma$. As previously described, cancer cell proliferation is positively regulated by miR-155 expression in cancer cells. Thus, this regulation is modeled as a stimulatory process represented by the regulating function $H(t)$, which is proportional to the concentration of miR-155 in cancer cells ($M_L(t)$) as governed by the potency factor $k_M$ and activation factor $A_{M,L}$. Based on this, note that when the concentration of miR-155 inside the cancer cells is zero, cancer cell proliferation is unaffected and occurs at its intrinsic rate $\gamma \cdot L(t)$. However, as $M_L(t)$ increases, proliferation increases, and the smaller the Michaelis constant $k_M$ is, the faster the kinetics to reach the asymptotic value $\gamma \cdot L(t) \cdot (1 + A_{M,L})$.

The second term represents CD8+ T cell-mediated death of cancerous cells. This is a second-order process as it depends on the interaction between CD8+ T-cells and cancer cells.

The death rate $\boldsymbol{\delta}_{\mathbf{immun}}$ governing this process is modulated by two variables, $\boldsymbol{C}_{\boldsymbol{P},\mathbf{bound}}^{\boldsymbol{L}}(\boldsymbol{t})$ from Eq. S3 and $\boldsymbol{C}_{\boldsymbol{P},\mathbf{bound}}^{\mathbf{M\Phi}}(\boldsymbol{t})$ from Eq. S6, which represent the level of PD-L1/PD-1 complexes on tumor cells and TAMs, respectively. The more PD-L1/PD-1 complexes form, the more CD8[+] T-cells become deactivated, preventing their cytotoxic effect on cancer cells as governed by an efficiency factor $\boldsymbol{\varepsilon}$,. Note that the positive effect of ICI therapy on CD8[+] T cell-mediated death is implicitly included by the negative influence of ICI on PD-L1/PD-1 complex formation in Eqs. S5 and S7.

The third term represents death due to chemotherapy, which depends upon the tumor concentration of the drug ($\boldsymbol{C}_{\mathbf{chemo},\boldsymbol{L}}(\boldsymbol{t})$) and its potency defined by $\mathbf{EC}_{\mathbf{50,chemo}}$. The larger the concentration of the chemotherapeutic agent in cancer cells and/or the smaller the $\mathbf{EC}_{\mathbf{50,chemo}}$, the greater the cell death. However, we note that the $\mathbf{EC}_{\mathbf{50,chemo}}$ of the drug is being affected by the regulating function $\boldsymbol{H}(\boldsymbol{t})$. Similar to its influence on tumor growth, $\boldsymbol{H}(\boldsymbol{t})$ captures the effect of miR-155 on chemoresistance. In the absence of miR-155, $\boldsymbol{H}(\boldsymbol{t}) = \mathbf{1}$ and $\mathbf{EC}_{\mathbf{50,chemo}}$ remains unchanged. Otherwise, $\boldsymbol{H}(\boldsymbol{t})$ approaches the asymptotic value $\mathbf{1} + \boldsymbol{A}_{\boldsymbol{M},\boldsymbol{L}}$ as the concentration of miR-155 increases, thereby increasing $\mathbf{EC}_{\mathbf{50,chemo}}$ and making the drug less potent. $\boldsymbol{L}_{\mathbf{0}}$ is the initial cancer cell volume. Note that the total tumor volume kinetics is obtain by combining the solutions of Eqs. S8,S9,S10.

*The remaining equations describe the kinetics of the drug delivery system, i.e., nanoparticles, its cargo anti-miR-155, ICI immunotherapies, and chemotherapy.*

**Equation for NP mass kinetics in systemic circulation ($\boldsymbol{N}_{\boldsymbol{P}}(\boldsymbol{t})$):**

$$\frac{dN_P(t)}{dt} = \overbrace{-P_{\mathrm{NP}} \cdot S \cdot N_P(t)}^{\text{Delivery to tumor}} - \overbrace{k_{\mathrm{Cl}} \cdot N_P(t)}^{\text{Clearance}} - \overbrace{\delta_{\mathrm{NP}} \cdot N_P(t)}^{\text{Degradation}},$$

$$N_P(t) = \begin{cases} 0, & t = 0 \\ N_P(t^-) + N_0, & t = t_i\,, t_i \text{ in } S_N \end{cases} \qquad \textbf{(S11)}$$

The rate of change of NPs in the plasma compartment $(\boldsymbol{N_P(t)})$ (units, %ID) is a negative function. This implies that once $\boldsymbol{N_0}$ NPs are injected at the time $\boldsymbol{t = t_i}$, the function $\boldsymbol{N_P(t)}$ increases by $\boldsymbol{N_0}$ and immediately starts to decrease monotonically. The notation $\boldsymbol{N_P(t^-)}$ represents the number of NPs just before the time $\boldsymbol{t}$. The injection times $\boldsymbol{t_i}$ belong to a predefined set of times, which we denote by $\boldsymbol{S_N}$. There are three mechanisms that modify $\boldsymbol{N_P(t)}$. The first is the delivery of NPs from the bloodstream into the tumor interstitium. This depends on the tumor microvascular surface area $\boldsymbol{S}$ and the tumor microvascular permeability $\boldsymbol{P_{\mathrm{NP}}}$. The second term denotes the clearance of NPs with rate $\boldsymbol{k_{\mathrm{Cl}}}$, and the third represents the degradation of NPs with rate $\boldsymbol{\delta_{\mathrm{NP}}}$.

**Equation for NP mass kinetics in the tumor interstitium ($N_I(t)$):**

$$\frac{dN_I(t)}{dt} = \overbrace{\boldsymbol{P_{\mathrm{NP}} \cdot S \cdot N_P(t)}}^{Influx\ from\ plasma} - \overbrace{\frac{D_{\mathrm{NP}}}{\mathrm{Len}^2} \cdot \boldsymbol{N_I(t)}}^{Diffusion\ to\ cells} - \overbrace{\boldsymbol{\delta_{\mathrm{NP}} \cdot N_I(t)}}^{Degradation}, \qquad \boldsymbol{N_I(0) = 0} \qquad (\mathbf{S12})$$

The mass of NPs in the tumor interstitium $(\boldsymbol{N_I(t)})$ depends on three processes. The first term is equivalent to the one in Eq. S11 and denotes the incoming NPs from the plasma compartment. The second term represents the rate of diffusion through the interstitium to the nearby cells, which is proportional to the local diffusivity of NPs, $\boldsymbol{D_{\mathrm{NP}}}$, and the characteristic length $(\mathbf{Len})$ of the intercapillary distance in the tumor interstitium. The last term is analogous to the third term in Eq. S11 representing NP degradation. NPs are delivered to the various cells in the tumor while diffusing through the interstitium, and we assume that the extent of delivery to each cell type depends on their population fraction in the tumor.

**Equation for NP mass kinetics in cancer cells ($N_L(t)$):**

$$\frac{dN_L(t)}{dt} = \overbrace{\frac{L(t)}{L(t)+\mathrm{M\phi}(t)+\mathrm{CD8}(t)} \cdot \frac{D_{\mathrm{NP}}}{\mathrm{Len}^2} \cdot \boldsymbol{N_I(t)}}^{Delivery\ from\ interstitium} - \overbrace{\boldsymbol{\delta_{\mathrm{NP}} \cdot N_L(t)}}^{Degradation}, \qquad \boldsymbol{N_L(0) = 0} \qquad (\mathbf{S13})$$

The first term describes the mass of NPs delivered to the cancer cells, which is proportional to the population fraction of cancer cells in the tumor. Hence, the second term in Eq. S12 reappears in Eq. S13 multiplied by the ratio of cancer cell volume to the total tumor volume (i.e., cancer cells + TAMs + CD8⁺ T cells). The second term represents NP degradation.

**Equation for NP mass kinetics in TAMs ($N_{\mathrm{M\phi}}(t)$):**

$$\frac{dN_{\mathrm{M\phi}}(t)}{dt} = \overbrace{\frac{\mathrm{M\phi}(t)}{L(t)+\mathrm{M\phi}(t)+\mathrm{CD8}(t)} \cdot \frac{D_{\mathrm{NP}}}{\mathrm{Len}^2} \cdot \boldsymbol{N_I(t)}}^{Delivery\ from\ interstitium} - \overbrace{\boldsymbol{\delta_{\mathrm{NP}} \cdot N_{\mathrm{M\phi}}(t)}}^{Degradation}, \qquad \boldsymbol{N_{\mathrm{M\phi}}(0) = 0} \qquad (\mathbf{S14})$$

The equation governing the mass of NPs in TAMs $\left(\boldsymbol{N_{\mathrm{M\phi}}(t)}\right)$ is analogous to Eq. S13. In this case, the fraction of NPs delivered to the TAMs is proportional to the population fraction of TAMs in the tumor. The second term denotes degradation. Note that the remaining NPs in the interstitium are delivered to CD8⁺ T-cells, thereby conserving NP mass. We do not describe an equation for this process since cargo delivery to CD8⁺ T cells does not elicit a pharmacological effect in our model.

**Equation for anti-miR-155 concentration kinetics in cancer cells ($\mathbf{AM}_L(t)$):**

$$\frac{d\mathrm{AM}_L(\mathrm{t})}{dt} = \overbrace{\boldsymbol{k_{\mathrm{rel}} \cdot \textstyle\sum_{t_i \text{ in } S_N}\left(\mathbf{1}_{t\geq t_i}(t) \cdot N_L(t) \cdot \mathrm{AM}_0 \cdot L^{-1}(t) \cdot e^{-k_{\mathrm{rel}}\cdot(t-t_i)}\right)}}^{Release} - \overbrace{\boldsymbol{\delta_{\mathrm{AM}} \cdot \mathrm{AM}_L(\mathrm{t})}}^{Degradation},$$

$$\mathbf{AM}_L(\mathbf{0}) = \mathbf{0} \qquad (\mathbf{S15})$$

Once the NPs are delivered to the cancer cells, they release the therapeutic agent anti-miR-155. The concentration of anti-miR-155 in cancer cells $(\mathbf{AM}_L(\mathbf{t}))$ (units, $\mathbf{mg \cdot mL^{-1}}$) thus depends on the release rate of the cargo (first term) and the degradation rate of anti-miR-155 (second term). Assuming each NP contains $\mathbf{AM_0}$ mg of anti-miR-155, the total available amount is given by the product of $\mathbf{AM_0}$ and the number of NPs inside the cancer cells $\boldsymbol{N_L(t)}$. If we divide this quantity by the total volume of cancer cells $\boldsymbol{L(t)}$, we obtain the maximum theoretical concentration of anti-miR-155 at time $\boldsymbol{t}$, i.e., $\boldsymbol{N_L(t) \cdot \mathrm{AM}_0 \cdot L^{-1}(t)}$. Assuming first-order release at rate $\boldsymbol{k_{\mathrm{rel}}}$, anti-miR-155 is released from NPs following a cumulative exponential decay rate model with release constant $\boldsymbol{k_{\mathrm{rel}}}$. Note that the argument of the sum is multiplied by the indicator function $\mathbf{1}_{t\geq t_i}(\boldsymbol{t})$, which is zero for $t < t_i$, and one otherwise. This is important because we are summing over all injection times $t_i$ in the set $S_N$, and if the current time $t$ is smaller than some $t_i$, then that means that the $i$-th injection has not yet taken place and the whole argument should be zero for that index. The second term is a first-order decay process with decay constant $\delta_{\mathrm{AM}}$.

**Equation for anti-miR-155 concentration kinetics in TAMs ($\mathrm{AM}_{\mathrm{M\Phi}}(t)$):**

$$\frac{d\mathrm{AM}_{\mathrm{M\Phi}}(\mathrm{t})}{dt} = \overbrace{k_{\mathrm{rel}} \cdot \sum_{t_i \text{ in } S_N}\left(\mathbf{1}_{t \geq t_i}(t) \cdot N_{\mathrm{M\Phi}}(t) \cdot \mathrm{AM}_0 \cdot \mathrm{M\Phi}^{-1}(t) \cdot e^{-k_{\mathrm{rel}} \cdot (t - t_i)}\right)}^{Release} - \overbrace{\delta_{\mathrm{AM}} \cdot \mathrm{AM}_{\mathrm{M\Phi}}(\mathrm{t})}^{Degradation},$$

$$\mathrm{AM}_{\mathrm{M\Phi}}(0) = 0 \qquad \textbf{(S16)}$$

The concentration of anti-miR-155 in TAMs $\left(\mathrm{AM}_{\mathrm{M\Phi}}(\mathrm{t})\right)$ (units, $\mathrm{mg} \cdot \mathrm{mL}^{-1}$) is a function of the same processes introduced in Eq. S15, with the role of cancer cells replaced by TAMs.

**Equation for anti-PD-L1 antibody concentration kinetics in plasma ($C_{\mathrm{Ab1},P}(t)$):**

$$\frac{dC_{\mathrm{Ab1},P}(t)}{dt} = \overbrace{k_{\mathrm{abs}} \cdot \sum_{t_i \text{ in } S_I}\left(\mathbf{1}_{t \geq t_i}(t) \cdot \mathrm{Dose}_{\mathrm{Ab1}} \cdot V_{\mathrm{Ab1}}^{-1} \cdot e^{-k_{\mathrm{abs}} \cdot (t - t_i)}\right)}^{Absorption\ from\ peritoneal\ cavity} - \overbrace{P_{\mathrm{Ab}} \cdot S \cdot C_{\mathrm{Ab1},P}(t)}^{Delivery\ to\ tumor} -$$

$$\overbrace{\mathrm{Cl}_{\mathrm{Ab1}} \cdot V_{\mathrm{Ab1}}^{-1} \cdot C_{\mathrm{Ab1},P}(t)}^{Clearance} - \overbrace{\delta_{\mathrm{Ab}} \cdot C_{\mathrm{Ab1},P}(t)}^{Degradation}, \qquad C_{\mathrm{Ab1},P}(0) = 0 \qquad \textbf{(S17)}$$

In addition to anti-miR-155 therapy, we also used ICI in the form of anti-PD-L1 and anti-PD-1 antibodies. The concentration of anti-PD-L1 in the plasma compartment is denoted by $C_{\mathrm{Ab1},P}(t)$. ICI is injected into the peritoneal cavity in the case of *in vivo* studies and the plasma compartment in the clinical scenario at times $t_i$, which belong to the set $S_I$. For the preclinical scenario, the first term describes the absorption kinetics from the peritoneal cavity and has a similar structure to the first term in Eq. S16, where the release rate $k_{\mathrm{rel}}$ is replaced by the systemic absorption rate $k_{\mathrm{abs}}$. The key difference between the first terms of Eq. S16 and Eq. S17 are the concentrations of the drug. Since a unit dose of the antibody is represented by $\mathrm{Dose}_{\mathrm{Ab1}}$ (units, mg), and the volume of distribution of anti-PD-L1 antibody is $V_{\mathrm{Ab1}}$, the local concentration in the peritoneal cavity is

$\mathbf{Dose_{Ab1}} \cdot \boldsymbol{V_{Ab1}^{-1}}$. These antibodies will be continuously absorbed at rate $\boldsymbol{k_{abs}}$ into the plasma following the kinetics given in Eq. S17 until the source is depleted.

Notably, because we model intravenous injection in the clinical scenario, the first term of Eq. S17 is set to zero, while the initial condition following the first injection is equated to $\mathbf{Dose_{Ab1}} \cdot \boldsymbol{V_{Ab1}^{-1}}$. Thus, for the entire clinical treatment involving multiple injections, the initial condition can be described as:

$$\boldsymbol{C_{\mathrm{Ab1},P}(t)} = \begin{cases} \mathbf{0}, \quad \boldsymbol{t = 0} \\ \boldsymbol{C_{\mathrm{Ab1},P}(t^-)} + \mathbf{Dose_{Ab1}} \cdot \boldsymbol{V_{\mathrm{Ab1}}^{-1}}, \boldsymbol{t = t_i}, \boldsymbol{t_i} \text{ in } \boldsymbol{S_I} \end{cases}.$$

The notation $\boldsymbol{C_{\mathrm{Ab1},P}(t^-)}$ represents ICI concentration just before the time $\boldsymbol{t}$. The injection times $\boldsymbol{t_i}$ belong to a predefined set of times, which we denote by $\boldsymbol{S_I}$., such that $\boldsymbol{t_i} \in \boldsymbol{S_I}$.

Once inside the plasma compartment, some antibodies will extravasate to the tumor interstitium in a permeation-limited fashion. This is shown in the second term of Eq. S17 and is analogous to the first term in Eq. S12. The third term describes how another portion of the antibodies will be cleared from the systemic circulation at a rate $\mathbf{Cl_{Ab1}}$. Lastly, antibodies in the plasma will also be degraded following a first-order process with decay constant $\boldsymbol{\delta_{\mathrm{Ab}}}$ as described in the fourth term.

**Equation for anti-PD-L1 antibody concentration kinetics in the tumor interstitium ($\boldsymbol{C_{\mathrm{Ab1},I}(t)}$):**

$$\frac{d\boldsymbol{C_{\mathrm{Ab1},I}(t)}}{dt} = \overbrace{\boldsymbol{P_{\mathrm{Ab}} \cdot S \cdot C_{\mathrm{Ab1},P}(t) \cdot \frac{V_{\mathrm{Ab1}}}{V_{T,I}(t)}}}^{\textit{Delivery from plasma}} - \overbrace{\frac{\boldsymbol{D_{\mathrm{Ab}}}}{\mathbf{Len^2}} \cdot \boldsymbol{C_{\mathrm{Ab1},I}(t)}}^{\textit{Diffusion to cells}} - \overbrace{\boldsymbol{\delta_{\mathrm{Ab}} \cdot C_{\mathrm{Ab1},I}(t)}}^{\textit{Degradation}},$$

$$\boldsymbol{C_{\mathrm{Ab1},I}(0) = 0} \qquad \textbf{(S18)}$$

The concentration of anti-PD-L1 antibodies in the tumor interstitium $\left(\boldsymbol{C_{\mathrm{Ab1},I}(t)}\right)$ relies on 3 effects. One is the permeation-limited delivery from the plasma into the interstitium as shown in the first term of Eq. S18, with $\boldsymbol{V_{T,I}(t)}$ being the volume of tumor interstitium, which is assumed to be a constant fraction ($\boldsymbol{f_I}$ = 27.5%) of the total tumor volume (1). The second is the diffusion of antibodies through the interstitium to the cancer cells and TAMs, which is analogous to the diffusion of NPs as described in Eq. S12. The third is degradation, which is analogous to the last term of Eq. S17.

**Equation for unbound anti-PD-L1 antibody concentration kinetics near the membrane of cancer cells ($\boldsymbol{C^L_{\text{Ab1,free}}(t)}$):**

$$\frac{dC^L_{\text{Ab1,free}}(t)}{dt} = \overbrace{\frac{V_{T,I}(t)}{L(t)+\text{M}\phi(t)+\text{CD8}(t)} \cdot \frac{D_{\text{Ab}}}{\text{Len}^2} \cdot C_{\text{Ab1},I}(t)}^{Delivery\ from\ interstitium} - \overbrace{\delta_{\text{Ab}} \cdot C^L_{\text{Ab1,free}}(t)}^{Degradation} -$$

$$\overbrace{k_{\text{on,Ab1}} \cdot C^L_{P,\text{free}}(t) \cdot C^L_{\text{Ab1,free}}(t)}^{Binding\ to\ PD-L1} + \overbrace{k_{\text{off,Ab1}} \cdot C^L_{\text{Ab1,bound}}(t)}^{Unbinding\ from\ PD-L1}, \qquad C^L_{\text{Ab1,free}}(0) = 0 \qquad \textbf{(S19)}$$

Similar to the process where the NPs diffused from the tumor interstitium, in Eq. S12, towards the cancer cells, in Eq. S13, the first term of Eq. S19 shows the analogous process with the immunotherapy located on the vicinity of cancer cells $\left(\boldsymbol{C^L_{\text{Ab1,free}}(t)}\right)$. This expression is also derived based on the assumption that mass of ICI delivered to the cancer cells is proportional to the population fraction of cancer cells in the tumor. The second term represents degradation of ICI. The third term is a second-order binding process between the ICI and the PD-L1 ligand on cancer cells and is analogous to the fourth term of Eq. S2. Likewise, the last term is analogous to the unbinding process shown in the last term of Eq. S2.

**Equation for the concentration of the anti-PD-L1 antibody/PD-L1 complex on cancer cells ($\boldsymbol{C^L_{\text{Ab1,bound}}(t)}$):**

$$\frac{dC^L_{\text{Ab1,bound}}(t)}{dt} = \overbrace{k_{\text{on,Ab1}} \cdot C^L_{P,\text{free}}(t) \cdot C^L_{\text{Ab1,free}}(t)}^{Binding} - \overbrace{k_{\text{off,Ab1}} \cdot C^L_{\text{Ab1,bound}}(t)}^{Unbinding},$$

$$C^L_{\text{Ab1,bound}}(0) = 0 \qquad \textbf{(S20)}$$

The concentration of the anti-PD-L1/PD-L1 complex on the surface of cancer cells $\left(C^{L}_{\mathrm{Ab1,bound}}(t)\right)$ is a direct consequence of the last two terms in Eq. S19, i.e., the balance between binding between binding and unbinding of the ICI to the PD-L1 ligand.

**Equation for unbound anti-PD-L1 antibody concentration kinetics near the TAM membrane ($C^{\mathrm{M\phi}}_{\mathrm{Ab1,free}}(t)$):**

$$\frac{dC^{\mathrm{M\phi}}_{\mathrm{Ab1,free}}(t)}{dt} = \overbrace{\frac{V_{T,I}(t)}{L(t)+\mathrm{M\phi}(t)+\mathrm{CD8}(t)} \cdot \frac{D_{\mathrm{Ab}}}{\mathrm{Len}^2} \cdot C_{\mathrm{Ab},I}(t)}^{Delivery\ from\ interstitium} - \overbrace{\delta_{\mathrm{Ab}} \cdot C^{\mathrm{M\phi}}_{\mathrm{Ab1,free}}(t)}^{Degradation} - \overbrace{k_{\mathrm{on,Ab1}} \cdot C^{\mathrm{M\phi}}_{P,\mathrm{free}}(t) \cdot C^{\mathrm{M\phi}}_{\mathrm{Ab1,free}}(t)}^{Binding\ to\ PD-L1} + \overbrace{k_{\mathrm{off,Ab1}} \cdot C^{\mathrm{M\phi}}_{\mathrm{Ab1,bound}}(t)}^{Unbinding\ from\ PD-L1}, \quad C^{\mathrm{M\phi}}_{\mathrm{Ab1,free}}(0) = 0 \qquad \mathbf{(S21)}$$

Eq. S21 characterizes the concentration of free ICI antibodies close to TAMs $\left(C^{\mathrm{M\phi}}_{\mathrm{Ab1,free}}(t)\right)$. This equation is analogous to Eq. S19, but instead of taking place in the vicinity of cancer cells, it takes place near TAMs.

**Equation for the concentration of the anti-PD-L1 antibody/PD-L1 complex on TAMs ($C^{\mathrm{M\phi}}_{\mathrm{Ab1,bound}}(t)$):**

$$\frac{dC^{\mathrm{M\phi}}_{\mathrm{Ab1,bound}}(t)}{dt} = \overbrace{k_{\mathrm{on,Ab1}} \cdot C^{\mathrm{M\phi}}_{P,\mathrm{free}}(t) \cdot C^{\mathrm{M\phi}}_{\mathrm{Ab1,free}}(t)}^{Binding} - \overbrace{k_{\mathrm{off,Ab1}} \cdot C^{\mathrm{M\phi}}_{\mathrm{Ab1,bound}}(t)}^{Unbinding}, \quad C^{\mathrm{M\phi}}_{\mathrm{Ab1,bound}}(0) = 0 \qquad \mathbf{(S22)}$$

This equation is analogous to Eq. S20 but centered around the TAMs.

**Equation for anti-PD-1 antibody concentration kinetics in plasma ($C_{\mathrm{Ab2},P}(t)$):**

$$\frac{dC_{\mathrm{Ab2},P}(t)}{dt} = \overbrace{k_{\mathrm{abs}} \cdot \sum_{t_i \text{ in } S_I}\left(1_{t\geq t_i}(t) \cdot \mathrm{Dose}_{\mathrm{Ab2}} \cdot V^{-1}_{\mathrm{Ab2}} \cdot e^{-k_{\mathrm{abs}}\cdot(t-t_i)}\right)}^{Absorption\ from\ peritoneal\ cavity} - \overbrace{P_{\mathrm{Ab}} \cdot S \cdot C_{\mathrm{Ab2},P}(t)}^{Delivery\ to\ tumor} - \overbrace{\mathrm{Cl}_{\mathrm{Ab2}} \cdot V^{-1}_{\mathrm{Ab2}} \cdot C_{\mathrm{Ab2},P}(t)}^{Clearance} - \overbrace{\delta_{\mathrm{Ab}} \cdot C_{\mathrm{Ab2},P}(t)}^{Degradation}, \quad C_{\mathrm{Ab2},P}(t) = 0 \qquad \mathbf{(S23)}$$

This equation mirrors Eq. S17, but instead of referring to the ICI targeting PD-L1 (subscript $\mathbf{Ab1}$), it applies to the ICI targeting the CD8+ T-cell receptor PD-1 (subscript $\mathbf{Ab2}$). Similarly, the *in vivo* scenario follows intraperitoneal injection, while the clinical scenario is based on intravenous injection.

**Equation for anti-PD-1 antibody concentration kinetics in the tumor interstitium ($C_{\mathrm{Ab2},I}(t)$):**

$$\frac{dC_{\mathrm{Ab2},I}(t)}{dt} = \overbrace{P_{\mathrm{Ab}} \cdot S \cdot C_{\mathrm{Ab2},P}(t) \cdot \frac{V_{\mathrm{AB2}}}{V_{T,I}(t)}}^{Delivery\ from\ plasma} - \overbrace{\frac{D_{\mathrm{Ab}}}{\mathrm{Len}^2} \cdot C_{\mathrm{Ab2},I}(t)}^{Diffusion\ to\ cells} - \overbrace{\delta_{\mathrm{Ab}} \cdot C_{\mathrm{Ab2},I}(t)}^{Degradation},$$

$$C_{\mathrm{Ab2},I}(0) = 0 \qquad (\mathbf{S24})$$

Similarly, Eq. S24 is analogous to Eq. S18.

**Equation for unbound anti-PD-1 antibody concentration kinetics near the CD8+ T-cell membrane ($C^T_{\mathrm{Ab2,free}}(t)$):**

$$\frac{dC^T_{\mathrm{Ab2,free}}(t)}{dt} = \overbrace{\frac{V_{T,I}(t)}{L(t)+\mathrm{M\phi}(t)+\mathrm{CD8}(t)} \cdot \frac{D_{\mathrm{Ab}}}{\mathrm{Len}^2} \cdot C_{\mathrm{Ab2},I}(t)}^{Delivery\ from\ interstitium} - \overbrace{\delta_{\mathrm{Ab}} \cdot C^T_{\mathrm{Ab2,free}}(t)}^{Degradation} -$$

$$\overbrace{k_{\mathrm{on,Ab2}} \cdot C^T_{P,\mathrm{free}}(t) \cdot C^T_{\mathrm{Ab2,free}}(t)}^{Binding\ to\ PD-1} + \overbrace{k_{\mathrm{off,Ab2}} \cdot C^T_{\mathrm{Ab2,bound}}(t)}^{Unbinding\ from\ PD-1}, \qquad C^T_{\mathrm{Ab2,free}}(0) = 0 \qquad (\mathbf{S25})$$

This equation is analogous to Eq. S19 or Eq. S21, with the delivery to cancer cells or TAMs replaced by delivery to T cells.

**Equation for concentration of anti-PD-1 antibody/PD-1 complex ($C^T_{\mathrm{Ab2,bound}}(t)$):**

$$\frac{dC^T_{\mathrm{Ab2,bound}}(t)}{dt} = \overbrace{k_{\mathrm{on,Ab2}} \cdot C^T_{P,\mathrm{free}}(t) \cdot C^T_{\mathrm{Ab2,free}}(t)}^{Binding\ to\ PD-1} - \overbrace{k_{\mathrm{off,Ab2}} \cdot C^T_{\mathrm{Ab2,bound}}(t)}^{Unbinding\ from\ PD-1},$$

$$C^T_{\mathrm{Ab2,bound}}(0) = 0 \qquad (\mathbf{S26})$$

Similarly, this equation is analogous to Eq. S20 or Eq. S22, but now the complex forms on the surface of CD8+ T-cells.

So far, we have described equations for the therapeutic agents anti-miR-155, anti-PD-L1, and anti-PD-1. Next, we characterize the equations for the chemotherapy.

**Equation for chemotherapy concentration kinetics in plasma ($C_{\text{chemo},P}(t)$):**

$$\frac{dC_{\text{chemo},P}(t)}{dt} = \overbrace{k_{\text{abs,chemo}} \cdot \sum_{t_i \text{ in } S_C} \left(1_{t \geq t_i}(t) \cdot \text{Dose}_{\text{Chemo}} \cdot V_{\text{chemo}}^{-1} \cdot e^{-k_{\text{abs,chemo}} \cdot (t-t_i)}\right)}^{\textit{Absorption from peritoneal cavity}} -$$

$$\overbrace{D_{\text{chemo}} \cdot \frac{\left(C_{\text{chemo,P}}(t) - C_{\text{chemo,I}}(t)\right)}{\Delta x} \cdot S \cdot \frac{V_{\text{tumor}}(t)}{V_{\text{chemo}}}}^{\textit{Delivery to tumor}} - \overbrace{\text{Cl}_{\text{chemo}} \cdot V_{\text{chemo}}^{-1} \cdot C_{\text{chemo},P}(t)}^{\textit{Clearance}} - \overbrace{\beta_{\text{chemo}} \cdot C_{\text{chemo},P}(t)}^{\textit{Degradation}},$$

$$C_{\text{chemo},P}(t) = 0 \qquad \textbf{(S27)}$$

This equation follows a similar structure to that given in Eq. S17. One difference is that the injection times $t_i$ belong to a different set of times $S_C$ (i.e., $t_i \in S_C$), and the systemic absorption rate is given by $k_{\text{abs,chemo}}$. The main difference is the second term that expresses the transport of the chemotherapeutic agent from the plasma compartment into the tumor interstitium through the capillary walls of thickness $\Delta x$. This term represents diffusive flux and is proportional to the concentration gradient between these two compartments $\left(C_{\text{chemo,P}}(t) - C_{\text{chemo,I}}(t)\right) \cdot (\Delta x)^{-1}$. The proportionality constants are the chemotherapy diffusivity $D_{\text{chemo}}$, and the tumor microvascular surface area $S$. Here, $V_{\text{chemo}}$ is the volume of distribution of the chemotherapeutic agent and $V_{\text{tumor}}(t)$ is the total tumor volume at a given time $t$.

$V_{\text{tumor}}(t)$ is calculated as $\left(L(t) + \text{M}\phi(t) + \text{CD8}(t)\right)/(1 - f_I - f_V)$, where $f_I$ is the interstitial volume fraction of the tumor ($f_I$ = 27.5%) (1), and $f_V$ is the vascular volume fraction of the tumor ($f_V$ = 17%) (2). Lastly, the clearance and degradation rate of the chemotherapy are given by $\text{Cl}_{\text{chemo}}$ and $\beta_{\text{chemo}}$, respectively. Note that similar to prior therapies, chemotherapy was modeled via intraperitoneal injection in the *in vivo* scenario and intravenous injection in the clinical scenario. Thus, the first term of Eq. S27 is set to zero in the latter scenario, while the initial condition following the first injection is equated to $\mathbf{Dose_{chemo}} \cdot \boldsymbol{V}_{\mathbf{chemo}}^{-1}$. Thus, for the entire clinical treatment involving multiple injections, the initial condition can be described as:

$$\boldsymbol{C}_{\mathbf{chemo},\boldsymbol{P}}(\boldsymbol{t}) = \begin{cases} \mathbf{0}, \quad \boldsymbol{t} = \mathbf{0} \\ \boldsymbol{C}_{\mathbf{chemo},\boldsymbol{P}}(\boldsymbol{t}^{-}) + \mathbf{Dose_{chemo}} \cdot \boldsymbol{V}_{\mathbf{chemo}}^{-1}, \boldsymbol{t} = \boldsymbol{t}_i\,, \boldsymbol{t}_i \text{ in } \boldsymbol{S}_{\boldsymbol{C}} \end{cases}.$$

The notation $\boldsymbol{C}_{\mathbf{chemo},\boldsymbol{P}}(\boldsymbol{t}^{-})$ represents concentration of chemotherapy just before the time $\boldsymbol{t}$. The injection times $\boldsymbol{t}_i$ belong to a predefined set of times, which we denote by $\boldsymbol{S}_{\boldsymbol{C}}$.

**Equation for chemotherapy concentration kinetics in the tumor interstitium ($\boldsymbol{C}_{\mathbf{chemo},\boldsymbol{I}}(\boldsymbol{t})$):**

$$\frac{d\boldsymbol{C}_{\mathbf{chemo},\boldsymbol{I}}(\boldsymbol{t})}{d\boldsymbol{t}} = \overbrace{\boldsymbol{D}_{\mathbf{chemo}} \cdot \frac{\left(\boldsymbol{C}_{\mathbf{chemo,P}}(\boldsymbol{t}) - \boldsymbol{C}_{\mathbf{chemo,I}}(\boldsymbol{t})\right)}{\boldsymbol{\Delta x}} \cdot \boldsymbol{S} \cdot \frac{\boldsymbol{V}_{\mathbf{tumor}}(\boldsymbol{t})}{\boldsymbol{V}_{\boldsymbol{T},\boldsymbol{I}}(\boldsymbol{t})}}^{\textit{Delivery from plasma}} - \overbrace{\frac{\boldsymbol{D}_{\mathbf{chemo}}}{\mathbf{Len}^2} \cdot \boldsymbol{C}_{\mathbf{chemo},\boldsymbol{I}}(\boldsymbol{t})}^{\textit{Diffusion to canc⍰⍰r cells}} - \overbrace{\boldsymbol{\beta}_{\mathbf{chemo}} \cdot \boldsymbol{C}_{\mathbf{chemo},\boldsymbol{I}}(\boldsymbol{t})}^{\textit{Degradation}}, \qquad \boldsymbol{C}_{\mathbf{chemo},\boldsymbol{I}}(\mathbf{0}) = \mathbf{0} \qquad (\mathbf{S28})$$

The concentration of chemotherapy in the tumor interstitium $\left(\boldsymbol{C}_{\mathbf{chemo},\boldsymbol{I}}(\boldsymbol{t})\right)$ is influenced by three processes. The first one is the concentration gradient described in the second term Eq. S27. The second one is the diffusion of the chemotherapeutic agent to cancer cells, where $\boldsymbol{D}_{\mathbf{chemo}}$ is the diffusivity of chemotherapy. The third is the degradation of chemotherapy.

**Equation for chemotherapy concentration kinetics in cancer cells ($\boldsymbol{C}_{\mathbf{chemo},\boldsymbol{L}}(\boldsymbol{t})$):**

$$\frac{d\boldsymbol{C}_{\mathbf{chemo},\boldsymbol{L}}(\boldsymbol{t})}{d\boldsymbol{t}} = \overbrace{\frac{\boldsymbol{D}_{\mathbf{chemo}}}{\mathbf{Len}^2} \cdot \boldsymbol{C}_{\mathbf{chemo},\boldsymbol{I}}(\boldsymbol{t}) \cdot \frac{\boldsymbol{V}_{\boldsymbol{T},\boldsymbol{I}}(\boldsymbol{t})}{\boldsymbol{L}(\boldsymbol{t}) + \mathbf{M\Phi}(\boldsymbol{t}) + \mathbf{CD8}(\boldsymbol{t})}}^{\textit{Delivery from interstitium}} - \overbrace{\boldsymbol{\beta}_{\mathbf{chemo}} \cdot \boldsymbol{C}_{\mathbf{chemo},\boldsymbol{L}}(\boldsymbol{t})}^{\textit{Degradation}}, \qquad \boldsymbol{C}_{\mathbf{chemo},\boldsymbol{L}}(\mathbf{0}) = \mathbf{0} \qquad (\mathbf{S29})$$

The last equation of our model characterizes the rate of change of chemotherapy concentration inside cancer cells $\left(\boldsymbol{C}_{\mathbf{chemo},\boldsymbol{L}}(\boldsymbol{t})\right)$. The first term is the diffusion process that appeared in the second term of Eq. S28. The second term is chemotherapy degradation.

#### S2. Treatment response evaluation

TGI was assessed by comparing the simulated tumor growth under control and treatment scenarios, and was calculated as: $\mathbf{TGI}(\%) = (\mathbf{1} - \boldsymbol{D}_{\mathbf{treated}}/\boldsymbol{D}_{\mathbf{control}}) \cdot \mathbf{100}$, where $\boldsymbol{D}_{\mathbf{treated}}$ and $\boldsymbol{D}_{\mathbf{control}}$ represent tumor diameters under treatment and control conditions, respectively, at the end of treatment. A TGI value of 100% indicates complete tumor growth inhibition, while lower values correspond to varying degrees of inhibition.

To perform a clinically relevant assessment of treatment response, we monitored the diameter of simulated tumors following treatment initiation. Then, employing the RECIST 1.1 guidelines (3), we determined the time to progression (TTP), which is defined as the time from treatment initiation to when the criteria for progressive disease (PD) were met. As per RECIST 1.1, treatment response can be classified into one of four categories: 1) Complete Response (CR): disappearance of tumor; 2) Partial Response (PR): at least a 30% decrease in tumor diameter from the initiation of treatment; 3) PD: at least a 20% increase in tumor diameter, with reference to the smallest diameter recorded since the beginning of treatment, and an absolute increase of at least 5 mm; 4) Stable Disease (SD): neither a sufficient shrinkage to qualify for PR nor sufficient growth to qualify for PD, taking as reference the smallest diameter recorded since treatment initiation.

Thus, based on the TTP values for the entire patient cohort, Kaplan-Meier survival analysis was performed to estimate progression-free survival (PFS) probability over time. From this, the median PFS value was determined, which represents the time at which 50% of the virtual patient cohort has experienced PD. Median PFS thus served as a key measure of drug efficacy in our analysis. Additionally, hazard ratios were calculated using Cox Proportional Hazards model to estimate relative risk of PD occurring between treatment and control groups.

#### S3. Parameter sensitivity analysis

We performed global sensitivity analysis (GSA) and local sensitivity analysis (LSA) by perturbing specific parameters of interest to investigate the importance of the various parameters in causing tumor shrinkage under treatment with the various monotherapies.

For GSA, all relevant model parameters (25 for cisplatin and 28 for the other drugs, i.e., anti-miR-155 and immune checkpoint inhibitors; see **Figs. 3A, S10**) were concurrently perturbed over a range of ±50% of their baseline values, and TGI was calculated for each combination of parameters using model-based simulations. To explore the extensive multiparameter space while maintaining computational efficiency, we employed Latin Hypercube Sampling (LHS) (4-6). Thus, 10,000 parameter combinations were sampled, and multiple linear regression analysis (MLRA) was applied using the corresponding TGI estimates from model simulations. The MLRA regression coefficients served as sensitivity indices (SI) to quantify parameter sensitivity. To ensure a robust analysis, the process was repeated five times, resulting in a distribution of SI values for each parameter. One-way ANOVA and Tukey's test were then employed to rank the parameters based on their significance, with a higher SI value indicating a more pronounced impact on TGI.

Subsequently, we performed LSA on the top ten parameters derived from the GSA ranking for anti-miR-155. In LSA, each parameter was altered one at a time, while the remaining parameters were held constant at their baseline values. Each parameter was tested at 100 levels within the range of ±50% of its baseline value, and TGI was calculated to establish the qualitative relationship between individual parameter changes and TGI.

It is important to note that sensitivity analyses were conducted under a treatment regimen involving once every three weeks (Q3W) injections of 0.026 mg/kg anti-miR-155-loaded NPs for nine treatment cycles, starting at 124 weeks post-tumor initiation. The chosen dose of 0.026 mg/kg represents the human equivalent dose of the *in vivo* dose of anti-miR-155, as calculated through allometric scaling (7). We also conducted GSA individually for the standard-of-care drugs (cisplatin, atezolizumab, and pembrolizumab), using the same methodology as above, to identify drug-related parameters that were key to governing tumor response to these drugs, and thus fine-tune the identified parameter/s to improve the accuracy of the allometrically scaled model for clinically relevant predictions of PFS.

#### S4. Generation of virtual patient cohort

To generate a virtual patient cohort, we adapted the methodology of Allen et al. (8), where we characterized a virtual patient by a set of 23 biological/physiological model parameters. These parameters included $g_0^M$, $g_{\text{tran}}$, $\delta_M$, $\varepsilon_M$, $k_M$, $A_{M,L}$, $g_0^{\text{PDL1}}$, $\delta_{\text{PDL1}}$, $k_{\text{on,P}}$, $k_{\text{off,P}}$, $g_0^{\text{PD1}}$, $\delta_{\text{PD1}}$, $\gamma_0^{\text{M}\Phi}$, $\delta_{\text{M}\Phi}$, $\gamma_0^{\text{CD8}}$, $\delta_{\text{CD8}}$, $\delta_{\text{immun}}$, $\varepsilon$, $k_{L,\text{M}\Phi}$, $k_{L,\text{CD8}}$, $\gamma$, $\text{Len}$, and $\Delta x$; see **Table S1** for definitions, marked by **†**. For each parameter, we set ±25% of its baseline value (defined in **Table S1**) as the biologically feasible upper and lower bounds. We randomly sampled a parameter value from this defined range for each parameter (assuming uniform distributions) to generate a combination of 23 parameters that characterized a patient. Using these sampled values, we simulated tumor growth under control conditions for up to 136 weeks post-tumor inception with a single cell on day zero. Finally, we applied Simulated Annealing (an optimization algorithm; using a built-in MATLAB function known as *simulannealbnd*) to adjust the sampled parameter values, while staying within the ±25% bounds, to minimize the sum of squared distance between the predicted tumor size at 136 weeks and the closet size within the target range (i.e., 1–2.68 cm). Using an iterative approach we generated a primary cohort of 10,000 patients, out of which we randomly sampled 1,000 patients as the final virtual cohort used for clinically relevant simulations in our study.

As shown in **Fig. S9A**, the distribution of % change in the parameter values after optimization (compared to their respective baseline values) confirms to the imposed bounds of ±25%. Further, the corresponding distributions of tumor size of the primary cohort of patients ($N$ = 10,000) and the smaller cohort randomly sampled from the primary cohort for simulation experiments ($N$ = 1,000) are restricted within the intended range of 1–2.68 cm (**Fig. S9B,C**). These distributions qualitatively resemble the gamma distributions observed for tumor sizes of stage N0, M0 (i.e., localized) lung cancer in the SEER (Surveillance, Epidemiology and End Results) database (9). The above observations support the biological plausibility of our virtual patient cohort.

**Table S1. List of biological parameters of the model.**

| Parameter | Description | Units | Value | Ref. |
|---|---|---|---|---|
| **miR-155 related parameters** | | | | |
| $^{\dagger}g_0^M$ | mir-155 production rate | $\mathbf{pM \cdot wk^{-1}}$ | 15.4 | Fit |
| $^{\dagger}g_{\mathrm{tran}}$ | mir-155 transfer rate from TAMs to cancer cells via exosomes | $\mathbf{wk^{-1}}$ | 0.045 | Fit |
| $^{\dagger}\delta_M$ | mir-155 degradation rate | $\mathbf{wk^{-1}}$ | 11 | (10) |
| $^{\dagger}\varepsilon_M$ | Efficiency of mir-155 at suppressing PD-L1 | $\mathbf{pM^{-1}}$ | 9.17 | Fit |
| $^{\dagger}k_M$ | Michaelis constant for mir-155 effects on tumor growth and chemoresistance | $\mathbf{pM}$ | 0.26 | Fit |
| $^{\dagger}A_{M,L}$ | Stimulation factor for mir-155 effects on tumor growth and chemoresistance | - | 9.89 | Fit |
| **PD-L1 related parameters** | | | | |
| $^{\dagger}g_0^{\mathrm{PDL1}}$ | Production rate of PD-L1 | $\mathbf{pM \cdot wk^{-1}}$ | 99.49 (M)<br>12.9 (H) | Fit,<br>IS |
| $^{\dagger}\delta_{\mathrm{PDL1}}$ | Degradation rate constant of PD-L1 | $\mathbf{wk^{-1}}$ | 49.99 (M)<br>6.5 (H) | IS,<br>(11) |
| $^{\dagger}k_{\mathrm{on,P}}$ | Binding rate of PD-L1 to PD-1 | $\mathbf{pM^{-1}wk^{-1}}$ | 2.17728e-2 (M)<br>2.8e-3 (H) | (12),<br>IS |
| $k_{\mathrm{on,Ab1}}$ | Binding rate of PD-L1 to atezolizumab | $\mathbf{pM^{-1}wk^{-1}}$ | 4.1588 (M)<br>0.54 (H) | IS,<br>(13) |
| $^{\dagger}k_{\mathrm{off,P}}$ | Unbinding rate constant between PD-1 and PD-L1 | $\mathbf{wk^{-1}}$ | 9.072e4 (M)<br>1.1795e4 (H) | (12),<br>IS |
| $k_{\mathrm{off,Ab1}}$ | Unbinding rate of PD-L1 from atezolizumab | $\mathbf{wk^{-1}}$ | 8.3923e2 (M)<br>109.1 (H) | IS,<br>(13) |
| **PD-1 related parameters** | | | | |
| $^{\dagger}g_0^{\mathrm{PD1}}$ | Production rate of PD-1 | $\mathbf{pM \cdot wk^{-1}}$ | 98.2 (M)<br>12.77 (H) | Fit, IS |
| $^{\dagger}\delta_{\mathrm{PD1}}$ | Degradation rate constant of PD-1 | $\mathbf{wk^{-1}}$ | 97 (M)<br>12.6111 (H) | (14)<br>IS |
| $k_{\mathrm{on,Ab2}}$ | Binding rate of PD-1 to pembrolizumab | $\mathbf{pM^{-1}wk^{-1}}$ | 2.8191 (M)<br>0.3665 (H) | IS<br>(13) |
| $k_{\mathrm{off,Ab2}}$ | Unbinding rate of PD-1 from pembrolizumab | $\mathbf{wk^{-1}}$ | 1.1873e3 (M)<br>1.5436e2 (H) | IS<br>(13) |
| **TAM related parameters** | | | | |
| $^{\dagger}\gamma_0^{\mathrm{M\Phi}}$ | Maximum recruitment rate of TAMs | $\mathbf{cm^3 \cdot wk^{-1}}$ | 0.089 (study 1),<br>0.086 (study 2),<br>0.062 (study 3)<br>0.0103 (H) | Fit, IS |

| | | | | |
|---|---|---|---|---|
| $^{\dagger}\boldsymbol{\delta_{\mathrm{M\phi}}}$ | Death rate constant of TAMs | $\mathbf{wk^{-1}}$ | 0.16 (M)<br>0.0208 (H) | (15), IS |
| **CD8+ T-lymphocyte related parameters** | | | | |
| $^{\dagger}\boldsymbol{\gamma_{0}^{\mathrm{CD8}}}$ | Maximum infiltration rate of CD8$^{+}$ T cells | $\mathbf{cm^{3}\cdot wk^{-1}}$ | 0.104 (M)<br>0.0135 (H) | Fit, IS |
| $^{\dagger}\boldsymbol{\delta_{\mathrm{CD8}}}$ | Death rate constant of CD8$^{+}$ T cells | $\mathbf{wk^{-1}}$ | 1.6152 (M)<br>0.21 (H) | IS, (16) |
| $^{\dagger}\boldsymbol{\delta_{\mathrm{immun}}}$ | Maximal CD8$^{+}$ T cell-induced tumor death rate constant | $\mathbf{cm^{-3}\cdot wk^{-1}}$ | 23.1 (M)<br>3 (H) | Fit, IS |
| $^{\dagger}\boldsymbol{\varepsilon}$ | Efficiency of the PD-L1/PD-1 complex in suppressing CD8$^{+}$ T cell-induced tumor death | $\mathbf{pM^{-1}}$ | 9.8e6 (M)<br>4.49e9 (H) | Fit, IS |
| **Tumor related parameters** | | | | |
| $^{\dagger}\boldsymbol{k_{L,\mathrm{M\phi}}}$ | Michaelis constant for tumor effects on TAM recruitment | $\mathbf{cm^{3}}$ | 0.27 | Fit |
| $^{\dagger}\boldsymbol{k_{L,\mathrm{CD8}}}$ | Michaelis constant for tumor effects on CD8$^{+}$ T-cell infiltration | $\mathbf{cm^{3}}$ | 0.077 | Fit |
| $^{\dagger}\boldsymbol{\gamma}$ | Tumor growth rate | $\mathbf{wk^{-1}}$ | 0.087 (study 1),<br>0.21 (study 2),<br>0.1 (study 3),<br>0.0176 (H) | Fit, IS |
| $\boldsymbol{S}$ | Tumor microvascular surface area | cm$^{2}$/cm$^{3}$ | 566.6 | (17) |
| $\boldsymbol{\phi_{\mathrm{pore}}}$ | Diameter of tumor vessel wall pores | cm | 17e-5 | (6) |
| $^{\dagger}\mathbf{Len}$ | Characteristic length of intercapillary distance in tumor interstitium | $\mathbf{cm}$ | 0.01 | (18) |
| $^{\dagger}\boldsymbol{\Delta x}$ | Thickness of capillary wall | $\mathbf{cm}$ | 0.0005 | (19) |
| $\boldsymbol{f_{I}}$ | Tumor interstitial volume fraction | - | 27.5% | (1) |
| $\boldsymbol{f_{V}}$ | Tumor vascular volume fraction | - | 17% | (2) |

**Note:** In column 4, M and H notations denote the parameter values for mice and humans, respectively. Fit indicates parameter values obtained through data fitting and IS denotes the values obtained through interspecies scaling.

† denotes parameters that were used to generate the virtual patient cohort. Study 1, study 2, and study 3 denote *in vivo* datasets for anti-miR-155, atezolizumab, and pembrolizumab, respectively.

**Table S2. List of therapy related parameters of the model.**

| Parameter | Description | Units | Value | Ref. |
|---|---|---|---|---|
| **Anti-mir-155 related parameters** | | | | |
| $\mathbf{EC_{50,AM}}$ | Half maximal effective concentration of anti-mir-155 | $\mathbf{mg \cdot cm^{-3}}$ | 0.19 | Fit |
| $\boldsymbol{A_{\mathrm{AM},M}}$ | Stimulation factor for anti-mir-155 degradation effects on mir-155 | - | 127.8 | Fit |
| $\boldsymbol{\delta_{\mathrm{AM}}}$ | Decay rate of anti-mir-155 | $\mathbf{wk^{-1}}$ | 11 | (10) |
| **Immunotherapy related parameters** | | | | |
| $\mathbf{Dose_{Ab1}}$ | Dose of anti-PD-L1 antibody (atezolizumab) | $\mathbf{mg}$ | 0.2 (M)<br>1200 (H) | (20,21) |
| $\mathbf{Dose_{Ab2}}$ | Dose of anti-PD-1 antibody (pembrolizumab) | $\mathbf{mg}$ | 0.1 (M)<br>200 (H) | (22,23) |
| $\boldsymbol{k_{\mathrm{abs}}}$ | Systemic absorption rate of antibodies from peritoneal cavity | $\mathbf{wk^{-1}}$ | 16.6 (M) | Fit |
| $\boldsymbol{\phi_{\mathrm{Ab}}}$ | Antibody diameter | $\mathbf{cm}$ | 1e-6 | |
| $\boldsymbol{P_{\mathrm{Ab}}}$ | Tumor microvascular permeability of antibodies | $\mathbf{cm \cdot wk^{-1}}$ | 0.0727 | Calc. |
| $\mathbf{Cl_{Ab1}}$ | Clearance of atezolizumab | $\mathbf{mL \cdot wk^{-1}}$ | 3.07 (M)<br>1397 (H) | (24), IS |
| $\mathbf{Cl_{Ab2}}$ | Clearance of pembrolizumab | $\mathbf{mL \cdot wk^{-1}}$ | 3.38 (M)<br>1538 (H) | (25), IS |
| $\boldsymbol{V_{\mathrm{Ab1}}}$ | Volume of distribution of atezolizumab | $\mathbf{mL}$ | 1.97 (M)<br>6895 (H) | (24), IS |
| $\boldsymbol{V_{\mathrm{Ab2}}}$ | Volume of distribution of pembrolizumab | $\mathbf{mL}$ | 2.2 (M)<br>7700 (H) | (25), IS |
| $\boldsymbol{^*D_{\mathrm{Ab}}}$ | Diffusivity of antibodies in tumor interstitium | $\mathbf{cm^2 \cdot wk^{-1}}$ | 0.0784 | Calc. |
| $\boldsymbol{\delta_{\mathrm{Ab}}}$ | Antibody degradation rate | $\mathbf{wk^{-1}}$ | 2.4298 (M)<br>2.9157 (H; after scaling for atezolizumab)<br>1.142 (H; after scaling for pembrolizumab) | Fit |
| **Chemotherapy related parameters** | | | | |
| $\boldsymbol{\delta_{\mathrm{chemo}}}$ | Maximal chemotherapy-induced tumor death rate constant | $\mathbf{wk^{-1}}$ | 2.34 (M)<br>0.3053 (H; before scaling)<br>0.4335 (H; after scaling) | Fit, IS |
| $\mathbf{EC_{50,chemo}}$ | Half maximal effective concentration of cisplatin | $\mathbf{nmol \cdot mL^{-1}}$ | $\mathbf{70}$ | (26) |
| $\boldsymbol{^*D_{\mathrm{chemo}}}$ | Diffusivity of chemotherapy in tumor interstitium | $\mathbf{cm^2 \cdot wk^{-1}}$ | 0.4933 | Calc. |
| $\mathbf{Cl_{chemo}}$ | Clearance of cisplatin | $\mathbf{mL \cdot wk^{-1}}$ | 10014.5 (M)<br>4557000 (H) | (27), IS |
| $\boldsymbol{V_{\mathrm{chemo}}}$ | Volume of distribution of cisplatin | $\mathbf{mL}$ | 5.75 (M)<br>20125 (H) | (27), IS |
| $\mathbf{Dose_{chemo}}$ | Dose of chemotherapy | $\mathbf{mg}$ | 0.16 (M)<br>142.5 (H) | (7,21) |

| | | | | |
|---|---|---|---|---|
| $\boldsymbol{\beta}_{\mathbf{chemo}}$ | Chemotherapy degradation rate | $\mathbf{wk^{-1}}$ | 0.59 | Fit |
| $\boldsymbol{k}_{\mathbf{abs,chemo}}$ | Systemic absorption rate of chemotherapy from peritoneal cavity | $\mathbf{wk^{-1}}$ | 14.2 (M) | Fit |
| **Nanoparticle related parameters** | | | | |
| $\boldsymbol{\phi}_{\mathbf{NP}}$ | NP diameter | $\mathbf{cm}$ | 7e-6 | |
| ***$\boldsymbol{P}_{\mathbf{NP}}$ | Tumor microvascular permeability of NP | $\mathbf{cm \cdot wk^{-1}}$ | 0.0093 | Calc. |
| $\boldsymbol{\delta}_{\mathbf{NP}}$ | NP degradation rate | $\mathbf{wk^{-1}}$ | 0.46 | Fit |
| **$\boldsymbol{k}_{\mathbf{Cl}}$ | Clearance of NPs | $\mathbf{wk^{-1}}$ | 6.12 | Calc. |
| *$\boldsymbol{D}_{\mathbf{NP}}$ | Diffusivity of NPs in tumor interstitium | $\mathbf{cm^{2} \cdot wk^{-1}}$ | 0.0112 | |
| $\boldsymbol{k}_{\mathbf{rel}}$ | Release rate of anti-miR-155 from NPs | $\mathbf{wk^{-1}}$ | 1 | Fit |

**Note:** In column 4, M and H notations denote the parameter values for mice and humans, respectively. Fit indicates parameter values obtained through data fitting and IS denotes the values obtained through interspecies scaling.

***$\boldsymbol{P}_{\mathbf{NP}}$ is a function of $\boldsymbol{\phi}_{\mathbf{NP}}$ and $\boldsymbol{\phi}_{\mathbf{pore}}$, and $\boldsymbol{P}_{\mathbf{Ab}}$ is a function of $\boldsymbol{\phi}_{\mathbf{Ab}}$ and $\boldsymbol{\phi}_{\mathbf{pore}}$. (6)

**$\boldsymbol{k}_{\mathbf{Cl}}$ is calculated from the following relation (28,29): $\boldsymbol{k}_{\mathbf{Cl}} = \frac{\mathbf{ln(2)}}{\mathbf{0.11} \cdot e^{-1.33 \cdot \phi_{\mathbf{NP}}} - \mathbf{0.001} \cdot e^{-9.7 \cdot \phi_{\mathbf{NP}}}}$.

*Diffusivity is calculated from Stokes-Einstein relation.

**Table S3. Initial conditions of model variables.**

| Notation | Description | Units | Value | Ref. |
|---|---|---|---|---|
| $M_0$ | Baseline concentration of mir-155 | $\mathbf{pM}$ | 1 | |
| $L_0$ | Initial cancer cell volume | $\mathbf{mL}$ | $10^{-9}$ | (9) |
| $\mathbf{M\phi_0}$ | Initial TAM cell volume | $\mathbf{mL}$ | $10^{-9}$ | (9) |
| $\mathbf{CD8_0}$ | Initial $CD8^+$ T cell volume | $\mathbf{mL}$ | $10^{-9}$ | (9) |
| $C_0^{\mathrm{PD1}}$ | Baseline concentration of PD-1 | $\mathbf{pM}$ | 0.05 | |
| $C_0^{\mathrm{PDL1}}$ | Baseline concentration of PD-L1 | $\mathbf{pM}$ | 0.1 | |

### Supplementary Results

#### S1. Clinical model calibration

To assess the ability of the allometrically scaled model to predict clinical endpoints (progression free survival (PFS), as defined by RECIST 1.1) for standard-of-care drugs, we simulated the treatment of a virtual patient cohort (*N* = 1,000) with pembrolizumab, atezolizumab, and cisplatin monotherapies, and compared it to the results of Phase 3 clinical trials of these drugs in NSCLC patients (pembrolizumab: KEYNOTE-024 (23), atezolizumab: IMpower110 (21), and cisplatin: CATAPULT I (30)). In accordance with the reported treatment design of these trials, the simulations involved a Q3W administration of fixed doses: 200 mg for pembrolizumab, 1200 mg for atezolizumab, and 75 mg/$m^2$ for cisplatin.

Given possible discrepancy in drug-specific inter-species scaling of one or more model parameters, the initial predictions of PFS probability required adjustments to align well with clinical observations for these drugs. For this, we conducted global sensitivity analysis (GSA) individually for the three drugs to identify the key drug-related parameters that significantly influenced treatment response (**Fig. S10**). In the case of pembrolizumab and atezolizumab, $\boldsymbol{\delta}_{\mathbf{Ab}}$, representing the degradation rate of antibodies in the body, emerged as a critical drug-related parameter affecting treatment response (i.e., TGI); whereas for cisplatin, $\boldsymbol{\delta}_{\mathbf{chemo}}$, which denotes the chemotherapy-induced tumor death rate, played a pivotal role in determining treatment response.

To enhance the accuracy of our predictions, we thus performed a parameter sweep with respect to these parameters. This iterative process involved simulating the above treatment protocols while systematically varying the values of $\boldsymbol{\delta}_{\mathbf{Ab}}$ or $\boldsymbol{\delta}_{\mathbf{chemo}}$ (depending on the drug) to minimize the disparities between predicted and observed median PFS. This led to the identification of scaling factors, which were applied to scale $\boldsymbol{\delta}_{\mathbf{Ab}}$ and $\boldsymbol{\delta}_{\mathbf{chemo}}$. Specifically, $\boldsymbol{\delta}_{\mathbf{Ab}}$ was scaled by 0.47 for pembrolizumab and 1.2 for atezolizumab, and $\boldsymbol{\delta}_{\mathbf{chemo}}$ was scaled by 1.42 for cisplatin (**Table S2**). As a result, we are able to optimize the model predictions to better match clinical observations of PFS probability over time for these three drugs (**Fig. S11**). Thus, by fine-tuning the allometrically scaled values of drug-related parameters $\boldsymbol{\delta}_{\mathbf{Ab}}$ and $\boldsymbol{\delta}_{\mathbf{chemo}}$, this exercise aimed to improve the predictive accuracy of the model for clinical endpoints for standard-of-care drugs in drug combination studies.

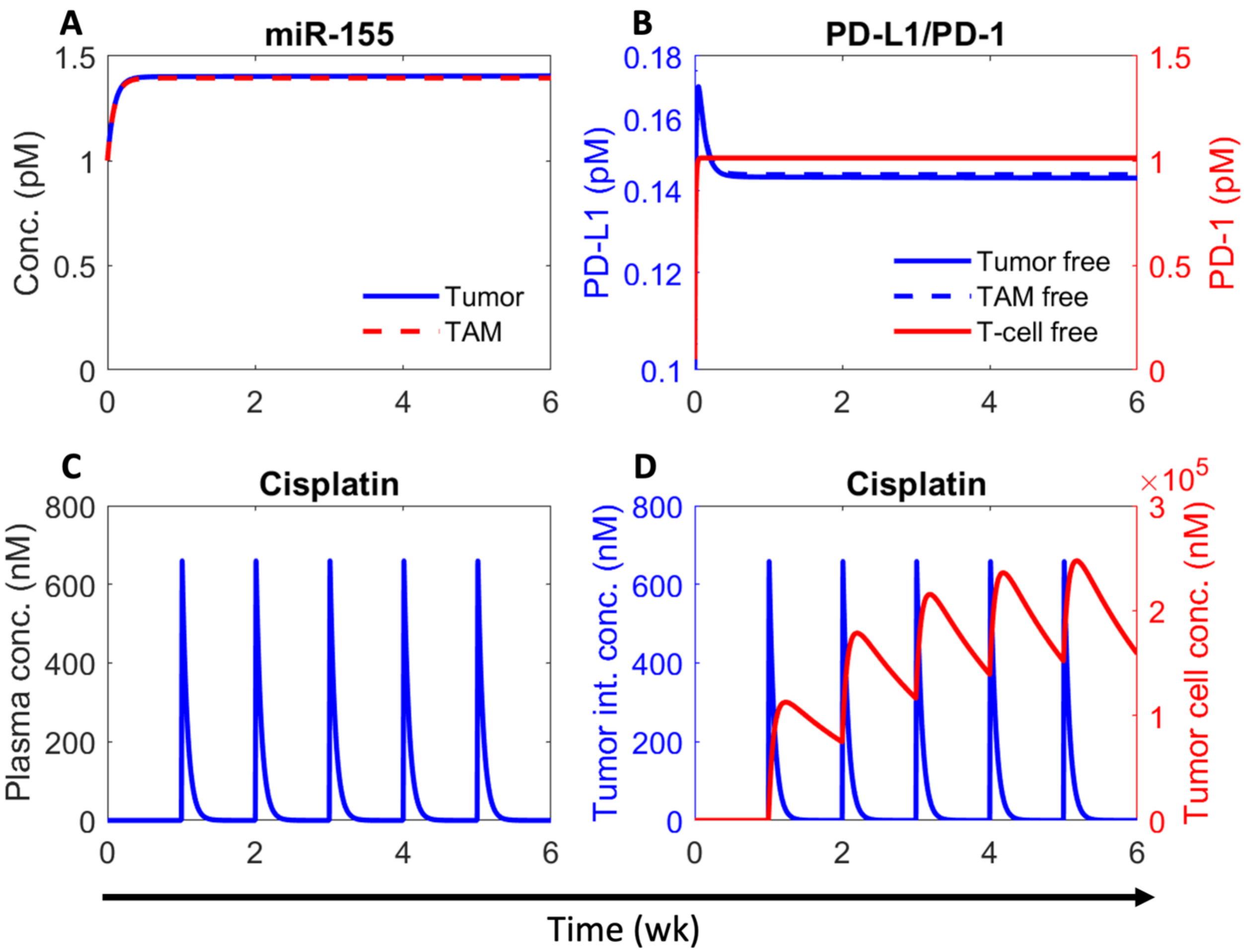


**Figure S1. Numerical solution of the model exhibiting kinetics of relevant variables under treatment with cisplatin.** A) Concentration kinetics of miR-155 in the tumor and TAMs. B) Concentration kinetics of unbound (i.e., free) PD-L1 on tumor and TAM (left y-axis) and unbound PD-1 on CD8+ T cells (right y-axis). C) Plasma concentration kinetics of cisplatin following once weekly injection at a dose of 8 mg/kg. D) Concentration kinetics of cisplatin in the tumor interstitium (left y-axis) and tumor cells (right y-axis). The corresponding tumor volumetric growth kinetics in shown in Figure 2E.

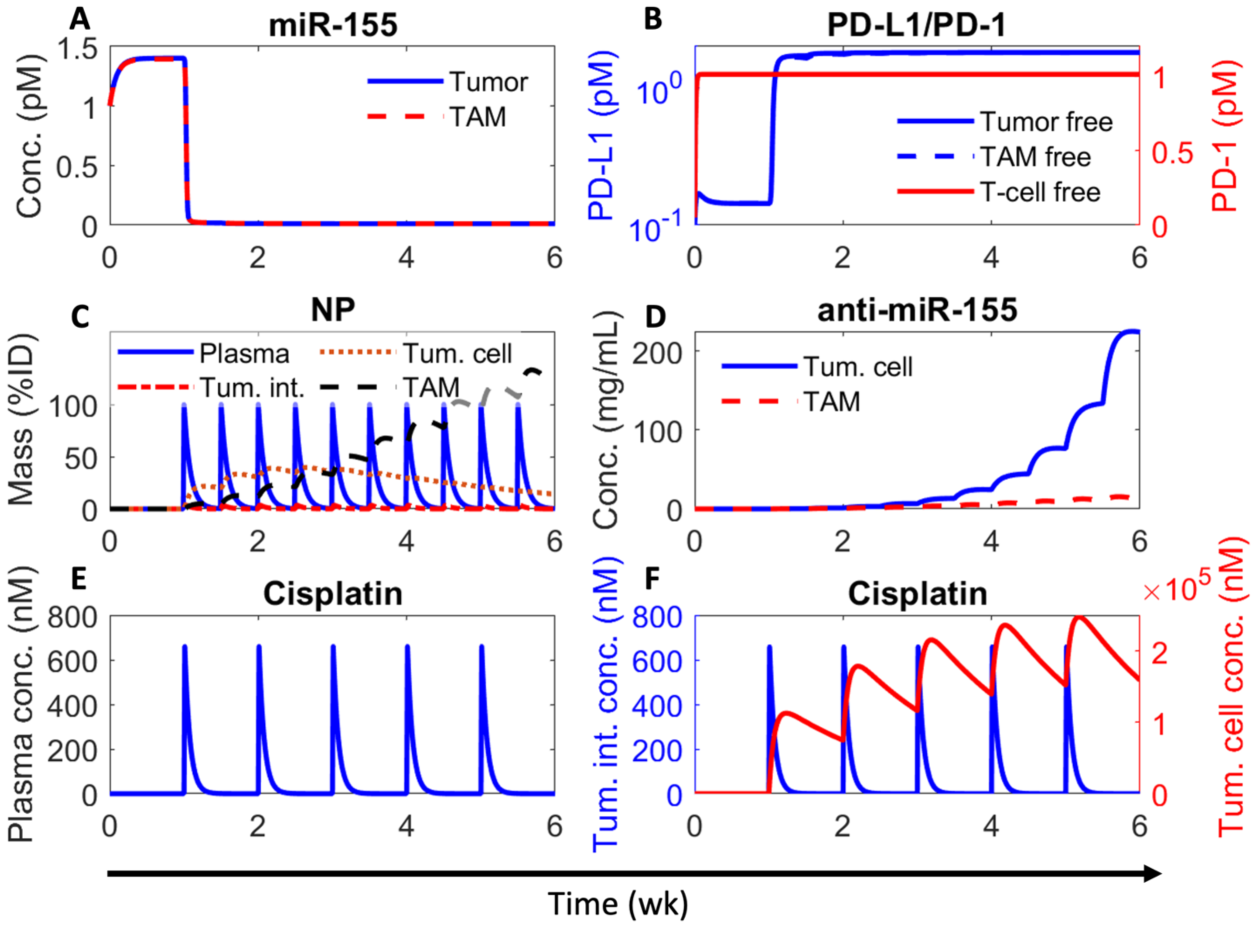


**Figure S2. Numerical solution of the model exhibiting kinetics of relevant variables under treatment with combination of cisplatin and nanoparticle-delivered anti-miR-155.** A) Concentration kinetics of miR-155 in the tumor and TAMs. B) Concentration kinetics of unbound (i.e., free) PD-L1 on tumor and TAM (left y-axis) and unbound PD-1 on CD8+ T cells (right y-axis). C) Mass kinetics of nanoparticles (NPs) in plasma, tumor interstitium, tumor cells, and TAMs following twice per weekly injection of NPs loaded with a dose of 4000 ng of anti-miR-155. %ID represents percent of injected dose. D) Concentration kinetics of anti-miR-155 in tumor cells and TAMs. E) Plasma concentration kinetics of cisplatin following once weekly injection at a dose of 8 mg/kg. F) Concentration kinetics of cisplatin in the tumor interstitium (left y-axis) and tumor cells (right y-axis). The corresponding tumor volumetric growth kinetics in shown in Figure 2E.

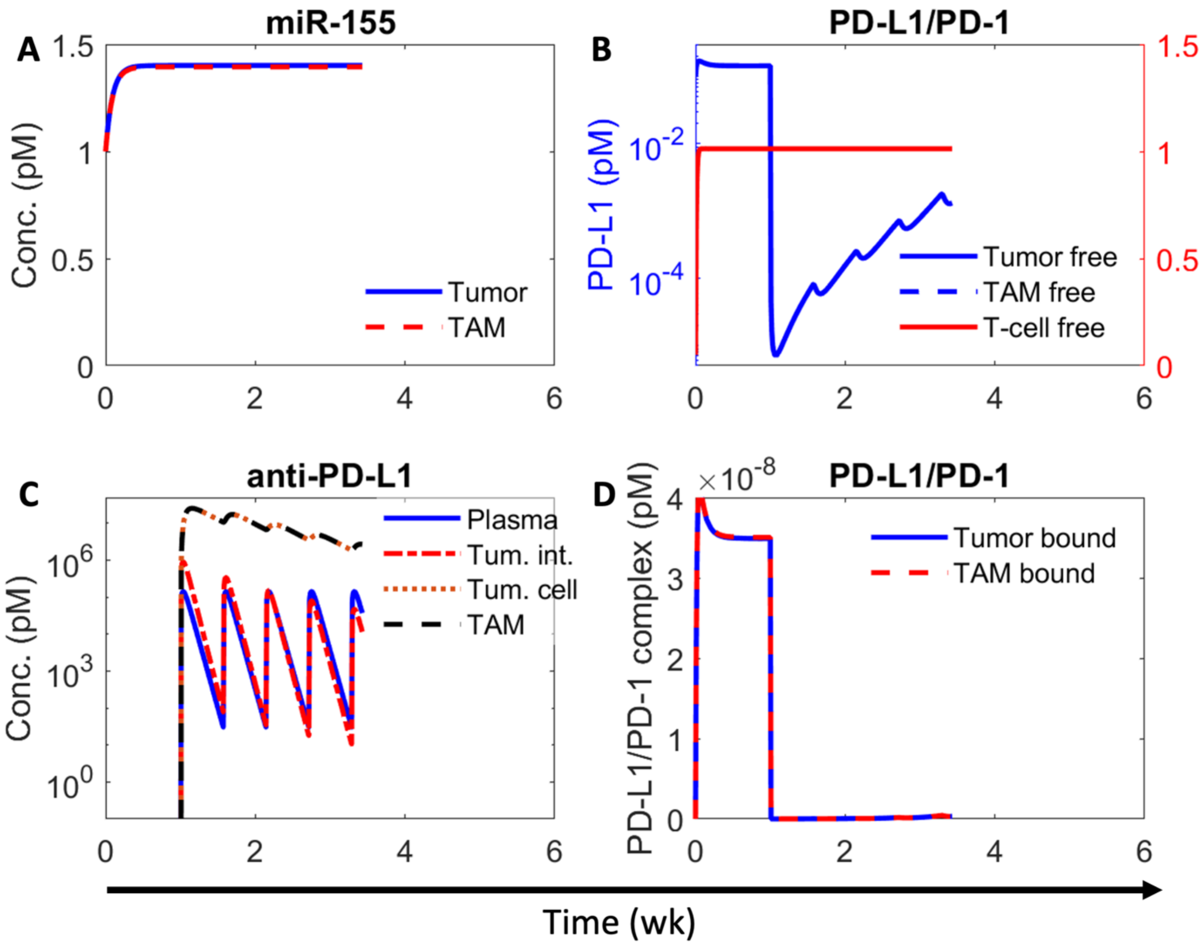


**Figure S3. Numerical solution of the model exhibiting kinetics of relevant variables under treatment with atezolizumab.** A) Concentration kinetics of miR-155 in the tumor and TAMs. B) Concentration kinetics of unbound (i.e., free) PD-L1 on tumor and TAM (left y-axis) and unbound PD-1 on CD8+ T cells (right y-axis). C) Concentration kinetics of anti-PD-L1 antibody atezolizumab in plasma, tumor interstitium, tumor cells, and TAMs following once every four days injection at a dose of 10 mg/kg. D) Concentration kinetics of PD-L1/PD-1 complex on tumor cells and TAMs. The corresponding tumor volumetric growth kinetics in shown in Figure 2F.

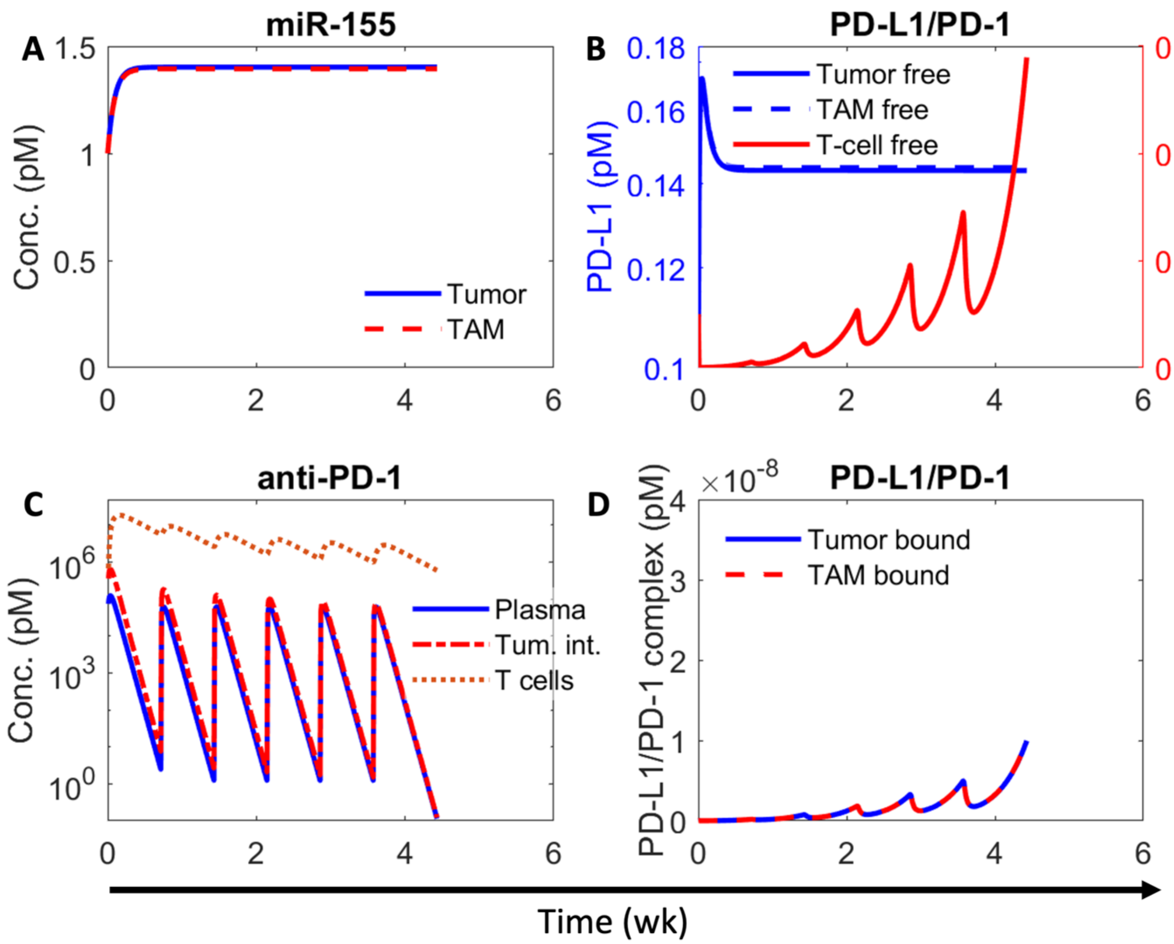


**Figure S4. Numerical solution of the model exhibiting kinetics of relevant variables under treatment with pembrolizumab.** A) Concentration kinetics of miR-155 in the tumor and TAMs. B) Concentration kinetics of unbound (i.e., free) PD-L1 on tumor and TAM (left y-axis) and unbound PD-1 on CD8+ T cells (right y-axis). C) Concentration kinetics of anti-PD-1 antibody pembrolizumab in plasma, tumor interstitium, and CD8+ T cells, following once every five days injection at a dose of 5 mg/kg. D) Concentration kinetics of PD-L1/PD-1 complex on tumor cells and TAMs. The corresponding tumor volumetric growth kinetics in shown in Figure 2G.

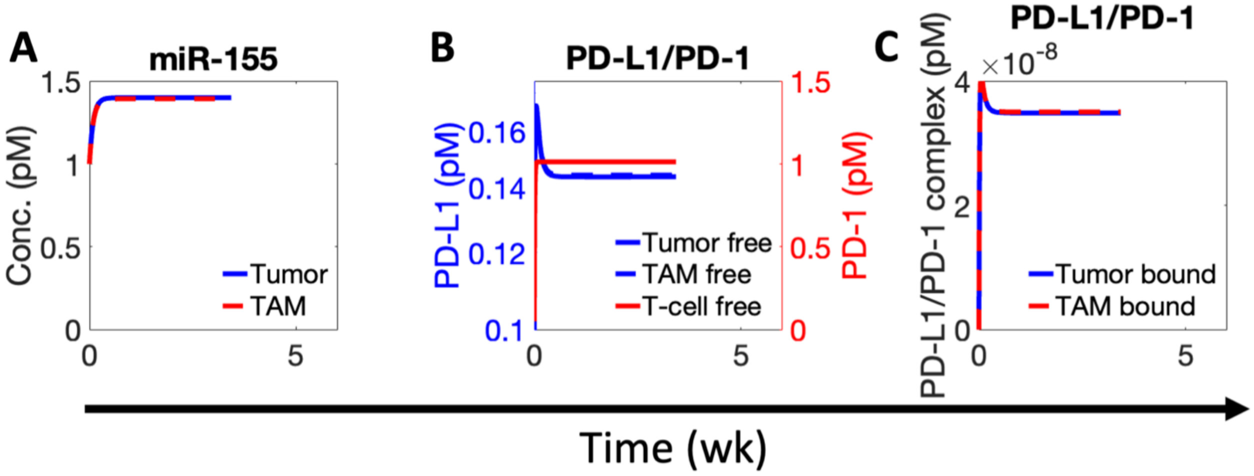


**Figure S5. Numerical solution of the model exhibiting kinetics of relevant variables under control conditions.** A) Concentration kinetics of miR-155 in the tumor and TAMs. B) Concentration kinetics of unbound (i.e., free) PD-L1 on tumor and TAM (left y-axis) and unbound PD-1 on CD8+ T cells (right y-axis). C) Concentration kinetics of PD-L1/PD-1 complex on tumor cells and TAMs. The corresponding tumor volumetric growth kinetics in shown in Figures 2F,G.

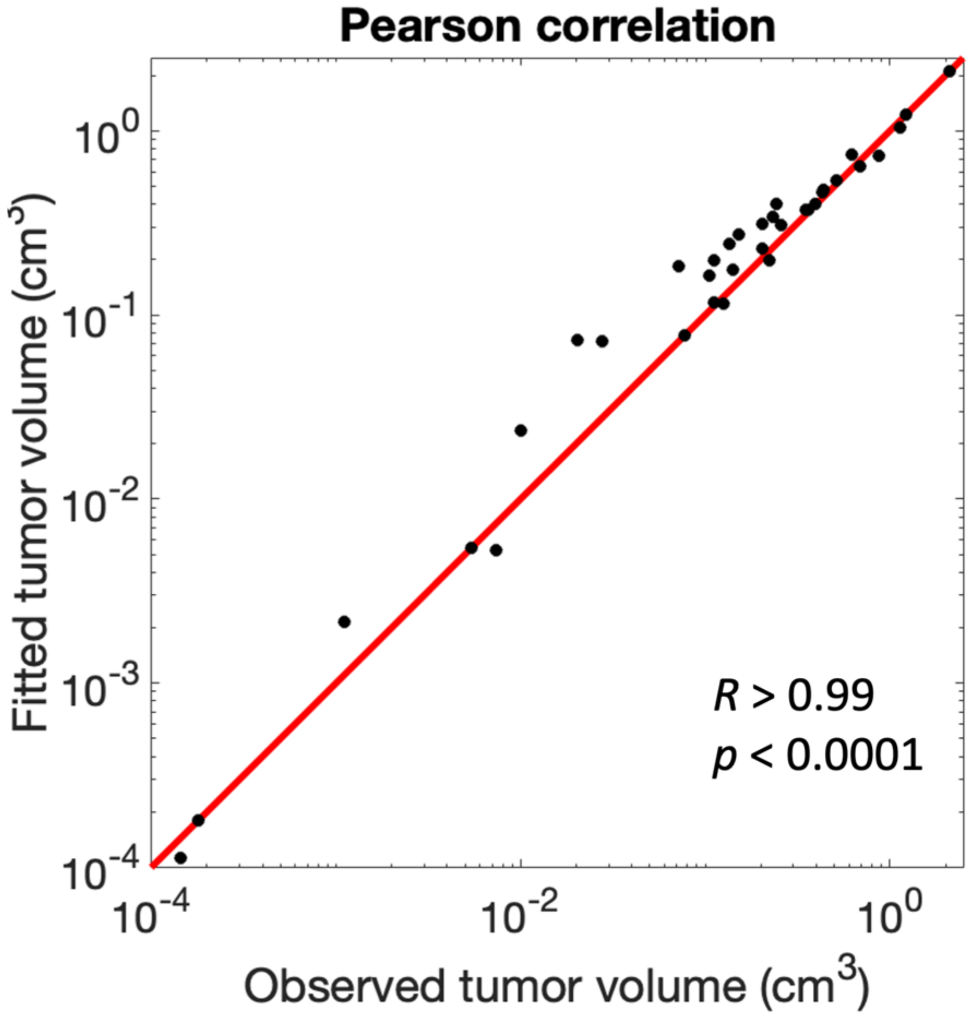


**Figure S6. Pearson correlation of model fits to in vivo data shown in Figures 2E-G.** Red line represents the y = x line.

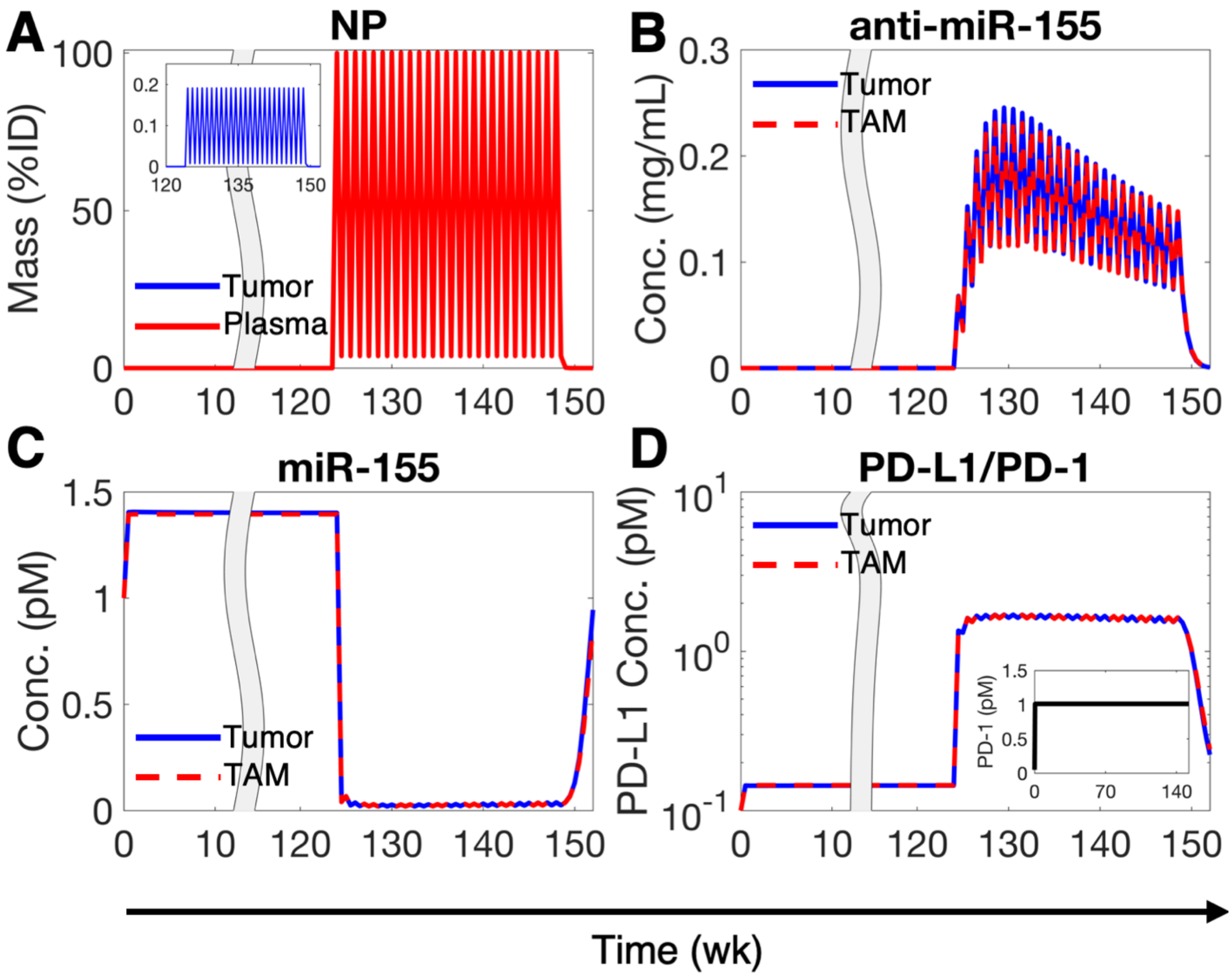


**Figure S7. Numerical solution of the allometrically scaled model showing key system variables following treatment with 0.026 mg/kg anti-miR-155 once weekly.** A) Mass kinetics of NPs in plasma and tumor interstitium (inset) following once weekly (QW) injection of NPs loaded with a dose of 0.026 mg/kg anti-miR-155. B) Concentration kinetics of NP-delivered anti-miR-155 in tumor cells and TAMs. C) Concentration kinetics of miR-155 in tumor cells and TAMs. D) Concentration kinetics of unbound PD-L1 on tumor cells and TAMs, and unbound PD-1 on CD8+ T cells (inset). The corresponding tumor growth kinetics is shown in Figure 2L.

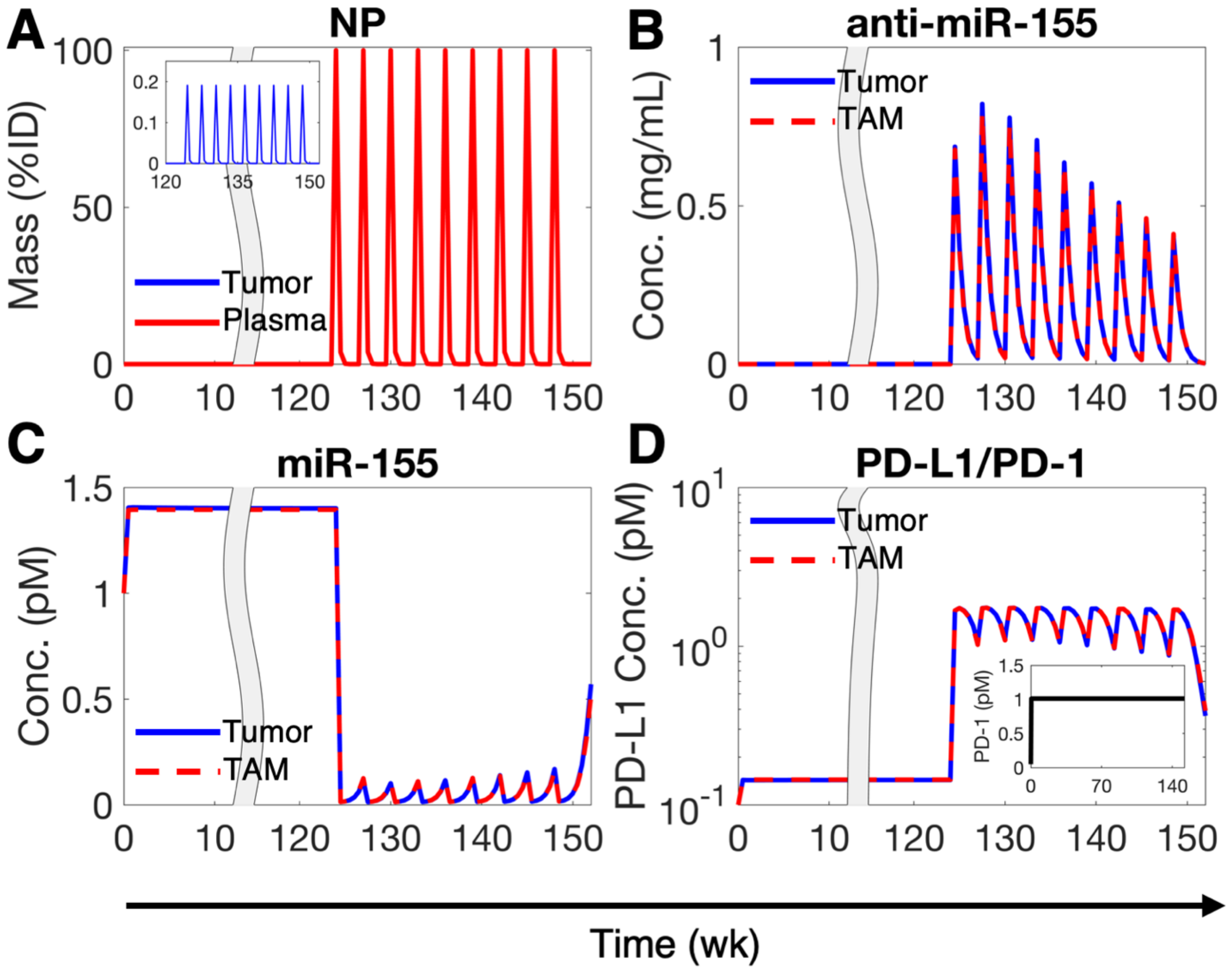


**Figure S8. Numerical solution of the allometrically scaled model showing key system variables following treatment with 0.26 mg/kg anti-miR-155 once in three weeks.** A) Mass kinetics of NPs in plasma and tumor interstitium (inset) following once in three weeks (Q3W) injection of NPs loaded with a dose of 0.26 mg/kg anti-miR-155. B) Concentration kinetics of NP-delivered anti-miR-155 in tumor cells and TAMs. C) Concentration kinetics of miR-155 in tumor cells and TAMs. D) Concentration kinetics of unbound PD-L1 on tumor cells and TAMs, and unbound PD-1 on CD8+ T cells (inset). The corresponding tumor growth kinetics is shown in Figure 2L.

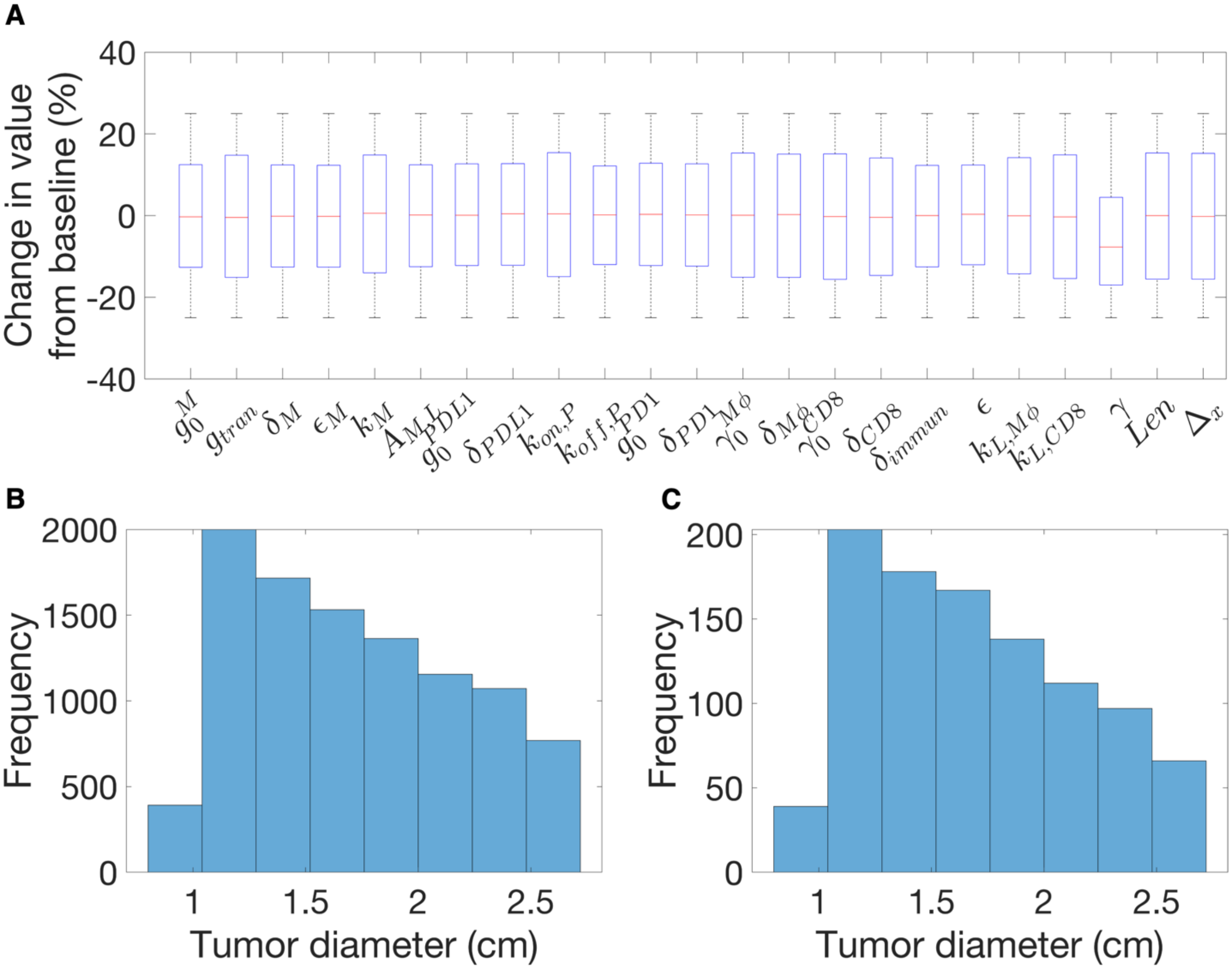


**Figure S9. Virtual patient cohorts.** A) Boxplots showing distribution of % change in parameter values from baseline after optimization through simulated annealing. B) Tumor size distribution for the primary cohort of 10,000 virtual patients, based on optimized parameters. C) Tumor size distribution of 1,000 randomly sampled virtual patients (without replacement from primary cohort) for clinical simulations.

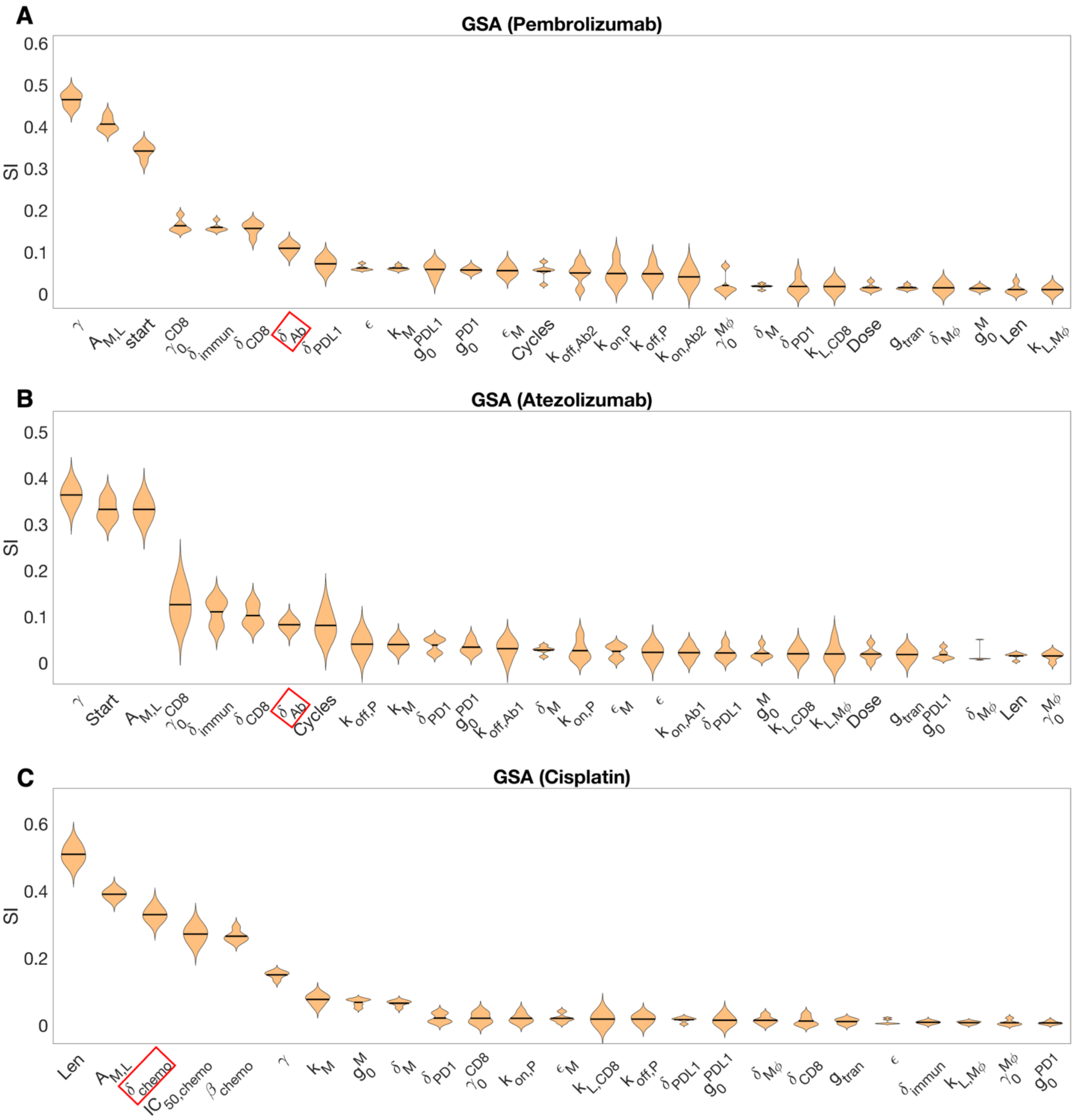


**Figure S10. Global sensitivity analysis.** Violin plots displaying the ranking of model parameters for their impact on A) pembrolizumab-, B) atezolizumab-, and C) cisplatin-induced tumor growth inhibition (TGI), as obtain from global sensitivity analysis (GSA). Multivariate linear regression analysis-based regression coefficients (labeled as sensitivity indices (SI)) were used to rank order the parameters using one-way ANOVA and Tukey's test. Parameters highlighted in red squares on the *x*-axis represent the highest-ranking drug-related parameters, chosen for optimizing model-based PFS predictions in Fig. S11.

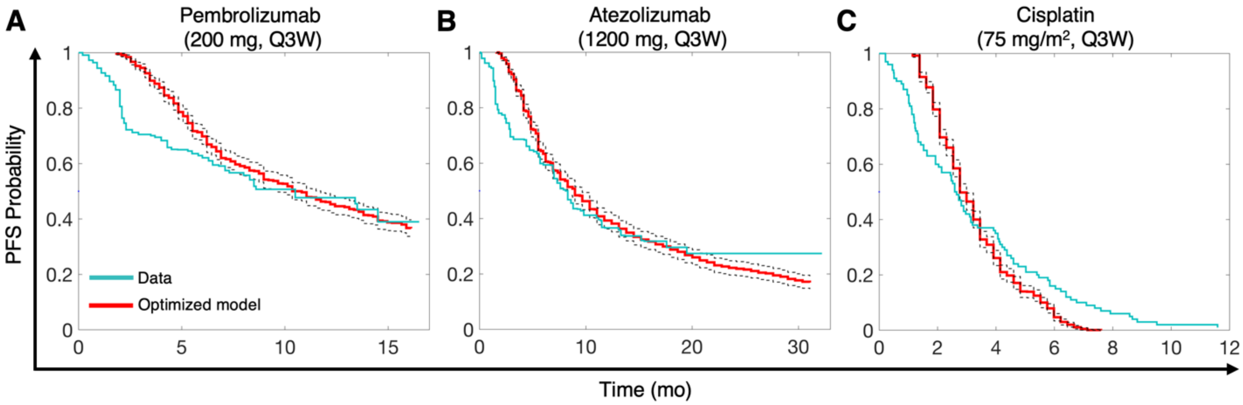


**Figure S11. Clinical model calibration.** A-C) Predictions of progression free survival (PFS) with the optimized model (red) for treatment of 1,000 virtual patients with A) 200 mg Q3W Pembrolizumab, B) 1,200 mg Q3W atezolizumab, and C) 75 mg/m$^2$ Q3W cisplatin in comparison with published clinical trial data (cyan) for the same dosage of the drugs.

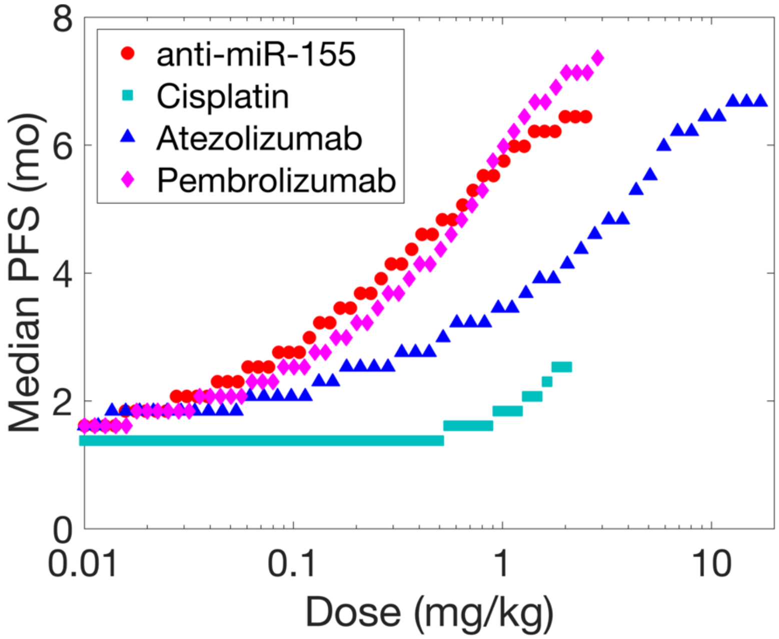


**Figure S12. Predictions of median PFS for a virtual patient cohort under once in three weeks monotherapy regimen.** Simulated treatments for a virtual patient cohort (N = 1,000) using uniformly spaced doses (log scale; 50 samples) following the Q3W regimen (9 treatment cycles, except 6 cycles for cisplatin) to predict median progression-free survival (PFS). Upper dose limits were set to clinically prescribed doses for standard-of-care drugs, with anti-miR-155 at 2.5 mg/kg.

**Table S4. Combination index (CI) values for combinations of anti-miR-155 and cisplatin.**

| Dose anti-miR-155 (mg/kg) | Dose Cisplatin (mg/kg) | TGI (%) | Median PFS (mo) | CI |
|---|---|---|---|---|
| 1.62 | 1.89 | 93.04 | 11.05 | 0.06 |
| 0.21 | 1.36 | 87.16 | 9.21 | 0.07 |
| 0.53 | 0.85 | 86.12 | 8.98 | 0.08 |
| 1.32 | 0.91 | 88.37 | 9.44 | 0.10 |
| 0.12 | 0.75 | 76.34 | 7.59 | 0.11 |
| 0.09 | 0.97 | 76.55 | 7.59 | 0.12 |
| 0.39 | 0.51 | 80.16 | 8.05 | 0.12 |
| 1.05 | 0.63 | 85.14 | 8.75 | 0.13 |
| 0.13 | 0.50 | 71.57 | 7.13 | 0.14 |
| 0.36 | 0.41 | 77.30 | 7.59 | 0.14 |
| 0.07 | 0.63 | 66.36 | 6.44 | 0.17 |
| 0.22 | 0.34 | 71.94 | 7.13 | 0.17 |
| 0.02 | 1.49 | 65.25 | 5.98 | 0.26 |
| 0.67 | 0.26 | 76.18 | 7.59 | 0.26 |
| 1.84 | 0.41 | 82.55 | 8.28 | 0.29 |
| 0.02 | 1.11 | 56.82 | 4.60 | 0.30 |
| 0.02 | 1.74 | 63.90 | 5.75 | 0.31 |
| 0.63 | 0.20 | 73.96 | 7.36 | 0.31 |
| 0.04 | 0.28 | 48.54 | 3.22 | 0.33 |
| 0.01 | 1.29 | 54.11 | 4.14 | 0.37 |
| 0.09 | 0.13 | 53.21 | 3.91 | 0.38 |
| 0.19 | 0.09 | 59.19 | 4.83 | 0.41 |
| 0.03 | 0.23 | 41.52 | 2.53 | 0.43 |
| 0.04 | 0.18 | 43.24 | 2.76 | 0.44 |
| 0.02 | 0.31 | 38.90 | 2.53 | 0.44 |
| 0.06 | 0.12 | 45.83 | 2.99 | 0.47 |
| 0.08 | 0.10 | 48.10 | 3.22 | 0.47 |
| 1.24 | 0.16 | 74.79 | 7.36 | 0.53 |
| 0.33 | 0.04 | 61.54 | 5.06 | 0.56 |
| 0.04 | 0.11 | 37.82 | 2.53 | 0.58 |
| 0.52 | 0.05 | 65.69 | 6.21 | 0.59 |
| 0.03 | 0.16 | 34.88 | 2.30 | 0.59 |
| 0.15 | 0.02 | 52.32 | 3.45 | 0.59 |
| 0.16 | 0.01 | 52.66 | 3.45 | 0.61 |
| 0.25 | 0.01 | 57.33 | 4.14 | 0.62 |
| 0.30 | 0.02 | 59.42 | 4.37 | 0.63 |
| 0.05 | 0.06 | 39.51 | 2.53 | 0.64 |
| 0.10 | 0.01 | 47.41 | 2.99 | 0.65 |
| 0.46 | 0.03 | 63.33 | 5.29 | 0.65 |
| 0.90 | 0.08 | 69.75 | 6.90 | 0.67 |
| 0.06 | 0.03 | 40.29 | 2.53 | 0.69 |
| 0.76 | 0.02 | 65.81 | 5.98 | 0.84 |
| 0.03 | 0.04 | 30.81 | 2.07 | 0.89 |
| 0.99 | 0.02 | 66.70 | 6.21 | 1.01 |
| 0.02 | 0.05 | 24.53 | 1.84 | 1.10 |
| 0.01 | 0.07 | 21.96 | 1.84 | 1.17 |
| 2.12 | 0.06 | 71.67 | 7.13 | 1.27 |
| 1.48 | 0.01 | 67.73 | 6.44 | 1.35 |
| 2.39 | 0.04 | 71.00 | 6.90 | 1.54 |
| 0.01 | 0.02 | 18.06 | 1.61 | 1.65 |

**Table S5. CI values for combinations of anti-miR-155 and atezolizumab.**

| Dose anti-miR-155 (mg/kg) | Dose Atezolizumab (mg/kg) | TGI (%) | Median PFS (mo) | CI |
|---|---|---|---|---|
| 0.28 | 4.67 | 81.41 | 8.28 | 0.09 |
| 0.23 | 10.11 | 83.51 | 8.75 | 0.09 |
| 0.43 | 7.02 | 83.65 | 8.75 | 0.10 |
| 0.19 | 8.99 | 82.69 | 8.52 | 0.10 |
| 0.21 | 15.94 | 84.59 | 8.98 | 0.10 |
| 0.25 | 1.69 | 76.86 | 7.82 | 0.12 |
| 0.16 | 1.48 | 74.14 | 7.59 | 0.12 |
| 0.45 | 2.03 | 79.49 | 8.05 | 0.13 |
| 0.93 | 6.39 | 84.44 | 8.75 | 0.13 |
| 0.07 | 13.27 | 81.20 | 8.52 | 0.15 |
| 1.17 | 5.70 | 84.38 | 8.75 | 0.16 |
| 0.73 | 2.16 | 80.74 | 8.28 | 0.16 |
| 0.05 | 4.10 | 73.84 | 7.59 | 0.17 |
| 0.05 | 1.16 | 65.17 | 6.44 | 0.19 |
| 0.06 | 0.47 | 60.07 | 5.29 | 0.21 |
| 0.54 | 0.74 | 75.87 | 7.59 | 0.21 |
| 1.57 | 3.68 | 83.44 | 8.75 | 0.22 |
| 0.11 | 0.26 | 61.95 | 5.52 | 0.22 |
| 0.17 | 0.27 | 65.37 | 6.21 | 0.22 |
| 0.03 | 3.18 | 69.76 | 7.13 | 0.23 |
| 0.13 | 0.22 | 62.66 | 5.52 | 0.23 |
| 0.04 | 0.31 | 54.16 | 3.91 | 0.27 |
| 0.03 | 0.53 | 55.93 | 4.14 | 0.28 |
| 0.06 | 0.10 | 51.60 | 3.45 | 0.31 |
| 0.07 | 0.08 | 52.37 | 3.68 | 0.32 |
| 0.02 | 0.99 | 57.81 | 4.83 | 0.33 |
| 0.02 | 2.64 | 65.12 | 6.44 | 0.34 |
| 0.02 | 0.38 | 50.67 | 3.45 | 0.35 |
| 0.04 | 0.08 | 46.03 | 2.99 | 0.36 |
| 0.12 | 0.04 | 55.28 | 3.91 | 0.37 |
| 0.03 | 0.15 | 45.78 | 3.22 | 0.37 |
| 1.15 | 0.61 | 76.97 | 7.82 | 0.38 |
| 0.09 | 0.03 | 52.02 | 3.45 | 0.38 |
| 0.02 | 0.13 | 43.68 | 2.76 | 0.39 |
| 0.01 | 11.24 | 73.80 | 7.59 | 0.41 |
| 0.09 | 0.01 | 50.27 | 3.22 | 0.44 |
| 0.01 | 1.28 | 56.85 | 4.60 | 0.45 |
| 1.88 | 0.78 | 78.62 | 8.05 | 0.50 |
| 0.32 | 0.02 | 62.43 | 5.06 | 0.50 |
| 0.87 | 0.17 | 72.07 | 7.13 | 0.51 |
| 0.37 | 0.02 | 63.51 | 5.29 | 0.51 |
| 0.01 | 0.12 | 38.43 | 2.53 | 0.53 |
| 0.01 | 0.04 | 33.47 | 2.30 | 0.53 |
| 0.48 | 0.02 | 65.26 | 5.98 | 0.56 |
| 0.76 | 0.06 | 68.87 | 6.67 | 0.62 |
| 0.66 | 0.03 | 67.41 | 6.44 | 0.62 |
| 1.30 | 0.01 | 68.87 | 6.67 | 1.05 |
| 1.71 | 0.06 | 71.43 | 6.90 | 1.05 |
| 2.10 | 0.02 | 70.20 | 6.67 | 1.47 |
| 2.44 | 0.03 | 71.01 | 6.90 | 1.57 |

**Table S6. CI values for combinations of anti-miR-155 and pembrolizumab.**

| Dose anti-miR-155 (mg/kg) | Dose Pembrolizumab (mg/kg) | TGI (%) | Median PFS (mo) | CI |
|---|---|---|---|---|
| 0.47 | 1.36 | 81.29 | 8.52 | 0.34 |
| 0.38 | 0.84 | 77.69 | 8.05 | 0.34 |
| 0.51 | 0.80 | 78.18 | 8.05 | 0.35 |
| 0.29 | 1.52 | 80.98 | 8.52 | 0.35 |
| 0.39 | 0.46 | 73.79 | 7.36 | 0.36 |
| 0.70 | 0.71 | 78.20 | 8.05 | 0.38 |
| 0.14 | 0.56 | 70.76 | 7.13 | 0.38 |
| 0.20 | 0.27 | 67.13 | 6.44 | 0.38 |
| 1.31 | 2.04 | 84.80 | 8.98 | 0.39 |
| 0.13 | 0.39 | 67.29 | 6.67 | 0.39 |
| 0.12 | 1.08 | 75.73 | 7.82 | 0.41 |
| 0.17 | 2.34 | 82.60 | 8.75 | 0.41 |
| 0.09 | 0.47 | 66.79 | 6.67 | 0.42 |
| 0.11 | 0.17 | 59.12 | 4.60 | 0.43 |
| 0.09 | 0.27 | 61.59 | 5.52 | 0.43 |
| 0.07 | 0.31 | 61.08 | 5.52 | 0.46 |
| 0.06 | 0.21 | 56.28 | 4.37 | 0.48 |
| 0.24 | 0.06 | 61.29 | 4.83 | 0.48 |
| 0.18 | 0.04 | 57.31 | 4.14 | 0.50 |
| 0.06 | 1.87 | 77.97 | 8.28 | 0.52 |
| 2.02 | 0.97 | 81.55 | 8.52 | 0.52 |
| 0.04 | 0.11 | 47.96 | 3.22 | 0.54 |
| 0.04 | 1.22 | 73.11 | 7.59 | 0.54 |
| 0.08 | 0.03 | 47.71 | 3.22 | 0.55 |
| 0.25 | 0.02 | 59.02 | 4.37 | 0.56 |
| 0.33 | 0.04 | 62.21 | 4.83 | 0.56 |
| 0.91 | 0.22 | 72.68 | 7.13 | 0.58 |
| 0.03 | 1.64 | 74.80 | 7.82 | 0.61 |
| 0.03 | 0.07 | 41.21 | 2.76 | 0.62 |
| 0.76 | 0.12 | 69.92 | 6.67 | 0.62 |
| 0.02 | 0.15 | 45.64 | 3.22 | 0.63 |
| 0.02 | 0.59 | 61.78 | 5.75 | 0.64 |
| 0.02 | 0.37 | 54.61 | 4.60 | 0.68 |
| 0.05 | 0.01 | 38.78 | 2.53 | 0.70 |
| 0.56 | 0.03 | 64.95 | 5.52 | 0.70 |
| 0.03 | 0.02 | 36.28 | 2.53 | 0.71 |
| 0.02 | 0.06 | 35.58 | 2.53 | 0.72 |
| 0.02 | 0.08 | 36.40 | 2.53 | 0.73 |
| 0.01 | 0.11 | 37.27 | 2.76 | 0.77 |
| 0.61 | 0.01 | 64.43 | 5.52 | 0.79 |
| 0.01 | 2.57 | 76.73 | 8.28 | 0.79 |
| 0.03 | 0.02 | 32.12 | 2.30 | 0.80 |
| 0.01 | 0.03 | 28.12 | 2.07 | 0.87 |
| 1.09 | 0.05 | 68.78 | 6.44 | 0.92 |
| 0.95 | 0.02 | 66.81 | 6.21 | 0.96 |
| 1.16 | 0.02 | 67.77 | 6.44 | 1.07 |
| 1.48 | 0.05 | 69.85 | 6.67 | 1.11 |
| 2.43 | 0.19 | 74.25 | 7.36 | 1.15 |
| 1.89 | 0.09 | 71.50 | 6.90 | 1.19 |
| 1.71 | 0.01 | 68.54 | 6.44 | 1.44 |

**Table S7. CI values for combinations of anti-miR-155, cisplatin, and atezolizumab.**

| Dose anti-miR-155 (mg/kg) | Dose Cisplatin (mg/kg) | Dose Atezolizumab (mg/kg) | TGI (%) | Median PFS (mo) | CI |
|---|---|---|---|---|---|
| 0.17 | 1.15 | 7.22 | 93.42 | 11.28 | 0.03 |
| 1.34 | 1.36 | 3.12 | 94.82 | 12.20 | 0.03 |
| 1.84 | 0.68 | 5.09 | 92.97 | 11.05 | 0.04 |
| 0.33 | 0.92 | 0.26 | 89.73 | 9.90 | 0.04 |
| 1.10 | 0.36 | 11.45 | 91.36 | 10.59 | 0.04 |
| 0.22 | 1.21 | 0.18 | 90.14 | 10.13 | 0.05 |
| 0.89 | 0.61 | 0.81 | 90.29 | 10.13 | 0.05 |
| 0.14 | 0.21 | 6.46 | 85.56 | 9.21 | 0.05 |
| 0.37 | 0.43 | 0.51 | 85.80 | 8.98 | 0.05 |
| 0.06 | 0.38 | 5.56 | 84.53 | 8.98 | 0.06 |
| 0.47 | 0.85 | 0.06 | 88.06 | 9.44 | 0.06 |
| 0.27 | 0.99 | 0.02 | 86.90 | 9.21 | 0.06 |
| 2.15 | 1.80 | 0.01 | 93.44 | 11.28 | 0.06 |
| 0.15 | 0.78 | 0.02 | 81.57 | 8.28 | 0.08 |
| 0.18 | 0.47 | 0.06 | 78.57 | 7.82 | 0.08 |
| 0.12 | 0.04 | 4.40 | 79.90 | 8.28 | 0.09 |
| 0.28 | 0.04 | 2.17 | 79.96 | 8.28 | 0.09 |
| 0.98 | 0.06 | 8.56 | 86.88 | 9.21 | 0.10 |
| 1.26 | 0.13 | 3.56 | 86.74 | 9.21 | 0.10 |
| 0.69 | 0.08 | 2.08 | 83.30 | 8.52 | 0.11 |
| 0.05 | 0.12 | 1.25 | 71.52 | 7.36 | 0.11 |
| 0.05 | 0.18 | 0.61 | 68.64 | 6.90 | 0.11 |
| 1.56 | 0.07 | 10.62 | 87.80 | 9.44 | 0.12 |
| 0.08 | 0.33 | 0.10 | 68.84 | 6.67 | 0.12 |
| 0.04 | 0.16 | 2.64 | 74.65 | 7.82 | 0.12 |
| 0.09 | 0.03 | 1.12 | 70.94 | 7.13 | 0.13 |
| 0.01 | 1.54 | 0.47 | 78.20 | 7.82 | 0.14 |
| 0.51 | 0.27 | 0.08 | 79.31 | 7.82 | 0.14 |
| 0.08 | 0.17 | 0.13 | 64.49 | 6.21 | 0.15 |
| 0.03 | 1.97 | 0.04 | 79.72 | 8.05 | 0.15 |
| 0.03 | 0.55 | 0.05 | 61.12 | 5.06 | 0.16 |
| 0.02 | 0.27 | 13.43 | 80.52 | 8.52 | 0.18 |
| 0.10 | 0.01 | 0.27 | 62.63 | 5.52 | 0.20 |
| 0.04 | 0.02 | 15.91 | 80.28 | 8.28 | 0.21 |
| 0.39 | 0.03 | 0.36 | 73.25 | 7.36 | 0.21 |
| 0.04 | 0.02 | 0.73 | 61.15 | 5.52 | 0.21 |
| 0.07 | 0.02 | 0.20 | 57.60 | 4.60 | 0.23 |
| 0.24 | 0.05 | 0.12 | 67.47 | 6.44 | 0.25 |
| 0.02 | 0.09 | 0.35 | 54.39 | 4.14 | 0.25 |
| 0.01 | 0.11 | 1.83 | 63.77 | 6.21 | 0.29 |
| 0.02 | 0.03 | 1.47 | 60.94 | 5.52 | 0.33 |
| 0.02 | 0.10 | 0.04 | 40.98 | 2.53 | 0.34 |
| 0.02 | 0.01 | 0.96 | 57.20 | 4.60 | 0.35 |
| 1.67 | 0.22 | 0.03 | 79.78 | 8.05 | 0.38 |
| 0.03 | 0.06 | 0.01 | 41.77 | 2.53 | 0.39 |
| 0.57 | 0.01 | 0.08 | 69.09 | 6.67 | 0.46 |
| 0.01 | 0.01 | 0.17 | 40.14 | 2.76 | 0.53 |
| 0.61 | 0.02 | 0.02 | 67.54 | 6.44 | 0.57 |
| 0.77 | 0.03 | 0.01 | 69.12 | 6.67 | 0.61 |
| 2.33 | 0.05 | 0.03 | 74.01 | 7.36 | 1.07 |

**Table S8. CI values for combinations of anti-miR-155, cisplatin, and pembrolizumab.**

| Dose anti-miR-155 (mg/kg) | Dose Cisplatin (mg/kg) | Dose Pembrolizumab (mg/kg) | TGI (%) | Median PFS (mo) | CI |
|---|---|---|---|---|---|
| 1.24 | 1.75 | 0.88 | 95.52 | 12.66 | 0.04 |
| 0.29 | 1.09 | 0.02 | 87.38 | 9.21 | 0.07 |
| 0.13 | 1.42 | 0.14 | 88.97 | 9.90 | 0.07 |
| 0.43 | 0.99 | 2.25 | 94.12 | 11.74 | 0.07 |
| 0.19 | 0.68 | 0.19 | 84.83 | 8.98 | 0.08 |
| 0.18 | 0.89 | 0.03 | 83.29 | 8.52 | 0.08 |
| 0.83 | 0.54 | 0.06 | 85.32 | 8.75 | 0.11 |
| 0.10 | 0.78 | 0.02 | 76.49 | 7.59 | 0.11 |
| 0.04 | 1.61 | 0.09 | 82.36 | 8.75 | 0.12 |
| 0.72 | 0.43 | 0.01 | 81.51 | 8.05 | 0.15 |
| 0.38 | 0.22 | 0.17 | 78.00 | 7.82 | 0.17 |
| 0.05 | 0.50 | 0.06 | 66.45 | 6.44 | 0.18 |
| 0.03 | 1.23 | 0.01 | 70.38 | 6.90 | 0.18 |
| 0.11 | 0.27 | 0.05 | 66.30 | 6.44 | 0.19 |
| 0.20 | 0.16 | 0.46 | 77.88 | 8.05 | 0.20 |
| 0.14 | 0.19 | 0.28 | 73.63 | 7.36 | 0.20 |
| 0.02 | 2.01 | 0.03 | 73.40 | 6.90 | 0.24 |
| 0.02 | 0.85 | 0.78 | 79.88 | 8.52 | 0.24 |
| 0.24 | 0.15 | 0.03 | 67.96 | 6.67 | 0.25 |
| 0.35 | 0.07 | 1.16 | 82.25 | 8.75 | 0.26 |
| 0.02 | 0.59 | 0.40 | 72.29 | 7.13 | 0.26 |
| 1.06 | 0.08 | 1.70 | 86.17 | 9.44 | 0.26 |
| 0.03 | 0.37 | 0.21 | 63.37 | 5.98 | 0.29 |
| 0.03 | 0.31 | 0.07 | 52.47 | 3.91 | 0.32 |
| 0.12 | 0.11 | 0.02 | 58.28 | 4.83 | 0.32 |
| 0.33 | 0.03 | 2.28 | 84.58 | 9.21 | 0.33 |
| 0.08 | 0.14 | 2.76 | 85.17 | 9.44 | 0.34 |
| 0.25 | 0.02 | 0.25 | 69.09 | 6.67 | 0.35 |
| 0.49 | 0.06 | 0.13 | 71.96 | 7.13 | 0.35 |
| 0.08 | 0.05 | 1.33 | 77.57 | 8.28 | 0.40 |
| 0.04 | 0.12 | 0.70 | 70.29 | 7.36 | 0.41 |
| 0.02 | 0.27 | 0.01 | 38.97 | 2.53 | 0.43 |
| 0.05 | 0.05 | 0.95 | 73.10 | 7.59 | 0.43 |
| 0.07 | 0.01 | 0.65 | 69.16 | 7.13 | 0.44 |
| 0.01 | 0.38 | 1.14 | 75.30 | 7.82 | 0.44 |
| 1.67 | 0.09 | 0.31 | 79.73 | 8.05 | 0.44 |
| 1.46 | 0.03 | 0.56 | 79.81 | 8.05 | 0.45 |
| 2.46 | 0.02 | 1.53 | 84.67 | 8.98 | 0.45 |
| 0.56 | 0.02 | 0.11 | 69.71 | 6.67 | 0.47 |
| 0.65 | 0.06 | 0.04 | 69.99 | 6.67 | 0.49 |
| 1.86 | 0.04 | 0.50 | 80.02 | 8.05 | 0.51 |
| 0.01 | 0.21 | 0.04 | 36.97 | 2.53 | 0.51 |
| 2.06 | 0.10 | 0.23 | 79.64 | 8.05 | 0.52 |
| 0.04 | 0.04 | 0.02 | 40.16 | 2.53 | 0.58 |
| 0.02 | 0.02 | 0.37 | 56.53 | 4.83 | 0.60 |
| 0.06 | 0.01 | 0.01 | 43.27 | 2.76 | 0.61 |
| 0.91 | 0.02 | 0.08 | 70.66 | 6.90 | 0.65 |
| 0.01 | 0.02 | 1.86 | 74.16 | 7.82 | 0.72 |
| 1.30 | 0.03 | 0.11 | 72.78 | 7.13 | 0.74 |
| 0.94 | 0.01 | 0.05 | 68.98 | 6.67 | 0.78 |
